# Rapid and robust identification of sepsis using SeptiCyte RAPID in a heterogenous patient population

**DOI:** 10.1101/2024.08.26.24312552

**Authors:** Robert Balk, Annette M. Esper, Greg S. Martin, Russell R. Miller, Bert K. Lopansri, John P. Burke, Mitchell Levy, Richard E. Rothman, Franco R. D’Alessio, Venkataramana K. Sidhaye, Neil R. Aggarwal, Jared A. Greenberg, Mark Yoder, Gourang Patel, Emily Gilbert, Jorge P. Parada, Majid Afshar, Jordan A. Kempker, Tom van der Poll, Marcus J. Schultz, Brendon P. Scicluna, Peter M. C. Klein Klouwenberg, Janice Liebler, Emily Blodget, Santhi Kumar, Xue W. Mei, Krupa Navalkar, Thomas D. Yager, Dayle Sampson, James T. Kirk, Silvia Cermelli, Roy F. Davis, Richard B. Brandon

**Author notes:** Correspondence (R.B.); (T.D.Y.); (R.B.B.).

## Abstract

**Background/Objective:** SeptiCyte RAPID is a transcriptional host response assay that discriminates between sepsis and non-infectious systemic inflammation (SIRS) with a one-hour turnaround time. The overall performance of this test in a cohort of 419 patients has recently been described [Balk et al., J Clin Med 2024, 13, 1194]. In this study we present results from a detailed stratification analysis in which SeptiCyte RAPID performance was evaluated in the same cohort across patient groups and subgroups encompassing different demographics, comorbidities and disease, sources and types of pathogens, interventional treatments, and clinically defined phenotypes. The aims were to identify variables that might affect the ability of SeptiCyte RAPID to discriminate between sepsis and SIRS, and to determine if any patient subgroups appeared to present a diagnostic challenge for the test.

**Methods:** 1) Subgroup analysis, with subgroups defined by individual demographic or clinical variables, using conventional statistical comparison tests. 2) Principal component analysis and k-means clustering analysis, to investigate phenotypic subgroups defined by unique combinations of demographic and clinical variables.

**Results:** No significant differences in SeptiCyte RAPID performance were observed between most groups and subgroups. One notable exception involved an enhanced SeptiCyte RAPID performance for a phenotypic subgroup defined by a combination of clinical variables suggesting a septic shock response.

**Conclusions:** We conclude that for this patient cohort SeptiCyte RAPID performance was largely unaffected by key variables associated with heterogeneity in patients suspected of sepsis.

## 1. Introduction

Accurate and rapid identification of sepsis is often clinically challenging, in part due to non-specific presenting clinical signs [1,2], patient heterogeneity [3–5], and a lack of timely information [6]. Clinical signs of sepsis are often vague and can include dyspnea, weakness, altered mental status, pain and cough, which are signs commonly associated with other disease conditions including heart failure, stroke and respiratory failure [1,2].

To enable better identification and personalized treatment for sepsis, efforts have been made to identify subclasses of patients based on “phenotypes” [3,7,8] or “endotypes” [9–11]. For example, Seymour et al. [3] described four phenotypes differentiated by clinical parameters such as vasopressor requirement, age, chronic illness, renal dysfunction, inflammation, pulmonary dysfunction, shock and liver dysfunction. As another example, Sinha et al. [8] showed that the “hyperinflammatory” and “hypoinflammatory” phenotypes previously identified in ARDS patients could also be applied to septic shock patients. Categorizing sepsis patients into clinically relevant phenotypes could potentially lead to personalized treatments (precision medicine) and better outcomes [12]. However, successful application of precision medicine in sepsis assumes that this condition can be identified early and accurately in the first place. Traditional sepsis diagnostics that rely on the isolation and/or identification of causative pathogens, such as blood culture, have been shown to lack sensitivity, timeliness and in general have not taken patient heterogeneity into account [13]. More recent efforts to improve sepsis diagnosis have included identification of host immune response biomarkers [14].

SeptiCyte RAPID is a host immune response test which measures mRNA expression levels of two genes, PLAC8 and PLA2G7, using a small peripheral blood sample [15,16]. Results are reported as a “SeptiScore”, on a scale of 0-15 and in four “Bands”, with increasing likelihood of sepsis associated with higher SeptiScores and Bands. The discovery of these biomarkers was achieved via machine learning on a heterogenous patient dataset [17]. Because the signature discovery process was performed on a heterogeneous set of patients, we hypothesized that the SeptiCyte RAPID signature would continue to perform robustly in independent heterogeneous validation datasets.

In the present study, as a test of the above hypothesis, we investigated the performance of SeptiCyte RAPID in a heterogeneous, critically ill adult patient cohort stratified by demographics, comorbidities and diseases, sources and types of infecting pathogen, therapeutic interventions, and clinical phenotypes. We sought to determine if the performance of SeptiCyte RAPID for differentiating sepsis from infection-negative systemic inflammatory response syndrome (SIRS) was robust and generalizable across different patient subgroups. The subgroups examined included patients with conditions that 1) present with overlapping clinical signs of sepsis, 2) may predispose to sepsis, and 3) could affect performance of a host immune response assay.

## 2. Methods

### Dataset

The dataset for this analysis (N=419 patients) was from the MARS, VENUS and NEPTUNE studies [16]. Stratifications were performed according to sex, age, race/ethnicity, comorbidities and diseases, therapeutic interventions, and clinically defined phenotypes.

### SeptiCyte RAPID Performance vs. Comparator

The performance of SeptiCyte RAPID for discrimination of sepsis vs. SIRS was evaluated using Retrospective Physician Diagnosis (RPD) as the comparator. The RPD process is a clinical evaluation by a panel of three expert clinicians not involved in the care of the patients [15, 16]. In this study only “forced” RPD was used. That is, if a patient was initially called “indeterminate” by the RPD panelists, the panelists were then forced to make a consensus or unanimous call of sepsis or SIRS.

### Statistics

Statistical tests were mainly conducted with R packages or with Medcalc (medcalc.org). Additional details and cross-checks were as follows. p-values for two-group comparisons were calculated with the Wilcoxon-Mann-Whitney test as implemented in the R ‘stats’ package [18] unless otherwise noted. Some p-value calculations for small N strata were conducted with Student’s t-test as implemented in Microsoft Excel, and also with the Mann-Whitney U test as implemented in Medcalc (medcalc.org) and cross-checked with the web applet at www.socscistatistics.com/tests/mannwhitney/default2.aspx. Proportions tests were conducted with the R ‘stats’ package and cross-checked with Medcalc. One-way ANOVA was conducted with the R ‘stats’ package. Cohen’s kappa was calculated with the web applet at http://vassarstats.net/kappa.html.

Receiver Operating Characteristic (ROC) curve analysis and calculation of area under curve (AUC) values was performed using the pROC library in R [19] and cross-checked with JROCFIT [http://www.rad.jhmi.edu/jeng/javarad/roc/JROCFITi.html] and Medcalc. Confidence intervals for AUC were calculated by the Binomial Exact method, the method of Hanley & McNeil [20], or the method of DeLong et al. [21] as implemented in Medcalc, or by bootstrapping as implemented with the web applet of Skalsk’a & Freylich [22] at http://www.freccom.cz/stomo/input.php. The Hanley & McNeil CI values were cross-checked with the web applet at https://riskcalc.org/ci/.

AUC comparisons were performed with either DeLong’s test [21] or the bootstrap method, both implemented in R. Note that the ROC analysis of small sample sets (AUC and 95% CI) relies on the equivalence relation AUC = U/(n_1_*n_2_) where U = the Mann-Whitney U statistic and n_1_, n_2_ are the sizes of the SIRS and sepsis groups ( see refs. [23, 24] and Supplementary Material, Section 6).

*<u>Principal components analysis (PCA)</u>* was performed using the FactoMineR package in R [25]. The following 16 quantitative clinical variables measured within the first 24 hours of ICU admission defined the dimensions (vectors) by which patients were separated in n-dimensional space: temperature (min & max), heart rate (HR, min & max), respiratory rate (RR, min & max), mean arterial pressure (MAP, min & max), glucose (min & max), white blood cells (WBC, min & max), platelets (min), procalcitonin (PCT), lactate and age. Each of these variables had <=33% missing data, and missing values were replaced with mean values before conducting the PCA. SeptiScore and RPD category were not used in the analysis.

The following qualitative or ordinal variables (not used in constructing the PCA dimensions) were also mapped onto the PCA plot: sex, vasopressor use, mechanical ventilation, pathogen type (bacterial, viral, fungal) and source of infection (blood, urine, sputum, or other), SOFA score, SOFA component scores, and qSOFA & component scores. Missing values for the ordinal variables SOFA, SOFA components and qSOFA were replaced with mean values, as for the quantitative variables.

*<u>Hierarchical clustering (HC)</u>* was then performed upon the PCA according to the method described in refs. [26–28]. This method allowed the PCA-based separation of patients to be further stratified into subgroups, based on combinations of all the variables (quantitative, qualitative, and ordinal) defined above.

*<u>k-means clustering:</u>* The k-means clustering algorithm, implemented in the stats package in R, was applied to either the sepsis group (N=176; results in the main text), or to the entire cohort (Sepsis+SIRS) (N=419; results in the Supplementary Material). Many of the same variables used for the PCA analysis were also used for the k-means, including: temperature (min & max within 24 hours), heart rate (HR, min & max), respiratory rate (RR, min & max), mean arterial pressure (MAP, min & max), glucose (min & max), white blood cell count (WBC, min & max), platelets (min), procalcitonin (PCT), lactate, SOFA score, individual SOFA component scores, qSOFA score, age, sex, vasopressor use (Y/N), mechanical ventilation (Y/N), culture/PCR result (+/-) for bacterial, viral or fungal pathogen, source of positive culture/PCR result (urine, blood, sputum, other). To impute missing data for each variable, the overall mean value for that variable was used. Prior to clustering, the continuous variables were transformed to have a mean of zero and a standard deviation of one. SeptiScore and RPD category were not used as input variables.

## 3. Results

### 3.1. Demographics

Patient stratification was performed according to sex, age, and race/ethnicity. Patient ages ranged from 18 – 90 years and were binarized into subgroups <60 years or ≥ 60 years of age. The performance of SeptiCyte RAPID for discriminating sepsis vs. SIRS never fell below AUC 0.80 for any subgroup. For each demographic comparison, the p-value for discriminating sepsis vs. SIRS was significant at p<0.05. When considered together in a multiple comparison context, the p-values all remained significant (p<0.05/8) after applying a Bonferroni correction. No significant AUC differences were observed between subgroups except for a marginal difference for White vs. Hispanic (p = 0.03) (**Table 1**).

**Table 1.** SeptiCyte RAPID performance for sepsis / SIRS discrimination for patients stratified by sex, age, or race/ethnicity. Performance quantified by AUC and p-value using forced adjudication. Comparison of AUC values by DeLong’s test [21]. AUC confidence intervals (CI) by formula of Hanley & McNeil [20] as computed by the applet at https://riskcalc.org/ci/.

| Condition | N sepsis | N SIRS | AUC | AUC 95% CI | p-value | DeLong’s p-value |
| --- | --- | --- | --- | --- | --- | --- |
| male (M) | 95 | 137 | 0.81 | 0.75 - 0.87 | 1.4 E-15 | M vs. F: 0.52 |
| female (F) | 81 | 106 | 0.84 | 0.78 - 0.90 | 1.6 E-15 |  |
| age < 60 years | 77 | 138 | 0.83 | 0.77 - 0.89 | 3.9 E-14 | Age (< 60) vs. (≥ 60): 0.82 |
| age ≥ 60 years | 99 | 105 | 0.82 | 0.76 - 0.88 | 5.7 E-16 |  |
| Black (B)* | 45 | 70 | 0.85 | 0.77 - 0.93 | 5.9 E-11 | B vs. W: 0.30<br>B vs. H: 0.21<br>B vs. A: 0.62<br>W vs. H: 0.03<br>W vs. A: 0.23<br>H vs. A: 0.63 |
| White (W)* | 108 | 146 | 0.80 | 0.74 - 0.86 | 7.2 E-16 |  |
| Hispanic (H)* | 10 | 12 | 0.93 | 0.81 - 1.05 | 0.00014 |  |
| Asian (A)* | 10 | 11 | 0.89 | 0.74 - 1.04 | 0.0013 |  |

\* Seven patients were unclassified as to race/ethnicity, so were left out of this analysis.

Upon further analysis, a significant White vs. Black difference in SeptiScore performance (p<0.003) was observed for the discrimination of *<u>septic shock</u>* vs. SIRS. For this stratification, the White demographic subgroup gave AUC = 0.83, while the Black subgroup gave AUC = 0.96. See Supplementary Material, Section 1.

### 3.2. Comorbidities and disease

Patients were stratified by the presence or absence of different comorbidities or diseases, specifically hyperglycemia, impaired immunity, hypertension, cardiovascular disease, kidney disease and obesity. These comorbidities and diseases were chosen for analysis based on available patient numbers, presumed influence on sepsis predisposition, and potential for presenting clinical signs that overlap with those of SIRS or sepsis. The ‘impaired immunity’ category included patients with organic immune deficiencies, cancer patients on immunosuppressant drugs, and patients that received immunomodulators such as glucocorticoids; some patients fell in more than one sub-category. Patients for whom a particular co-morbidity or disease was not mentioned in the physician notes were assumed to not have the condition. Point-estimate AUCs ranged from 0.81-0.86 with the only exceptions being AUC 0.79 for patients categorized as hypertensive, and AUC 0.75 for patients with diabetic hyperglycemia (**Table 2**). Box and whisker plots corresponding to the entries of Table 2 are shown in **Figure 1 (A-F).** By DeLong’s test there were no significant AUC differences in SeptiCyte performance between patients with vs. without the stated condition.

**Figure 1:**
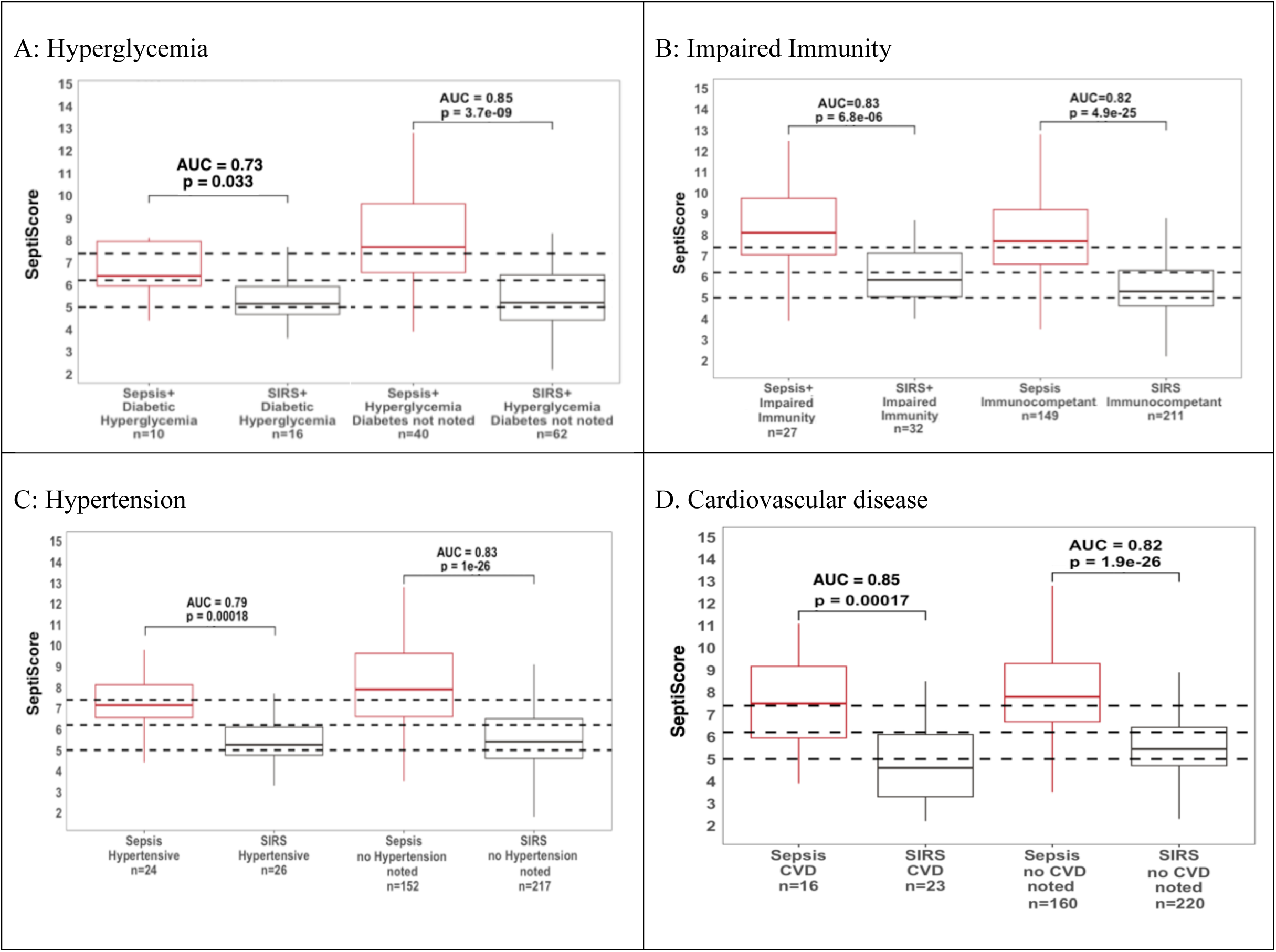
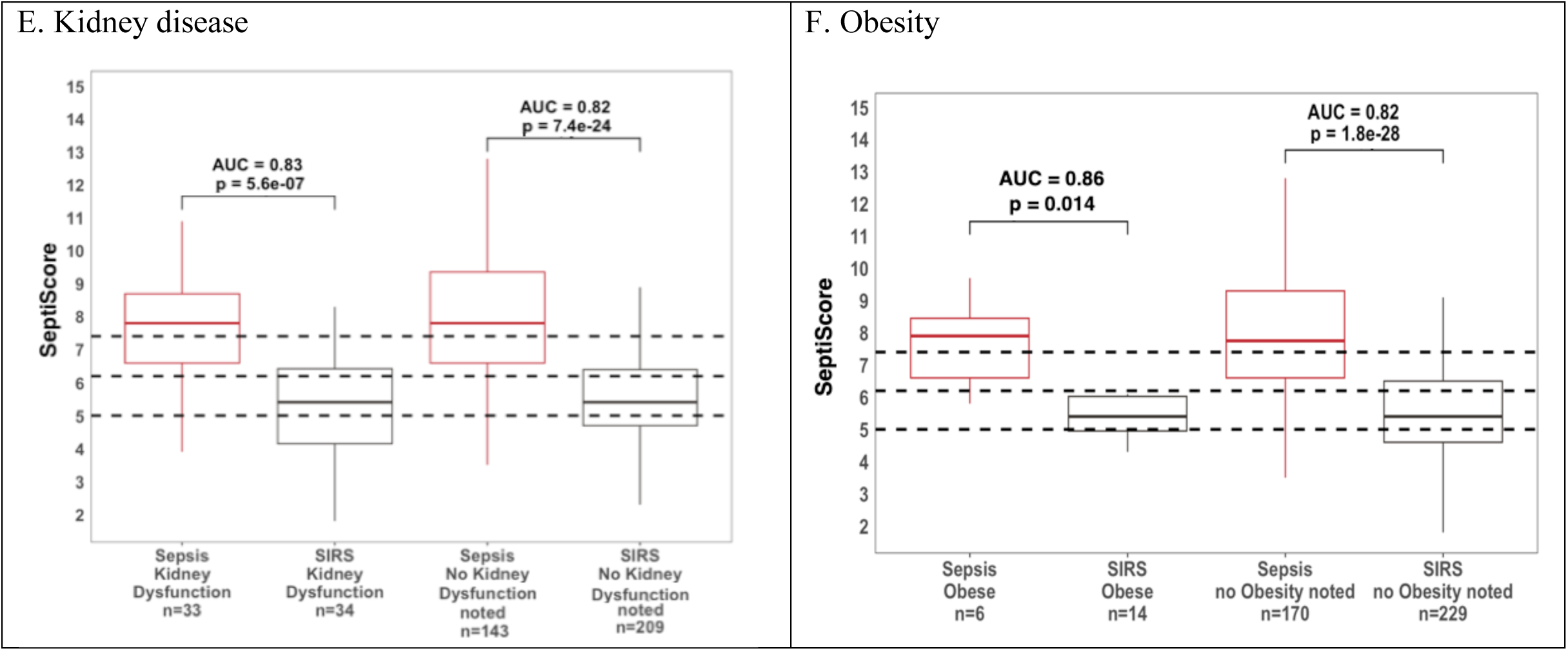
Box and whisker plots of SeptiCyte RAPID performance in patients with (A) hyperglycemia, (B) impaired immunity, (C) hypertension, (D) cardiovascular disease, (E) kidney disease, (F) obesity.

**Table 2.** SeptiCyte RAPID performance stratified by comorbidities and disease. AUC confidence intervals (CI) by formula of Hanley & McNeil [20] as computed by applet: https://riskcalc.org/ci/. Abbreviations: HG, hyperglycemia; II, impaired immunity; HT, hypertension; CVD, cardiovascular disease; KD, kidney disease. For small sample comparisons, the p-value was calculated using both the t-test (T) and the Mann-Whitney U test (U).

| Stratification | N sepsis | N SIRS | AUC | AUC 95% CI | p-value | DeLong's p-value |
| --- | --- | --- | --- | --- | --- | --- |
| hyperglycemia (HG) | 50 | 78 | 0.83 | 0.76 - 0.92 | 5.3 E-10 | <u>HG vs. no HG noted: 0.76</u> |
| diabetic hyperglycemia | 10 | 16 | 0.75 | 0.51 - 0.93 | 0.035 (T)<br>0.038 (U) |  |
| diabetic hyperglycemia not noted | 40 | 62 | 0.85 | 0.77 - 0.93 | 3.7 E-09 |  |
| no hyperglycemia noted (no HG) | 126 | 165 | 0.82 | 0.76 - 0.86 | 5.8 E-21 |  |
| impaired immunity (II) | 27 | 32 | 0.83 | 0.72 - 0.94 | 6.8 E-06 | <u>II vs. No II noted: 0.84</u> |
| no impaired immunity noted | 149 | 211 | 0.82 | 0.77 - 0.87 | 4.9 E-25 |  |
| hypertension (HT) | 24 | 26 | 0.79 | 0.66 - 0.92 | 1.8 E-04 | <u>HT vs. no HT noted: 0.59</u> |
| no hypertension noted | 152 | 217 | 0.83 | 0.78 - 0.88 | 1.03 E-26 |  |
| cardiovascular disease (CVD) | 16 | 23 | 0.85 | 0.72 - 0.98 | 1.7 E-04 | <u>CVD vs. no CVD noted: 0.63</u> |
| cardiovascular disease (acute) | 7 | 6 | 1.0 | 0.75 - 1.0 | 0.0031 (T)<br>0.0034 (U) |  |
| cardiovascular disease (chronic) | 9 | 17 | 0.81 | 0.62 - 1.00 | 0.0046 (T)<br>0.0105 (U) |  |
| no cardiovascular disease noted | 160 | 220 | 0.82 | 0.78 - 0.86 | 1.9 E-26 |  |
| kidney disease (KD) | 33 | 34 | 0.83 | 0.73 - 0.93 | 5.6 E-07 |  |
| kidney disease (acute) | 13 | 13 | 0.86 | 0.70 - 1.00 | 0.0012 (T)<br>0.0023 (U) | <u>KD vs. no KD noted: 0.89</u> |
| kidney disease (chronic) | 20 | 21 | 0.83 | 0.70 - 0.96 | 0.0001 T)<br>0.0004 (U) |  |
| no kidney disease noted | 143 | 209 | 0.82 | 0.77 - 0.87 | 7.4 E-24 |  |
| obesity (BMI ≥ 30) | 6 | 14 | 0.86 | 0.66 - 1.06 | 0.0141 (T)<br>0.0135 (U) | Obesity vs. no obesity noted:<br>0.63 |
| no obesity noted | 170 | 229 | 0.82 | 0.78 - 0.86 | 1.8 E-28 |  |

We note several limitations to this analysis, because of small sample sizes. The AUC value of 1.0 for patients with acute cardiovascular disease should be considered imprecise, because of low N. Additionally, while the uncorrected p-value for each individual comparison falls below the conventional cutoff for significance (p∼0.05), if we consider as a group the nine comorbidity/disease comparisons with low numbers, then upon applying a Bonferroni correction, the p-values for diabetes and obesity now fall above the adjusted significance cutoff (p∼0.05/9) and the p-value for chronic CVD becomes borderline significant.

Hyperglycemia - The cohort was stratified based on whether or not patients had hyperglycemia (>=200 mg/dl) (**Figure 1A**). A few patients with hypoglycemia were also noted, but their numbers were too small (14 sepsis, 3 SIRS) for reliable statistical analysis. Hyperglycemia included those patients with and without diabetes (as indicated in the physician notes). For patients without diabetes indicated, data were based on “Glucose.Min” or “Glucose.Max” values recorded over a 24-hour period. The SeptiCyte RAPID performance for patients with diabetic hyperglycemia (AUC 0.73) appeared marginally lower than for all hyperglycemic patients (AUC 0.83) or for non-hyperglycemic patients (AUC 0.82). Note that, although the p-value for sepsis vs. SIRS discrimination in the diabetic hyperglycemia subgroup (p ≈ 0.04) falls below the conventional cutoff (p∼0.05), after a Bonferroni correction this p-value now falls above the adjusted significance cutoff (p∼0.05/9).

Impaired immunity - The cohort was stratified based on whether or not patients could be considered to have impaired immunity (**Figure 1B**). The category of impaired immunity included use of immunosuppressants (including corticosteroids), adrenal insufficiency, splenectomy, asplenia, HIV/AIDS and cancer. Of the 13 patients with cancer, no consideration was given to the type of cancer, or whether the patients were being treated with cancer therapy, or the duration of cancer therapy. White blood cell counts (WBC, min) in patients with impaired immunity as defined above ranged from 300 – 37,000 cells / uL, and for patients with no such impaired immunity the range was 300 – 52,000 cells / uL. SeptiCyte RAPID AUCs were 0.83 and 0.82 for patients with impaired immunity vs immunocompetent respectively.

Hypertension - The cohort was stratified based on whether hypertension was noted in the physician comments (**Figure 1C**). The AUC for differentiating sepsis from SIRS was 0.79 for hypertensive patients and 0.83 for those without hypertension noted, a difference that was not significant according to DeLong’s test (p=0.59).

Cardiovascular disease (CVD) - The cohort was stratified based on whether patients had been diagnosed with CVD (**Figure 1D**). CVD patients were further subdivided into those with acute (cardiac arrest) vs. chronic disease. The chronic CVD patients included those with congestive heart failure, aortic valve replacement, and bradycardia with cardiogenic shock. Of the patients with CVD, there were 16 with sepsis, and 23 with SIRS. The remainder of the cohort (without CVD) consisted of 160 patients with sepsis and 220 with SIRS. We calculated AUC 0.85 for patients with CVD as opposed to AUC 0.82 for patients without CVD. This difference was deemed not significant by DeLong’s test (p = 0.63).

Kidney disease – the cohort was stratified into those with kidney disease (acute or chronic) vs. no kidney disease noted (**Figure 1E**). Kidney disease included patients with acute renal failure/injury vs. patients with chronic conditions (renal insufficiency/ disease, end stage renal disease, renal cell carcinoma, nephrolithiasis with bilateral ureteral stents, or chronic dialysis). AUCs for differentiating sepsis from SIRS were 0.82 (no kidney disease noted) and 0.83 for those with chronic kidney disease, a difference that was not significant.

Obesity - the cohort was stratified into obese (BMI>=30 or obesity noted in physician comments), vs. BMI<30 or no obesity noted (**Figure 1F**). Obesity is a known risk factor for sepsis [29, 30]. AUCs for differentiating sepsis from SIRS were 0.86 (obese patients) and 0.82 for non-obese patients; DeLong’s test indicates this AUC difference is not significant. Note that, while the p-value for sepsis vs. SIRS discrimination in the obese subgroup (p ≈ 0.014) falls below the conventional cutoff (p∼0.05), after a Bonferroni correction this p-value now falls above the adjusted significance cutoff (p∼0.05/9).

### 3.3 Source and Type of Infection

#### 3.1.1 Infection source

Of the 176 sepsis patients in the cohort, 150 had an identified source of infection, specifically pulmonary (n=59), abdominal (n=30), blood (n=17), central nervous system (CNS) (n=6), urinary tract (UTI) (n= 24) and “other source” (n=14, as detailed in legend of Figure 2). There were 26 sepsis patients for whom an initial source of infection could not be identified (NI). In the cohort, 243 patients were retrospectively determined to have SIRS (deemed non-infectious), however only 215 did not have a source of infection identified. **Figure 2** presents box and whisker plots for subgroups of septic patients vs. the SIRS patients, with AUCs and p values indicated. SIRS patients with an identified source of infection (4 abdominal, 5 CNS, 15 pulmonary, 2 urinary, 2 other) were excluded from the analysis. For all sepsis subgroups with identified sources of infection, the median SeptiScore was in Band 4 (highest sepsis probability), as defined in the banding scheme of Balk et al. [16]. For septic patients without an identified source of infection, the median SeptiScore was in Band 3. For the subgroups with sufficient numbers to allow for a reliable AUC comparison, DeLong’s test showed no significant differences (pulmonary vs. NI, p=0.08; abdominal vs. NI, p=0.21; UTI vs. NI, p=0.35).

**Figure 2.**
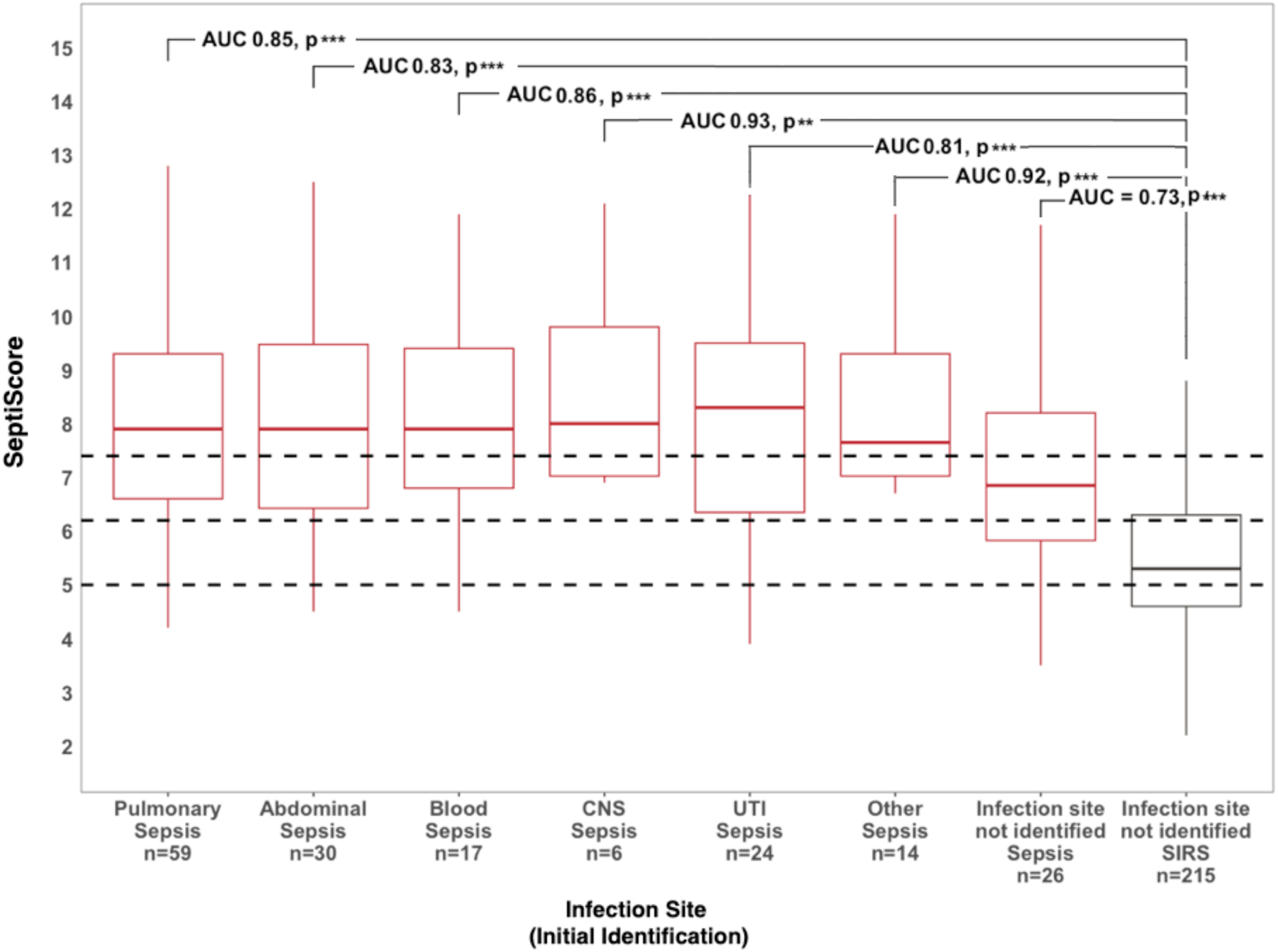
SeptiCyte RAPID performance stratified by infection source. The “other” group included the following (N per group): cellulitis (3), Fournier’s gangrene (1), hip arthroplasty (1), osteomyelitis (1), toe infection (1), sacral wound (1), post-surgical sternal wound (1), bladder/prostate abscess/ peritonitis from cecum micro perforation (1), influenza (1), tracheitis (1), erysipelas (1), skin or soft tissue necrotizing fasciitis (1). Significance: p<=0.01**, p<=0.001***.

#### 3.1.2 Infection type

We previously determined that SeptiCyte RAPID performance was not affected by the Gram (+/-) category of the sepsis pathogen (see Section 9 of the Supplement to Balk et al. [16]). In the present study, we extended this analysis to determine if different sepsis pathogens were associated with different sources of infection. Results of this analysis are provided in **Supplementary Material, Section 2.** To summarize, viral pathogens were more often detected in patients with pulmonary sepsis, which may reflect a bias in the types of pathogen identification tests ordered for this subgroup. *Staphylococcus aureus* was the most frequently isolated pathogen (19% of all pathogen detection events) and was mostly associated with pulmonary and blood sources of septic infection. Gram negative pathogens, in particular *Escherichia coli* and *Pseudomonas aeruginosa*, were most isolated in urosepsis.

### 3.4. Therapeutic Interventions

We examined the performance of SeptiCyte RAPID after stratification along two treatment dimensions: either (+/-) pharmaceuticals (immunosuppressants; antibiotics, inotropes, vasopressors); or (+/-) mechanical ventilation. Pharmaceutical treatments were selected from a list of >400 drugs noted in the patient records. Further detail on the immunosuppressants, antibiotics, anti-neoplastics, inotropes and vasopressors listed in patient records is provided in the **Supplementary Material, Section 3.**

Results in **Table 3** and **Figure 3** show that the use of a broad range of immunosuppressants did not affect the overall performance of SeptiCyte RAPID in our cohort. AUC values in patients treated with immunosuppressants (0.80) or not treated (0.82) were not statistically different (p=0.75 by DeLong’s test).

**Figure 3:**
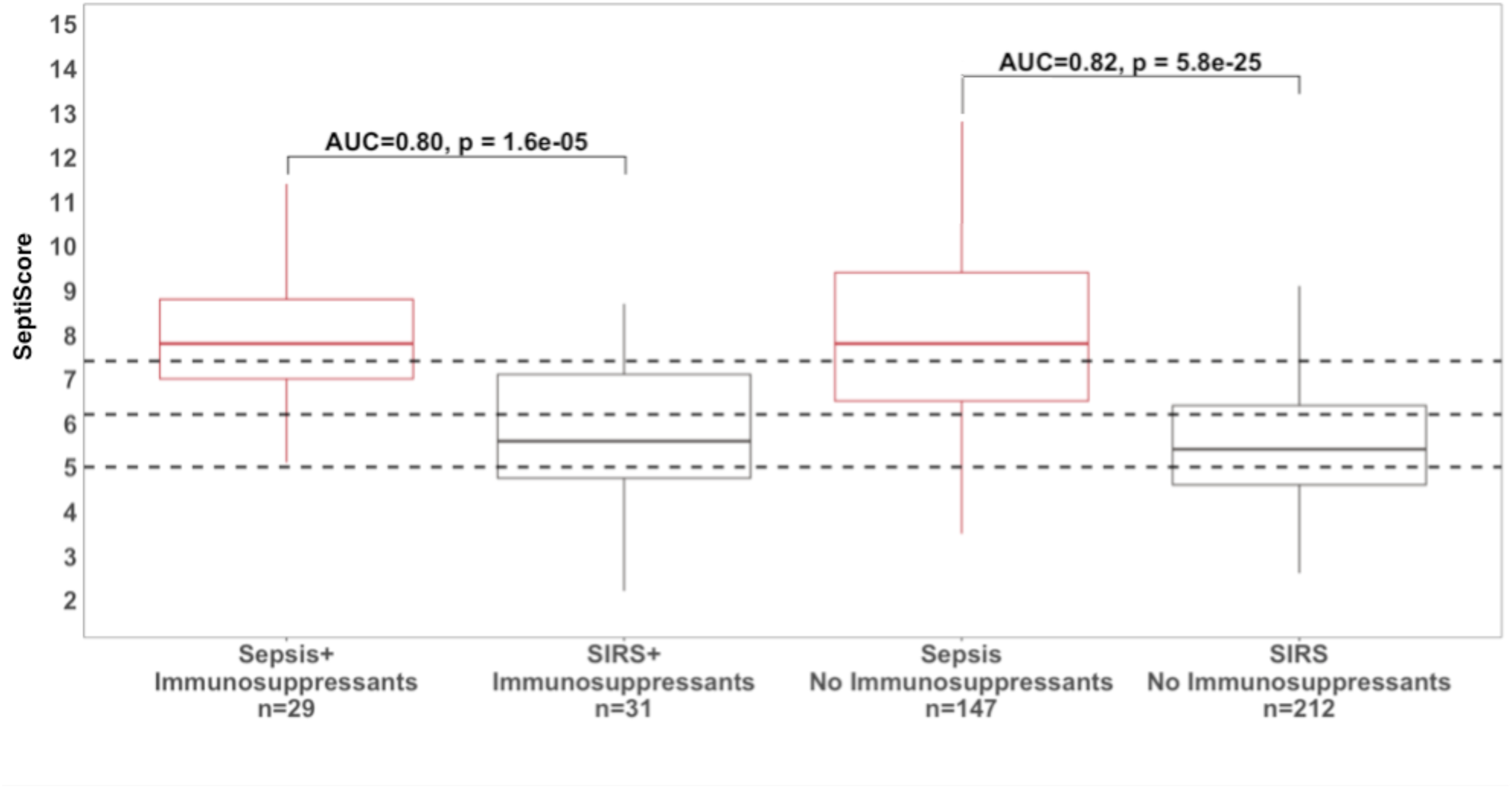
Box and whisker plot of SeptiCyte RAPID performance for patients treated vs. not treated with immunosuppressants.

**Table 3.** SeptiCyte RAPID performance stratified by therapeutic interventions. Abbreviations: Abx, antibiotics; I, inotropes; IS, immunosuppressants; MV, mechanical ventilation; V, vasopressors. AUC confidence intervals (CI) by formula of Hanley & McNeil [20] as computed by applet: https://riskcalc.org/ci/.

| **Therapeutic Intervention** | **N sepsis** | **N SIRS** | **AUC (95% CI)** | **sepsis vs. SIRS**<br>**p-value** | **DeLong's p-value** |
| --- | --- | --- | --- | --- | --- |
| Immunosuppressants (IS) | 29 | 31 | 0.80 (0.67-0.91) | 1.6 E-05 | IS vs. no IS: 0.75 |
| No immunosuppressants | 147 | 212 | 0.82 (0.77-0.87) | 5.8 E-25 |  |
| Antibiotics (Abx) 1 day prior to SeptiScore | 110 | 80 | 0.79 (0.73-0.85) | 7.5 E-13 | Abx prior vs. on = 0.46<br>Abx on vs. after = 0.71<br>Abx after vs. prior = 0.79 |
| Antibiotics same day as SeptiScore | 34 | 42 | 0.84 (0.75-0.93) | 4.9 E-08 |  |
| Antibiotics 1 day after SeptiScore | 29 | 119 | 0.81 (0.71-0.91) | 9.4 E-06 |  |
| Vasopressors (V) or inotropes (I) | 81 | 76 | 0.83 (0.77-0.89) | 2.3 E-14 | V/I vs. no V/I: 0.60 |
| No vasopressors (V) or inotropes (I) | 95 | 167 | 0.81 (0.75-0.87) | 4.8 E-15 |  |
| Mechanical ventilation (MV) | 62 | 91 | 0.80 (0.72-0.88) | 3.1 E-10 | MV vs. no MV: 0.36 |
| No mechanical ventilation | 114 | 152 | 0.84 (0.79-0.89) | 5.4 E-21 |  |

Patients treated with antibiotics (Abx) were subdivided into three groups based on when the antibiotic treatment was started relative to SeptiCyte RAPID blood sampling. In total, 190 patients (110 sepsis, 80 SIRS) were given antibiotics up to one day prior, 76 patients (34 sepsis, 42 SIRS) were given antibiotics on the same day, and 148 (29 sepsis, 119 SIRS) received antibiotics on the day following the blood sampling. SeptiCyte RAPID AUCs were not significantly affected by timing of antibiotics over these time periods, with AUCs ranging from 0.79 to 0.84 (**Figure 4**). The purpose of this comparison was to determine if antibiotics interfered with SeptiCyte RAPID performance in a restricted time window from −1 days to +1 day relative to blood draw at day 0. We limited the analysis to this relatively narrow time window, because a recent study [31] indicated that a reduction in SeptiScore is expected over the period 2-6 days following initiation of antibiotics.

**Figure 4:**
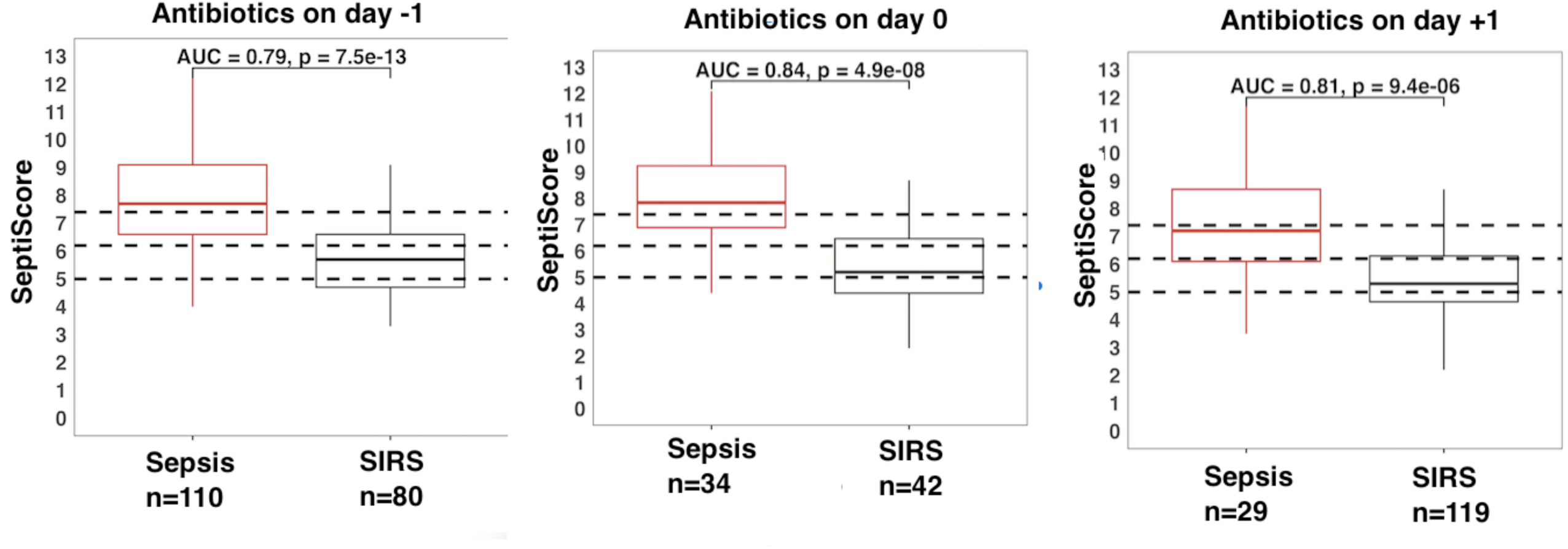
Box and whisker plots of the influence of antibiotic treatment (Abx) initiation time on SeptiCyte RAPID performance. The day of blood draw is defined as day 0. (A) treatment initiated −1 days to 0 days relative to blood draw; (B) antibiotic treatment initiated on the same day as blood draw. (C) antibiotic treatment initiated +1 day after blood draw.

Use of vasopressors or inotropes had no evident effect on SeptiCyte RAPID performance (AUC 0.83 versus AUC 0.81 in the absence of vasopressors or inotropes, p= 0.60 by DeLong’s test). Similarly, SeptiScores in patients on mechanical ventilation (AUC 0.80) did not differ significantly from patients not on mechanical ventilation (AUC 0.84), p= 0.36 by DeLong’s test.

### 3.5. Phenotypic Subgrouping

We used two unsupervised machine learning methods - Principal Component Analysis / Hierarchical clustering (PCA / HC) and k-means clustering - to conduct stratification analyses. These methods employed combinations of readily available clinical variables as input.

#### 3.5.1. Phenotypic subgrouping of the septic patients (N=176) by PCA / HC

We first conducted a PCA, based on 16 pre-selected clinical variables and excluding SeptiScores and RPD determinations. The first two PCA dimensions captured 15.32% and 12.29%, respectively, of the total variation. A hierarchical clustering (HC) analysis was then performed upon the PCA, which gave a separation into three major subgroups (**Figure 5**). There is additionally a single outlier patient, denoted as “subgroup 4” and represented by the blue point. This patient has a combination of elevated values for Glucose (min & max), lactate, WBC (max), SOFA (Liver component score) & lower values for temperature (min). Of note is that blood culture positive patients did not cluster in any particular subgroup (not shown in figure).

**Figure 5.**
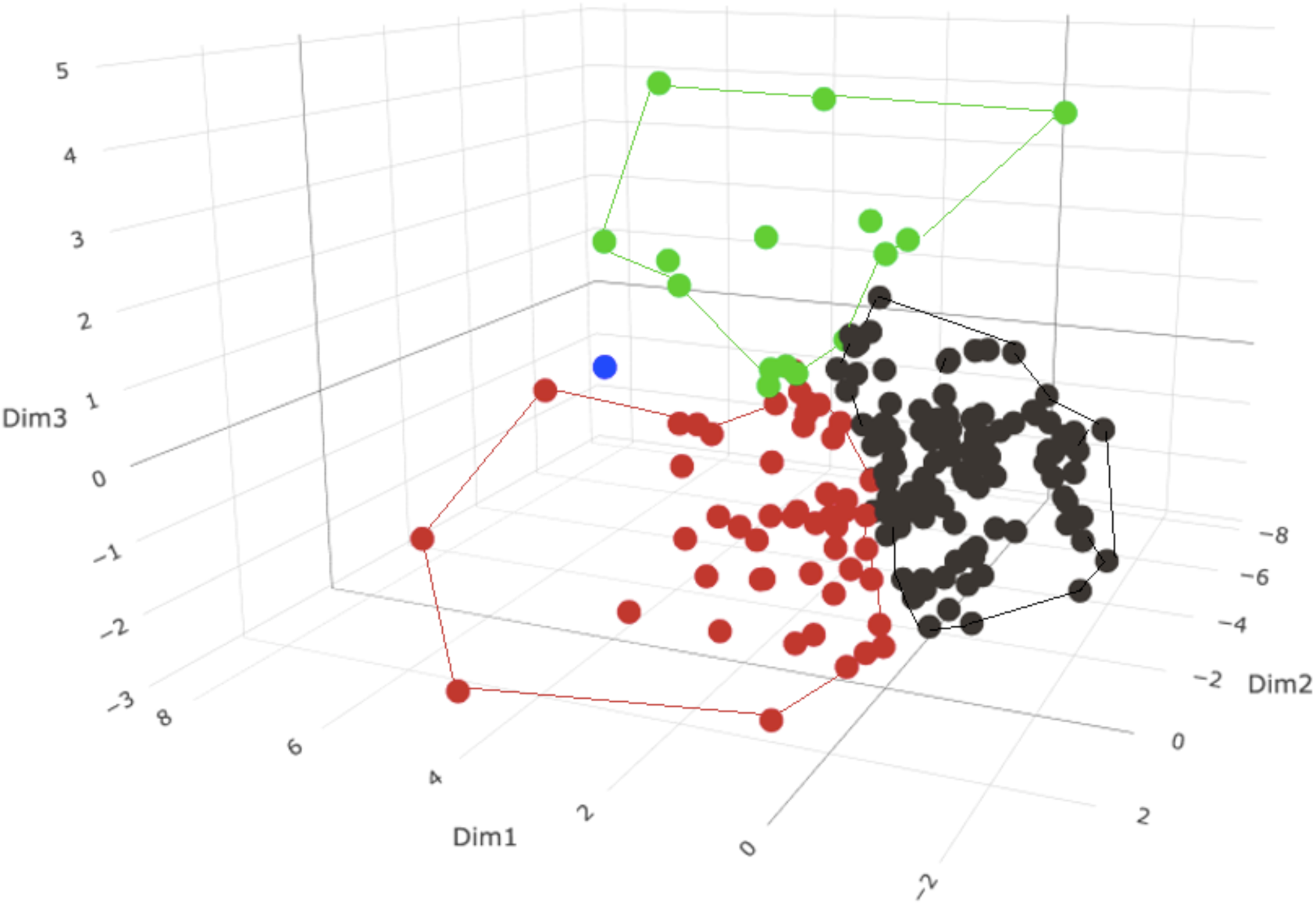
PCA plot of the sepsis group (N=176) with superimposed HC using 16 phenotypic variables. Each variable used in this analysis had <=33% missing data, and mean values were used to impute the missing data. In the plot, the peripheral points in each subgroup were used to define the cluster boundaries for that subgroup. Sepsis subgroup 1 (black) N=110. Sepsis subgroup 2 (red) N=50. Sepsis subgroup 3 (green) N=15. Sepsis subgroup 4 (blue) N=1.

We next evaluated the performance of SeptiCyte RAPID for discriminating the SIRS group (N=243) from each of the three sepsis subgroups defined by PCA / HC. Results are given in **Table 4**. DeLong’s test indicates that SeptiCyte RAPID performance is significantly better for subgroup 3 (AUC 0.93) than for subgroups 1 or 2 (AUC 0.81).

**Table 4.** SeptiCyte RAPID performance in the three subgroups defined by PCA + HCC analysis

| **Comparison** | **N sepsis** | **N SIRS** | **AUC** | **sepsis vs. SIRS p-value** | **DeLong's p-value** |
| --- | --- | --- | --- | --- | --- |
| sepsis PCA subgroup 1 vs. SIRS | 110 | 243 | 0.81 | 6.9 E-19 | 1vs2 p=0.95 |
| sepsis PCA subgroup 2 vs. SIRS | 50 | 243 | 0.81 | 4.5 E-09 | 1vs3 p=0.015 |
| sepsis PCA subgroup 3 vs. SIRS | 15 | 243 | 0.93 | 2.2 E-06 | 2vs3 p=0.035 |

Subgroup 4 only had 1 patient, so was not included in this table.

To identify the phenotypic variables that cause the three subgroups to differ - and particularly to identify the variables that make subgroup 3 distinct from subgroups 1 and 2 - we conducted an ANOVA. Phenotypic variables with greatest statistical significance (p<0.01) are presented in **Table 5**. This analysis revealed that patients in subgroup 3 appeared the most seriously ill as indicated by relatively higher SOFA, PCT, RR (min & max), HR (max), WBC (max), lactate values and vasopressor use, and relatively lower MAP (min), glucose (min), platelets (min) and temperature (min). Of the 15 patients in subgroup 3, a total of 11 were clinically diagnosed with septic shock and 3 with severe sepsis based on sepsis-2 definition with no viral infections. Notably, we find no significant differences between the three sepsis subgroups with respect to site of infection or type of infecting pathogen.

**Table 5.**
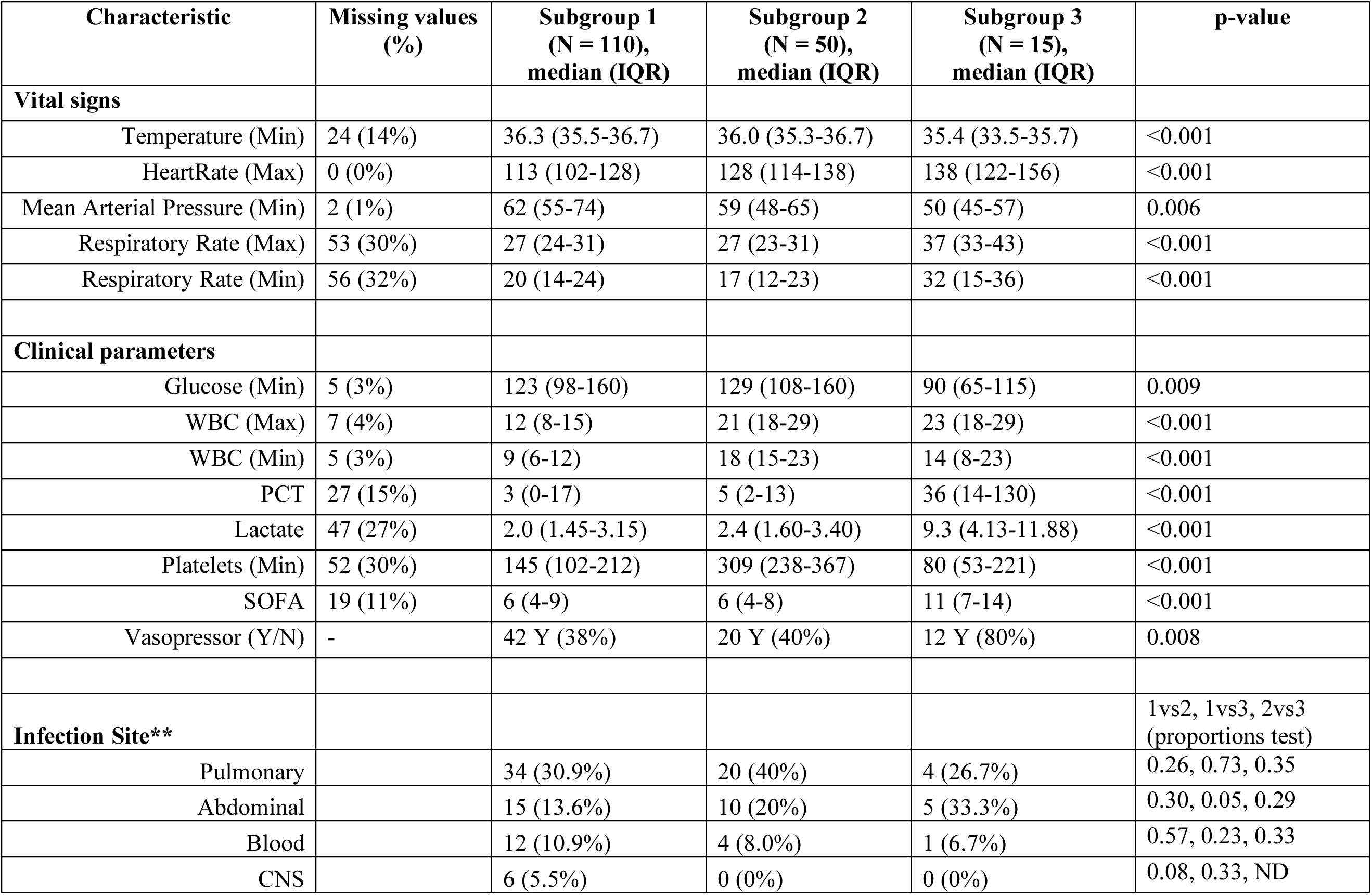
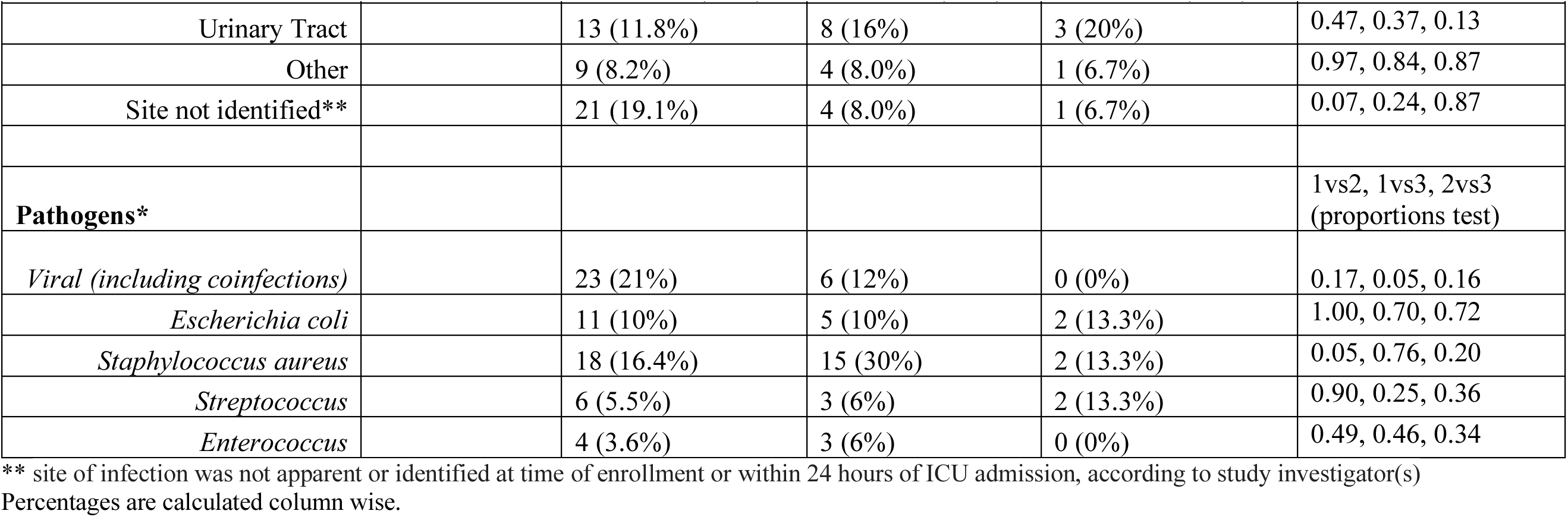
Phenotypic characteristics of patients in the three subgroups from the PCA / HC analysis). p-values were calculated by one-way ANOVA for quantitative variables, and by a proportions test for categorical variables.

In the sepsis group, the PCA variables contributing greater than average (>6%) phenotypic variability for dimensions 1 and 2 were WBC (min & max), glucose (min & max), platelets (min), Age, Temperature (min) and MAP (min). We analyzed SeptiCyte RAPID performance in different subgroups defined by either these individual driving variables or other biomarkers used for sepsis adjudication (**Figure 6**). Similar SeptiCyte RAPID performance levels (AUC 0.79-0.85) were observed for most subgroups, indicating that SeptiCyte RAPID should still have utility regardless of the values of these driving variables. The highest SeptiScore AUCs (0.85) were found for patient subgroups having high WBC counts (>12E+06/mL) or normal platelet counts (150,000 - 450,000/uL). The lowest SeptiScore AUC (0.71) was found for the subgroup with PCT <0.5 ng/mL. This low PCT subgroup contained 32 sepsis patients which appeared less seriously ill than the high PCT (>0.5 ng/mL) subgroup, as indicated by lower mean SOFA score, and a lower proportion on mechanical ventilation. The low PCT sepsis subgroup also had fewer patients that were bacterial-culture positive (40.6%), and a higher percentage of patients that were viral positive (31.3%), as compared to the high PCT subgroup which had 66% bacterial-culture positive (p<0.01) and only 15% viral positive (p<0.03). The detailed quantitative analysis corresponding to Figure 6 is presented in the **Supplementary Material, Section 5, Table S5.** We note that the individual clinical parameters driving separation in the PCA were not themselves effective at discriminating sepsis vs. SIRS (AUC 0.5-0.6), consistent with the previous analysis of Balk et al. [16].

**Figure 6:**
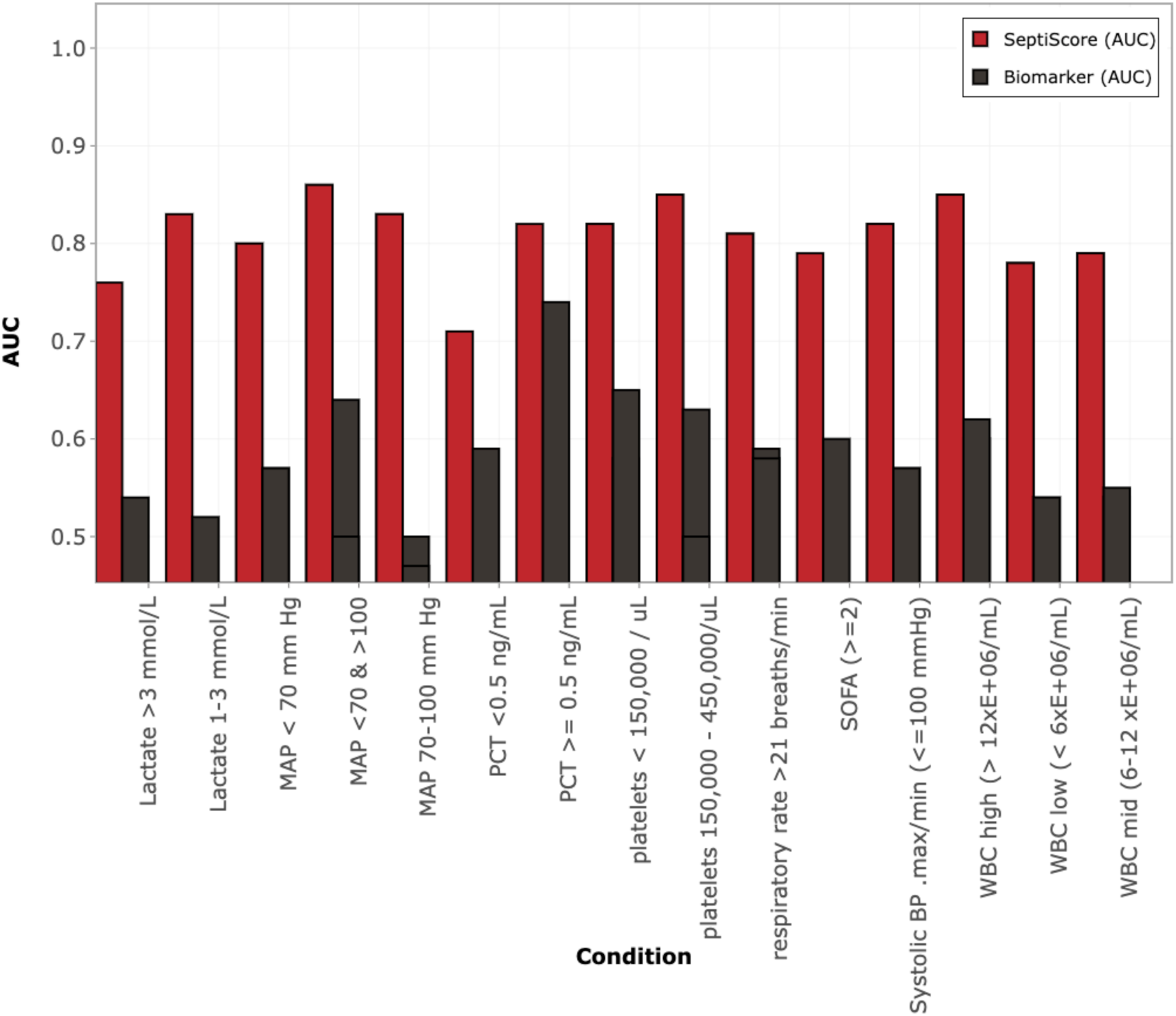
Performance for discriminating sepsis vs. SIRS in different phenotypic subgroups defined by individual driving variables in PCA / HC and other biomarkers used for sepsis adjudication. Red: SeptiScore. Black: driving variable. Additional details corresponding to this figure are presented in Supplement Table S5.

#### 3.5.2. Phenotypic subgrouping of the septic patients (N=176) by k-means clustering

We conducted another stratification of the septic patients using a different approach (k-means clustering). Results of this analysis are shown in **Figure 7**. The k-means clustering algorithm indicated that 2 subgroups was an optimal number, and produced a very clear separation of the sepsis patients into the two subgroups. We next conducted an ANOVA to identify which of the phenotypic variables in the analysis provided the most significant discrimination between the two subgroups. Results of the ANOVA are presented in **Table 6**. (For vital signs, clinical chemistry measurements, and interventions only those variables giving p<0.01 are presented.) Interestingly, with the sole exception of viral infections being more prominent in subgroup 1, neither the site of infection, or pathogen type, were significant drivers of the separation between the two k-means subgroups. Also, blood culture positive patients did not cluster in either of the two subgroups (not shown in figure). Compared to subgroup 1, the subgroup 2 appeared more seriously ill, being distinguished by depressed temperature and MAP; elevated WBC, lactate, PCT, and SOFA; and increased administration of vasopressors and mechanical ventilation. Despite these clinical differences between subgroups, SeptiCyte RAPID AUCs for differentiating subgroup 1 sepsis patients (n=96) and subgroup 2 sepsis patients (n=80) from SIRS patients (n=243) were 0.80 and 0.85, respectively.

**Figure 7.**
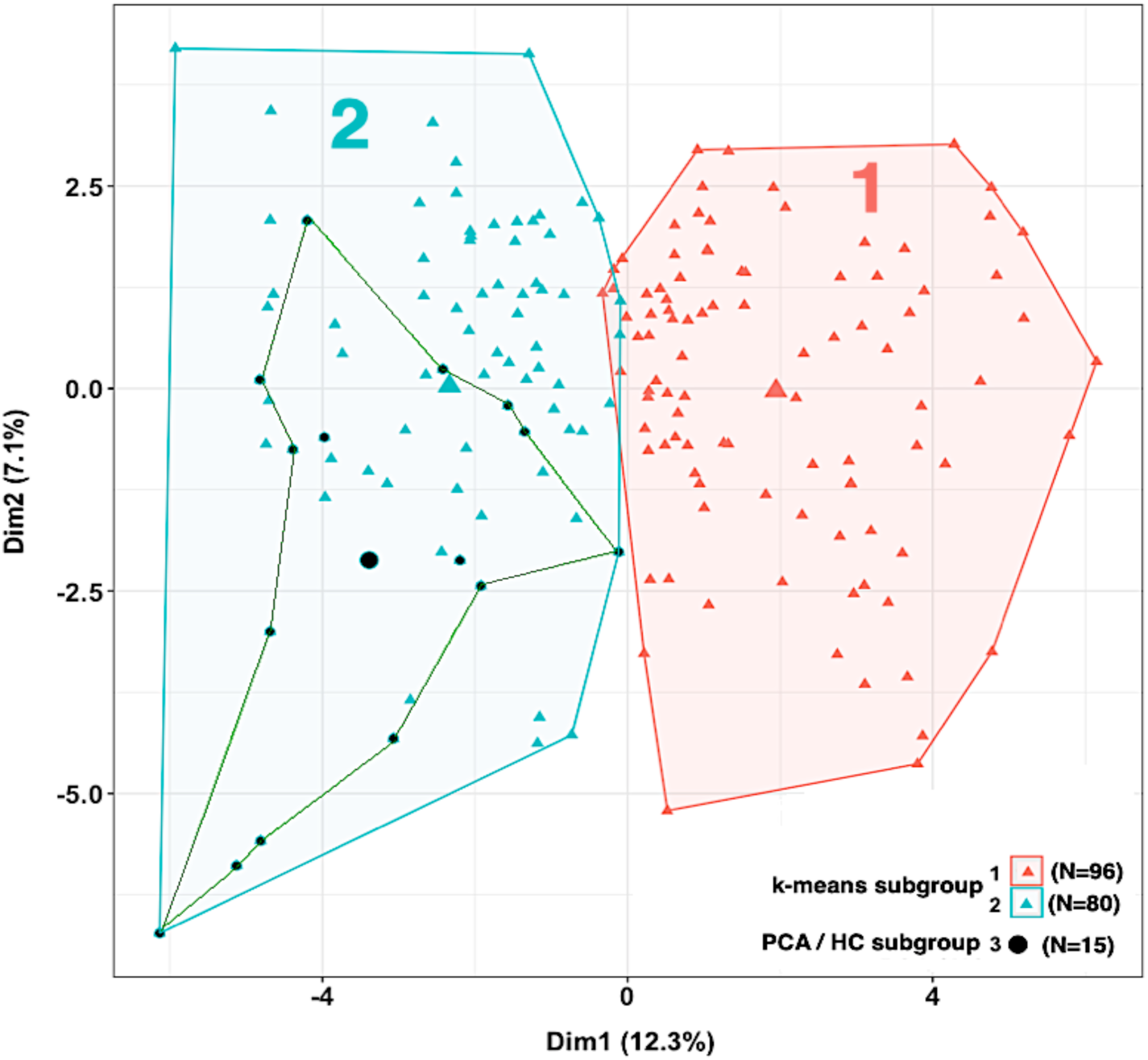
Sepsis subgroups 1 (red; N=96) and 2 (blue; N=80) identified by k-means clustering. The small, most seriously ill subgroup 3 from the PCA / HC analysis (black; N=15) is contained entirely within k-means group 2. The large colored symbols indicate the centroids of the subgroups.

**Table 6.** Phenotypic variables contributing to separation between the two sepsis subgroups in the k-means analysis of Figure 6. p-values were calculated with 1-way ANOVA for quantitative variables, and with Pearson’s Chi-squared test or two-sample proportions test for categorical variables. For vital signs, clinical chemistry measurements and interventions, only those variables giving p<0.01 are presented.

| Characteristic | Missing values (%) | Subgroup 1 (N = 96),<br>median (IQR) | Subgroup 2 (N = 80),<br>median (IQR) | p-value* |
| --- | --- | --- | --- | --- |
| **Vital signs** |  |  |  |  |
| Temperature (Min) | 24 (14%) | 36.4 (36.0-36.8) | 35.3 (34.4-35.8) | <0.001 |
| Mean Arterial Pressure (Min) | 2 (1%) | 67 (57-78) | 55 (46-61) | <0.001 |
| Respiratory Rate (Max) | 53 (30%) | 25 (23-31) | 29 (25-35) | 0.01 |
| **Clinical chemistry** |  |  |  |  |
| WBC (Max) | 7 (4%) | 12 (8-18) | 18 (14-25) | <0.001 |
| WBC (Min) | 5 (3%) | 10 (6-14) | 14 (9-19) | <0.001 |
| Lactate | 47 (27%) | 2.10 (1.45-3.00) | 3.00 (1.83-5.85) | <0.001 |
| GCS<15 (qSOFA component) | 25 (14%) | 18 (18.8%) | 42 (52.5%) | <0.001 |
| qSOFA≥2 | - | 22 (23%) | 68 (85%) | <0.001 |
| qSOFA<2 | - | 62 (65%) | 8 (10%) | <0.001 |
| SOFA | 19 (11%) | 4 (2-6) | 9 (7-12) | <0.001 |
| PCT_binary (>0.5 ng/ml) | 27 (15%) | 52 (64%) | 65 (96%) | <0.001 |
| **Interventions** |  |  |  |  |
| Vasopressors used | - | 15 (16%) | 60 (75%) | <0.001 |
| Mechanical Ventilation | - | 18 (19%) | 44 (55%) | <0.001 |
| **Infection Site\*** |  |  |  |  |
| Pulmonary | - | 36 (37.5%) | 23 (28.8%) | 0.22 |
| Abdominal | - | 13 (13.5%) | 17 (21.2%) | 0.18 |
| Blood | - | 13 (13.5%) | 4 (5%) | 0.06 |
| Central nervous system (CNS) | - | 4 (4.2%) | 2 (2.5%) | 0.54 |
| Other | - | 5 (5.2%) | 9 (11.2%) | 0.14 |
| Urinary tract infection (UTI) | - | 9 (9.4%) | 15 (18.8%) | 0.07 |
| Site not confirmed initially | - | 16 (16.7%) | 10 (12.5%) | 0.44 |
| **Pathogens\*** |  |  |  |  |
| *Viral (including coinfections)* | - | 23 (24%) | 6 (7.5%) | 0.006 |
| *Escherichia coli* | - | 8 (8.3%) | 10 (12.5%) | 0.36 |
| *Staphylococcus aureus* | - | 20 (20.8%) | 15 (18.8%) | 0.74 |
| *Streptococcus* | - | 5 (5.2%) | 6 (7.5%) | 0.53 |
| *Enterococcus* | - | 4 (4.2%) | 3 (3.8%) | 0.89 |

\* Percentages are calculated column wise.

Finally, we performed a Venn diagram analysis to determine overlap(s) between the subgroups identified by PCA / HC stratification vs. k-means stratification. We determined that all 15 of the patients classified as subgroup 3 in the PCA / HC analysis - patients resembling septic shock or severe sepsis according to the Sepsis-2 criteria - fell within k-means subgroup 2, as indicated in **Figure 7**.

## 4. Discussion

It is well known that patients with sepsis are heterogenous with respect to clinical signs, response to therapy, and outcome. Factors contributing to this heterogeneity include patient demographics, comorbidities and diseases, infecting pathogen, locus of the infection, concurrent therapies, and disease progression and stage [32]. For maximum clinical utility, any sepsis diagnostic test should retain a high level of performance when confronted with these sources of variability. Accordingly, we investigated the performance of a commercial gene-expression based sepsis test (SeptiCyte RAPID) across a broad range of patient characteristics. We demonstrated generally consistent and robust diagnostic performance of SeptiCyte RAPID in a heterogenous, adult, critically ill patient population suspected of sepsis.

### Demographics

We found no statistically significant differences in AUC for SeptiCyte performance in discriminating sepsis vs. SIRS with respect to age, sex or ethnic/racial group, when appropriate number of cases per group were analyzed (Table 1). Upon closer examination, however, we did observe a race/ethnicity -based difference in SeptiScore performance for distinguishing *septic shock* from SIRS (AUC 0.83 for septic shock vs. SIRS in Whites; AUC 0.96 for septic shock vs. SIRS in Blacks; p<0.003 by DeLong’s test; see Supplementary Material, Section 1). Higher SeptiScores for septic shock cases in Black as opposed to White patients could relate to genetics or environment, or a mixture of both. However, a number of studies suggest that genetic differences do not influence sepsis mortality or hospital length of stay once socioeconomic factors, such as number of comorbidities and access to healthcare, are taken into account [33, 34]. Without further data, to include increased patient numbers and details on socioeconomic variables, it is not possible to definitively identify the factors contributing to the observed higher SeptiScores in Black vs. White patients with septic shock.

### Comorbidities and diseases

Risk factors for sepsis include certain comorbidities, for example diabetes [35–37], hypertension [37], chronic kidney disease [38, 39], obesity [29, 30], cancer [40], and cardiovascular disease [41]. Because SeptiCyte RAPID is based on measurement of host immune response, comorbidities that involve the immune system could affect assay performance. AUC values for subgroups based on comorbidity and disease generally clustered around 0.82-0.83. One of the lowest SeptiCyte AUC values observed in this analysis was for patients with hypertension (AUC 0.79), but this was not significantly different (p=0.59) from the AUC 0.83 observed for patients with no hypertension noted. (**Table 2**). Another low SeptiCyte AUC value was observed for patients with diabetic hyperglycemia (AUC 0.73) – a finding which would benefit from additional study in an expanded patient cohort. Despite the small sample sizes in some patient subgroups, these results suggest that certain patients suspected of sepsis need not be excluded from SeptiCyte RAPID testing due to the specific pre-existing comorbidities and diseases we were able to examine in our study cohort.

### Type and site of infection

The immune status of leukocytes varies greatly depending upon the body compartment from which they are derived [42] and type of infection being responded to [43]. For example, in humans an intravenous lipopolysaccharide (LPS) injection suppresses the *ex vivo* peripheral blood mononuclear cell (PMBC) response to LPS but primes that of alveolar macrophages [44]. Therefore, the primary site of infection in sepsis could be expected to influence the PBMC response and hence SeptiCyte RAPID results. To investigate this further, we compared SeptiCyte RAPID performance in patient subgroups stratified by the primary site of infection, including the categories lung, abdomen, central nervous system, urinary tract, blood (bacteremia), other identified site, and ‘site not identified’ (i.e. sepsis of unknown origin) (Figure 2). We observed no statistically significant differences in SeptiCyte RAPID performance across a broad range of identified infection sites. AUC did appear lower (0.72) for patients with sepsis of unknown origin, which may relate to uncertainty in the retrospective diagnosis of patients in this subgroup [6, 45].

### Therapeutic interventions: immunosuppressants

We did not observe any significant effect of immunosuppressant therapy on performance of SeptiCyte RAPID (Table 3). However, this comparison covered a broad range of immunosuppressants, so no firm conclusions on the effect of any individual immunosuppressant on SeptiCyte RAPID can be drawn without performing a larger study with sufficient patient numbers. Regarding the glucocorticoid class specifically, it is known that short-term oral prednisone (4 days, 30mg/day) produces a distinct blood gene expression signature in COPD patients and that neither PLAC8 nor PLA2G7 were differentially expressed when comparing pre-treatment patients to Day 4 of treatment [46]. Further, PLAC8 and PLA2G7 are not known to be glucocorticoid regulated genes [47]. This suggests that treatment with glucocorticoids does not affect peripheral blood gene expression of the two biomarkers of SeptiCyte RAPID, and therefore is not expected to affect SeptiCyte RAPID results.

### Interventional treatment: antibiotics

Although blood cultures are the ‘gold standard’ for confirming bacteremia, up to fifty percent of patients suspected of sepsis and admitted to the ICU have negative cultures [48], some of which could be caused by prior use of antibiotics [49].

Standard practice in patients suspected of sepsis is to take blood cultures prior to antibiotic administration to avoid the potential negative impact on growth of organisms in blood culture. In this study we show that use of prophylactic antibiotics within 1 day prior to a patient presenting to ICU does not affect SeptiCyte RAPID performance. Therefore, a clinician need not withhold antibiotics prior to taking a blood sample for SeptiCyte RAPID analysis. However, treatment with antibiotics outside of this timeframe would be expected to affect SeptiScores as patients recover in response to appropriate antibiotic treatment [31].

### Phenotypic analysis

We conducted phenotypic analyses along the lines described by Seymour et al. [3] on our sepsis patient cohort (N=176). We employed two different unsupervised clustering methods (PCA/HC, and k-means) using a selection of readily available clinical variables. We identified three sepsis subgroups by PCA / HC, and two sepsis subgroups by k-means clustering. Key driving variables separating the sepsis subgroups included WBC, platelets, MAP, glucose, platelets, age, temperature.

We also conducted the same type of analyses on the entire sepsis+SIRS dataset (N=419). Similar to what we observed in the first analyses, these same phenotypic variables also resolved three subgroups in the PCA / HC analysis and two subgroups in the k-means analysis (Supplementary Material). Separation between subgroups in either analysis appeared to be driven at least partly by clinical severity of the sepsis response. Interestingly, neither the infection source or type of infecting pathogen was a significant factor, except that viral infections appeared to be less severe and more highly associated with sepsis of pulmonary origin.

Individual clinical variables that were identified as driving the PCA separation were assessed for their ability to discriminate sepsis from SIRS. Figure 6 compared the performance of these variables versus SeptiCyte RAPID. The results suggest that, in our study at least, patients with sepsis cannot easily be distinguished from those with SIRS using any of the individual clinical variables. In contrast, in subgroups defined by these driving variables, SeptiCyte RAPID differentiated sepsis from SIRS with AUCs ranging from 0.71 – 0.86. This finding is consistent with a previous demonstration [16] showing that combinations of up to 14 clinical variables do not outperform SeptiCyte RAPID. These results suggest that sepsis diagnostic tools that rely on these clinical variables alone (e.g. qSOFA or early warning scores) will be limited in their capacity to differentiate sepsis and SIRS patients.

The clinical variables we have identified as “driving” the separations in this study can be broadly mapped onto the phenotypes defined by Seymour et al. [3]: α, (systolic BP, limited use of vasopressors); β, (SOFA, chronic illness, renal dysfunction, age); γ (RR, WBC, PCT, platelets inflammation and pulmonary dysfunction); and δ (systolic BP, lactate, shock). Further, Sinha et al. [8] have identified two subgroups in ARDS and septic shock termed “hypo-inflammatory” and “hyper-inflammatory”. Patients with hyper-inflammatory septic shock had higher rates of blood culture positivity, increased inflammatory markers and poor outcomes (increased 28-day mortality and lower ICU-free days). In our study, although not restricted to septic shock patients, we identified a sepsis patient subgroup using PCA / HC analysis (subgroup 3) that is more likely to contain a higher proportion of patients with septic shock, as indicated by higher SOFA, RR, lactate and use of vasopressors and lower MAP. With respect to sepsis/SIRS discrimination, with the possible exception of higher SeptiCyte RAPID performance in septic shock patients, we observed similar SeptiCyte RAPID performance across our patient phenotypes and key driving variables. An important practical result is that SeptiCyte RAPID performance appears relatively unaffected by phenotype, or by key clinical variables used to define the phenotypes (Tables 4, 5, 6).

There are several limitations to this study. A first limitation is that the patients in our study cohort consisted of only 419 patients admitted to ICU and tested on the first day of ICU admission. If the study had included patients who were tested at times outside the first day of ICU admission, the relative performance estimates and factor sensitivities might have been different. A second limitation is that the cohort size (N=419) was not large enough to include quantitative analyses of the effects of other comorbidities and diseases. A third limitation is that all the patients were adults ≥18 years of age. We cannot say whether any differences in SeptiCyte RAPID performance occur in adolescents, children, infants or neonates. A fourth limitation is that, with respect to race/ethnicity, the dataset was sufficient only to analyze the main classes of subjects (White, Black, Asian, Hispanic) from North American and European geographic regions, and did not include other minorities or patients from other geographic regions. Future studies will need to be undertaken to establish performance in other racial/ethnic groups within North American and European populations, as well as racial/ethnic groups more generally from other areas of the world. A fifth limitation is that the metadata in our study was not sufficiently comprehensive or granular to allow a stratification according to socioeconomic factors, which are known to correlate with sepsis comorbidities [37].

Clinical identification of sepsis, even retrospectively, is characterized by significant diagnostic uncertainty [6,50]. This motivates the development of objective sepsis diagnostic tests [51,52]. In this study we have shown that SeptiCyte RAPID provides consistent discrimination of sepsis vs. SIRS across many subgroups within a heterogeneous adult, ICU patient population suspected of sepsis. Further studies involving larger numbers of patients will be required to confirm the robustness of SeptiCyte RAPID in broader populations.

## 5. Conclusions

We conducted a stratification analysis of SeptiCyte RAPID data from the pivotal studies MARS, VENUS, NEPTUNE (N=419). SeptiCyte RAPID demonstrated consistent performance for discrimination of sepsis vs. SIRS, using AUC as the performance measure, across a heterogenous adult, critically ill patient population when stratified by key demographic, clinical, microbiological and interventional parameters. This helps to support a clinical utility claim for SeptiCyte RAPID. Stratification analysis with phenotypic variables may be useful for resolving sepsis patients into subgroups based on clinical severity.

## Supplementary Material

Supplementary material is available for this manuscript. References [20–23, 53–62] are cited in the Supplementary Material.

## Author Contributions

Conceptualization: R.R.M. III, J.P.B., K.N., T.D.Y., J.T.K., S.C., R.F.D., R.B.B.; Data curation: J.P.B., V.K.S., J.A.K., B.P.S., S.K., K.N., D.S., J.T.K., S.C., R.B.B.; Formal analysis: M.L., S.O., R.E.R., M.J.S., K.N., T.D.Y., D.S., S.C., R.F.D., R.B.B.; Funding acquisition: M.J.S.; Investigation: R.B., A.M.E., G.S.M., B.K.L., F.R.D., V.K.S., N.R.A., J.A.G., M.Y., G.P., E.G., J.P.P., M.A., J.A.K., T.v.d.P., M.J.S., P.M.C.K.K., J.L., E.B., S.K., J.T.K., S.C., X.W.M.; Methodology: M.L., S.O., R.E.R., K.N., T.D.Y., D.S., J.T.K., R.B.B., X.W.M.; Project administration: A.M.E., F.R.D., J.L., S.K., T.D.Y., S.C., R.B.B.; Resources: R.B., A.M.E., F.R.D., V.K.S., T.v.d.P., B.P.S., J.L., S.K., D.S., S.C.; Software: K.N., D.S., X.W.M.; Supervision: R.B., A.M.E., B.K.L., F.R.D., E.G., J.P.P., M.A., T.v.d.P., S.K., S.C., R.B.B.; Validation: R.B., M.L., S.O., R.E.R., N.R.A., G.P., B.P.S., E.B., K.N., S.C., R.B.B., X.W.M.; Visualization: R.B., K.N., T.D.Y., D.S.; Writing - original draft: K.N., T.D.Y., S.C., R.B.B.; Writing - review & editing: R.B., G.S.M., R.R.M. III, J.P.B., M.L., R.E.R., V.K.S., N.R.A., J.A.G., M.Y., E.G., J.P.P., M.A., J.A.K., T.v.d.P., M.J.S., B.P.S., P.M.C.K.K., E.B., S.K., K.N., T.D.Y., D.S., J.T.K., S.C., R.F.D., R.B.B. All authors have read and agreed to the published version of the manuscript.

## Funding

This work was funded by Immunexpress, Inc.

## Institutional Review Board Statement

Ethics approval for the MARS trial was given by the medical ethics committee of AMC, Amsterdam (approval # 10-056C, 16 June 2010). Ethics approvals for the VENUS trial were given by the relevant Institutional Review Boards as follows: Intermountain Medical Center/Latter Day Saints Hospital (approval # 1024931, 21 January 2014); Johns Hopkins Hospital (approval # IRB00087839, 28 January 2016); Rush University Medical Center (approval # 15111104-IRB01, 11 March 2016); Loyola University Medical Center (approval # 208291, 10 March 2016); Northwell Healthcare (approval #16-02-42-03, 1 April 2016). Ethics approvals for the NEPTUNE trial were given by the relevant Institutional Review Boards as follows: Emory University (approval # IRB00115400, 3 December 2019); Grady Memorial Hospital (approval # 00-115400, 14 January 2020); Rush University Medical Center (approval # 19101603-IRB01, 16 January 2020); University of Southern California Medical Center (approval # HS-19-0884-CR001, 10 February 2020). All methods used in this study were carried out in accordance with the relevant guidelines and regulations.

## Informed Consent Statement

All subjects, or their legally authorized representatives, gave informed consent for participation in this study.

## Data Availability Statement

The datasets used and/or analyzed during the current study are available from the corresponding authors upon reasonable request.

## Data Availability

The datasets used and/or analyzed during the current study are available from the corresponding author upon reasonable request.

## Acknowledgments

The authors thank the clinical study coordinators of the NEPTUNE study: Joyce D. Brown (Rush University), Liliacna Jara (University of Southern California), Leona Wells and Maya C. Whaley (Grady Memorial Hospital).

## Conflicts of Interest

K.N., T.D.Y., D.S., J.T.K., S.C., R.F.D. and R.B.B. declare they are present or past employees or shareholders of Immunexpress, Inc. X.W.M. declares that she is a present employee of Princeton Pharmatech, Inc. R.R.M. III discloses that, over the time period of relevance to this work he was paid (as a consultant) an honorarium by Immunexpress for conducting a critical review of a Clinical Evaluation Report of SeptiCyte technology, submitted by Immunexpress to the Therapeutic Goods Administration (TGA) of Australia. N.R.A. declares that he has received grants from the NIH and Department of Defense for non-related work, and that payments from these grants are made to his institution, the University of Colorado. N.R.A. also declares that he is a committee member of the Pfizer Paxlovid US Medical Advisory Committee formed to broadly discuss COVID-19 related therapeutic priorities and populations of interest. B.K.L. declares that he is a consultant for Karius Inc., is a member of the Scientific Advisory Board for Seegene Inc., and has received honoraria for speaking for Qiagen Inc. No other competing interests are declared.

## Notes

### Author Declarations

Ethics approval for the MARS trial was given by the medical ethics committee of AMC, Amsterdam (approval # 10-056C, 16 June 2010). Ethics approvals for the VENUS trial were given by the relevant Institutional Review Boards as follows: Intermountain Medical Center/Latter Day Saints Hospital (approval # 1024931, 21 January 2014); Johns Hopkins Hospital (approval # IRB00087839, 28 January 2016); Rush University Medical Center (approval # 15111104-IRB01, 11 March 2016); Loyola University Medical Center (approval # 208291, 10 March 2016); Northwell Healthcare (approval #16-02-42-03, 1 April 2016). Ethics approvals for the NEPTUNE trial were given by the relevant Institutional Review Boards as follows: Emory University (approval # IRB00115400, 3 December 2019); Grady Memorial Hospital (approval # 00-115400, 14 January 2020); Rush University Medical Center (approval # 19101603-IRB01, 16 January 2020); University of Southern California Medical Center (approval # HS-19-0884-CR001, 10 February 2020). All methods used in this study were carried out in accordance with the relevant guidelines and regulations. Informed Consent Statement: All subjects, or their legally authorized representatives, gave informed consent for participation in this study.

### Summary of Updates

Updated affiliation and email address for Franco R D Alessio (author 9). Name correction for Xue W Mei (author 26). Updated conflicts of interest statement. Updated author contributions statement. Reformatted all references and added DOIs where missing. References now numbered consecutively as they appear in manuscript.

## References

1. Liu V.X., Bhimarao M., Greene J.D., Manickam R.N., Martinez A., Schuler A., et al. The Presentation, Pace, and Profile of Infection and Sepsis Patients Hospitalized Through the Emergency Department: An Exploratory Analysis. Crit. Care Explor. 2021, 3, e0344. DOI: 10.1097/CCE.0000000000000344.

2. Lengquist M., Varadarajan A., Alestam S., Friberg H., Frigyesi A., Mellhammar L. Sepsis mimics among presumed sepsis patients at intensive care admission: a retrospective observational study. Infection. 2024, 52, 1041–1053. DOI: 10.1007/s15010-023-02158-w.

3. Seymour C.W., Kennedy J.N., Wang S., Chang C.H., Elliott C.F., Xu Z., et al. Derivation, Validation, and Potential Treatment Implications of Novel Clinical Phenotypes for Sepsis. JAMA. 2019, 321, 2003–2017. DOI: 10.1001/jama.2019.5791.

4 Fohner A.E., Greene J.D., Lawson B.L., Chen J.H., Kipnis P., Escobar G.J., et al. Assessing clinical heterogeneity in sepsis through treatment patterns and machine learning. J. Am. Med. Inform. Assoc. 2019, 26, 1466–1477. doi: 10.1093/jamia/ocz106.

5. Reddy K., Sinha P., O’Kane C.M., Gordon A.C., Calfee C.S., McAuley D.F. Subphenotypes in critical care: translation into clinical practice. Lancet Respir. Med. 2020, 8, 631–643. DOI: 10.1016/S2213-2600(20)30124-7.

6. Lopansri BK, Miller, R.R. III, Burke J.P., Levy M., Opal S., Rothman R.E., et al. Physician agreement on the diagnosis of sepsis in the intensive care unit: estimation of concordance and analysis of underlying factors in a multicenter cohort. J. Intensive Care. 2019, 7, 13. DOI: 10.1186/s40560-019-0368-2.

7. Bhavani S.V., Semler M., Qian E.T., Verhoef P.A., Robichaux C., Churpek M.M., Coopersmith CM. Development and validation of novel sepsis subphenotypes using trajectories of vital signs. Intensive Care Med. 2022, 48, 1582–1592. DOI: 10.1007/s00134-022-06890-z.

8. Sinha P., Kerchberger V.E., Willmore A., Chambers J., Zhuo H., Abbott J., et al. Identifying molecular phenotypes in sepsis: an analysis of two prospective observational cohorts and secondary analysis of two randomised controlled trials. Lancet Respir Med. 2023, 11, 965–974. DOI: 10.1016/S2213-2600(23)00237-0.

9. Davenport E.E., Burnham K.L., Radhakrishnan J., Humburg P., Hutton P., Mills T.C., et al. Genomic landscape of the individual host response and outcomes in sepsis: a prospective cohort study. Lancet Respir Medicine. 2016, 4, 259–271. DOI:10.1016/s2213-2600(16)00046-1.

10. Scicluna B.P., van Vught L.A., Zwinderman A.H., Wiewel M.A., Davenport E.E., Burnham K.L., et al. Classification of patients with sepsis according to blood genomic endotype: a prospective cohort study. Lancet Respir. Med. 2017, 5, 816–826. DOI:10.1016/s2213-2600(17)30294-1.

11. Sweeney T.E., Azad T.D., Donato M., Haynes W.A., Perumal T.M., Henao R., et al. Unsupervised Analysis of Transcriptomics in Bacterial Sepsis Across Multiple Datasets Reveals Three Robust Clusters. Crit. Care Med. 2018, 46, 915–925. DOI: 10.1097/CCM.0000000000003084

12. DeMerle K.M., Angus D.C., Baillie J.K., Brant E., Calfee C.S., Carcillo J., et al. Sepsis Subclasses: A Framework for Development and Interpretation. Crit. Care Med. 2021, 49, 748–759. DOI: 10.1097/CCM.0000000000004842.

13. Coburn B., Morris A.M., Tomlinson G., Detsky A.S. Does this adult patient with suspected bacteremia require blood cultures? JAMA. 2012, 308, 502–511. DOI: 10.1001/jama.2012.8262. Erratum in: JAMA. 2013, 309, 343. DOI:10.1001/jama.2012.210984.

14. Vincent J.L., Bogossian E., Menozzi M. The Future of Biomarkers. Crit. Care Clin. 2020, 36, 177–187. DOI: 10.1016/j.ccc.2019.08.014.

15. Miller R.R. III, Lopansri B.K., Burke J.P., Levy M., Opal S., Rothman R.E., et al. Validation of a Host Response Assay, SeptiCyte LAB, for Discriminating Sepsis from Systemic Inflammatory Response Syndrome in the ICU. Am. J. Respir. Crit. Care Med. 2018, 198, 903–913. DOI: 10.1164/rccm.201712-2472OC. Erratum in: Am. J. Respir. Crit. Care Med. 2020, 202, 155. DOI: 10.1164/rccm.v202erratum2.

16. Balk R., Esper A.M., Martin G.S., Miller R.R. III, Lopansri B.K., Burke J.P., et al. Validation of SeptiCyte RAPID to Discriminate Sepsis from Non-Infectious Systemic Inflammation. J. Clin. Med. 2024, 13, 1194. DOI: 10.3390/jcm13051194

17. McHugh L., Seldon T.A., Brandon R.A., Kirk J.T., Rapisarda A., Sutherland A.J., et al. A Molecular Host Response Assay to Discriminate Between Sepsis and Infection-Negative Systemic Inflammation in Critically Ill Patients: Discovery and Validation in Independent Cohorts. PLoS Med. 2015, 12, e1001916. DOI: 10.1371/journal.pmed.1001916.

18. R Core Team. R: A language and environment for statistical computing. R Foundation for Statistical Computing, Vienna, Austria (2024). ISBN 3-900051-07-0. http://www.R-project.org/ (accessed September 1, 2024).

19. Robin X., Turck N., Hainard A., Tiberti N., Lisacek F., Sanchez J., Müller M. pROC: an open-source package for R and S+ to analyze and compare ROC curves. BMC Bioinformatics. 2011, 12,77. DOI: 10.1186/1471-2105-12-77.

20. Hanley J.A., McNeil B.J. The meaning and use of the area under a receiver operating characteristic (ROC) curve. Radiology. 1982, 143, 29–36. DOI: 10.1148/radiology.143.1.7063747.

21. DeLong E.R., DeLong D.M., Clarke-Pearson D.L. Comparing the areas under two or more correlated receiver operating characteristic curves: a nonparametric approach. Biometrics. 1988, 44, 837–845. DOI: 10.2307/2531595.

22. Skalsk’a H., Freylich V. (2006) Web-Bootstrap Estimate of Area Under ROC Curve. Austrian J Stat. 2006, *35*, 325–330. DOI: 10.17713/ajs.v35i2&3.379.

23. Mason S.J., Graham N.E. Areas beneath the relative operating characteristics (ROC) and relative operating levels (ROL) curves: Statistical significance and interpretation. Q. J. R. Meteorol. Soc. 2002, 128, 2145–2166. doi:10.1256/003590002320603584.

24. Xu, W., Dai, J., Hung, Y. S., & Wang, Q. Estimating the area under a receiver operating characteristic (ROC) curve: Parametric and nonparametric ways. Signal Process. 2013, 93, 3111– 3123. DOI:10.1016/j.sigpro.2013.05.010

25. Lê S., Josse J., Husson F. (2008). “FactoMineR: A Package for Multivariate Analysis.” J. Stat. Softw. 2008, *25*, 1–18. DOI:10.18637/jss.v025.i01.

26. Yang C., Yang K., Zhou B. (2015) A hierarchical Clustering Method Based on PCA-Clusters Merging for Quasi-linear SVM. Proceedings of the 2015 International Conference on Automation, Mechanical Control and Computational Engineering (AMCCE 2015). DOI: 10.2991/amcce-15.2015.407.

27. Maugeri, A., Barchitta, M., Basile, G. et al. Applying a hierarchical clustering on principal components approach to identify different patterns of the SARS-CoV-2 epidemic across Italian regions. Sci Rep. 2021, 11, 7082. DOI: 10.1038/s41598-021-86703-3.

28. Koh K.-Y., Ahmad S., Lee J-I., Suh G.-H., Lee C.-M. Hierarchical Clustering on Principal Components Analysis to Detect Clusters of Highly Pathogenic Avian Influenza Subtype H5N6 Epidemic across South Korean Poultry Farms. Symmetry. 2022; 14, 598. DOI: 10.3390/sym14030598.

29. Hu J., Gan Q., Zhou D., Xia X., Xiang W., Xiao R., Tang J., Li J. Evaluating the risk of sepsis attributing to obesity: a two-sample Mendelian randomization study. Postgrad. Med. J. 2023, 99, 1266–1271. DOI: 10.1093/postmj/qgad072.

30. Wang J., Hu Y., Zeng J., Li Q., He L., Hao W., Song X., Yan S., Lv C. Exploring the Causality Between Body Mass Index and Sepsis: A Two-Sample Mendelian Randomization Study. Int. J. Public Health. 2023, 68, 1605548. DOI: 10.3389/ijph.2023.1605548.

31. Peri A.M., Rafiei N., O’Callaghan K., Brischetto A., Graves B., Sinclair H., et al. Host response signature trends in persistent bacteraemia and metastatic infection due to Staphylococcus aureus and Gram-negative bacilli: a prospective multicentre observational study. Infect. Dis. (Lond). 2024, 56, 268–276. doi: 10.1080/23744235.2023.2294122.

32. Papathanakos G., Andrianopoulos I., Xenikakis M., Papathanasiou A., Koulenti D., Blot S., et al. Clinical Sepsis Phenotypes in Critically Ill Patients. Microorganisms. 2023, 11, 2165. DOI: 10.3390/microorganisms11092165.

33. DiMeglio M., Dubensky J., Schadt S., Potdar R., Laudanski K. Factors Underlying Racial Disparities in Sepsis Management. Healthcare (Basel). 2018, 6, 133. DOI: 10.3390/healthcare6040133.

34. Vazquez Guillamet M.C., Dodda S., Liu L., Kollef M.H., Micek S.T. Race Does Not Impact Sepsis Outcomes When Considering Socioeconomic Factors in Multilevel Modeling. Crit. Care Med. 2022, 50, 410–417. DOI: 10.1097/CCM.0000000000005217.

35. Frydrych L.M., Fattahi F., He K., Ward P.A., Delano M.J. Diabetes and Sepsis: Risk, Recurrence, and Ruination. Front. Endocrinol (Lausanne). 2017, 8, 271. DOI: 10.3389/fendo.2017.00271.

36. Carey I.M., Critchley J.A., DeWilde S., Harris T., Hosking F.J., Cook D.G. Risk of Infection in Type 1 and Type 2 Diabetes Compared With the General Population: A Matched Cohort Study. Diabetes Care. 2018, 41, 513–521. DOI: 10.2337/dc17-2131.

37. Ahlberg C.D., Wallam S., Tirba .LA., Itumba S.N., Gorman L., Galiatsatos P. Linking Sepsis with chronic arterial hypertension, diabetes mellitus, and socioeconomic factors in the United States: A scoping review. J. Crit. Care. 2023, 77, 154324. DOI: 10.1016/j.jcrc.2023.154324.

38. Doi K., Leelahavanichkul A., Hu X., Sidransky K.L., Zhou H., Qin Y., Eisner C., Schnermann J, Yuen PS, Star RA. Pre-existing renal disease promotes sepsis-induced acute kidney injury and worsens outcome. Kidney Int. 2008, 74, 1017–1025. DOI: 10.1038/ki.2008.346.

39. Ou S.M., Lee K.H., Tsai M.T., Tseng W.C., Chu Y.C., Tarng D.C. Sepsis and the Risks of Long-Term Renal Adverse Outcomes in Patients With Chronic Kidney Disease. Front. Med. (Lausanne). 2022, 9, 809292. DOI: 10.3389/fmed.2022.809292.

40. Gudiol C., Albasanz-Puig A., Cuervo G., Carratalà J. Understanding and Managing Sepsis in Patients With Cancer in the Era of Antimicrobial Resistance. Front Med. 2021, 8, 636547. DOI: 10.3389/fmed.2021.636547.

41. Nakada T.A., Takahashi W., Nakada E., Shimada T., Russell J.A., Walley K.R. Genetic Polymorphisms in Sepsis and Cardiovascular Disease: Do Similar Risk Genes Suggest Similar Drug Targets? Chest. 2019, 155, 1260–1271. DOI: 10.1016/j.chest.2019.01.003.

42. Cavaillon J.-M., Chousterman B.G., Skirecki T. Compartmentalization of the inflammatory response during bacterial sepsis and severe COVID-19. J. Intensive Med. 2024, 4, 326–340. DOI: 10.1016/j.jointm.2024.01.001.

43. Nguyen Q.-T., Nguyen T.-H.T., Ju S.-A., Lee Y.-S., Han S.H., Lee S.-C., et al. CD137 Expressed on Neutrophils Plays Dual Roles in Antibacterial Responses against Gram-Positive and Gram-Negative Bacterial Infections. Infect Immun. 2013, 81, 2168–2177. DOI: 10.1128/IAI.00115-13.

44. Cavaillon J.-M., Chousterman B.G., Skirecki T. Compartmentalization of the inflammatory response during bacterial sepsis and severe COVID-19. J. Intensiv. Med. 2024, 4, 326–340. DOI: 10.1016/j.jointm.2024.01.001.

45. Roger P.-M., Keïta-Perse O., Mainardi J.-L. Diagnostic uncertainty in infectious diseases: advocacy for a nosological framework. Infect. Dis. Now. 2023, 53, 104751. DOI: 10.1016/j.idnow.2023.104751.

46. Takiguchi H., Chen V., Obeidat M., Hollander Z., FitzGerald J.M., McManus B.M., Ng R.T., Sin D.D. Effect of short-term oral prednisone therapy on blood gene expression: a randomised controlled clinical trial. Respir. Res. 2019, 20, 176. DOI: 10.1186/s12931-019-1147-2.

47. Chinenov Y, Coppo M, Gupte R, Sacta MA, Rogatsky I. Glucocorticoid receptor coordinates transcription factor-dominated regulatory network in macrophages. BMC Genomics. 2014, 15, 656. DOI: 10.1186/1471-2164-15-656.

48. Gupta S., Sakhuja A., Kumar G., McGrath E., Nanchal R.S., Kashani K.B. Culture-Negative Severe Sepsis: Nationwide Trends and Outcomes. Chest 2016, 150, 1251–1259. DOI: 10.1016/j.chest.2016.08.1460

49. Hirosawa T., Sakamoto T., Hanai S., Harada Y., Shimizu T. Effect of Prior Antibiotic Treatment on Blood Culture in an Outpatient Department of General Internal Medicine: A Retrospective Case-Control Analysis. Int. J. Gen. Med. 2023, 16, 2709–2717. DOI: 10.2147/IJGM.S416235.

50. Rhee C., Kadri S.S., Danner R.L., Suffredini A.F., Massaro A.F., Kitch B.T., et al. Diagnosing sepsis is subjective and highly variable: a survey of intensivists using case vignettes. Crit Care. 2016, 20, 89. DOI: 10.1186/s13054-016-1266-9.

51. Opal S.M., Wittebole X. Biomarkers of Infection and Sepsis. Crit. Care Clin. 2020, 36, 11–22. DOI: 10.1016/j.ccc.2019.08.002.

52. Pierrakos C., Velissaris D., Bisdorff M., Marshall J.C., Vincent J.-L. Biomarkers of sepsis: Time for a Reappraisal. Crit Care. 2020, 24, 287. 10.1186/s13054-020-02993-5.

53. Cohen, J. A coefficient of agreement for nominal scales. Educ. Psychol. Meas.1960, 20, 213–220. DOI: 10.1177/001316446002000104.

54. Fleiss, J.L., Cohen, J., and Everitt, B.S. Large sample standard errors of kappa and weighted kappa. PsychoL. Bull. 1969, 72, 323–327. DOI:10.1037/h0028106.

55. Goldstein-Greenwood, J. (UVA Library StatLab). ROC Curves and AUC for Models Used for Binary Classification. 2022, *Available online:* https://library.virginia.edu/data/articles/roc-curves-and-auc-for-models-used-for-binary-classification (accessed August 19, 2024).

56. Kuye I., Anand V., Klompas M., Chan C., Kadri S.S., Rhee C. Prevalence and Clinical Characteristics of Patients With Sepsis Discharge Diagnosis Codes and Short Lengths of Stay in U.S. Hospitals. Critical Care Explor. 2021, 3, e0373. DOI: 10.1097/CCE.0000000000000373.

57. Qin C., Zhang S., Zhao Y., Ding X., Yang F., Zhao Y. Diagnostic value of metagenomic next-generation sequencing in sepsis and bloodstream infection. Front. Cell. Infect. Microbiol. 2023, 13, 1117987. DOI: 10.3389/fcimb.2023.1117987.

58. Shoonjans, F. MedCalc manual: Easy-to-use statistical software. 2017. MedCalc Software bvba, Ostend, Belgium. ISBN 978-1520321578.

59. Xu, W., Dai, J., Hung, Y. S., & Wang, Q. Estimating the area under a receiver operating characteristic (ROC) curve: Parametric and nonparametric ways. Signal Process. 2013, 93, 3111–3123. DOI:10.1016/j.sigpro.2013.05.010.

60. Yang L., Lin Y., Wang J., Song J., Wei B., Zhang X., et al. Comparison of Clinical Characteristics and Outcomes Between Positive and Negative Blood Culture Septic Patients: A Retrospective Cohort Study. Infect Drug Resist. 2021, 14, 4191–205. DOI: 10.2147/IDR.S334161.

61. Zhou X.H., Obuchowski N.A., McClish D.K. Statistical methods in diagnostic medicine, 2^nd^ Edition. John Wiley and Sons, Hoboken, New Jersey, USA. 2011. ISBN: 978-0-470-18314-4.

62. Zweig M.H., Campbell G. Receiver-operating characteristic (ROC) plots: a fundamental evaluation tool in clinical medicine. Clin. Chem. 1993, 39, 561–577. DOI: 10.1093/clinchem/39.4.561. Erratum in: Clin. Chem. 1993, 39, 1589. DOI: 10.1093/clinchem/39.8.1589.

