## Supplementary Material for "Rapid and robust identification of sepsis using SeptiCyte RAPID in a heterogenous patient population"

**Table of Contents**

Section 1 - Race / Ethnicity

Section 2 - Infection Source and Type

Section 3 - Pharmaceuticals

Section 4 - Potential Interferents

Section 5 - Phenotypic Stratification

Section 6 – Notes on Statistical Analysis of Small Strata

Section 7 - References

### 1. Race / Ethnicity – Further Examination

Following from Table 1 in the main text, we conducted a further stratification of the sepsis group according to race/ethnicity, and then searched for differences in SeptiCyte RAPID performance. The only significant finding ($p<0.003$) was a higher AUC value for septic shock vs. SIRS in Black patients (AUC 0.96) as opposed to White patients (AUC 0.83). Upon closer examination, this could be attributed to higher SeptiScores for Black vs. White septic shock patients (ANOVA, p-value: 0.072), as the SIRS distributions for Black vs. White subgroups were indistinguishable (**Figure S1**). When evaluating the clinical presentation between White & Black septic shock patients, several parameters were statistically significantly different between the two populations. As opposed to Black septic shock patients, the White patients had elevated values for minimum respiratory rate (RR.min) (ANOVA, p-value: 0.03). As opposed to White septic shock patients, the Black patients had elevated glucose (max) values (ANOVA, p-value: 0.02). Finally, a greater proportion of maximum mean arterial pressure (MAP.max) values for Black patients fell at either <70 or >100 mm Hg when compared to the White septic shock patients (ANOVA, p-value: 0.02).

***Supplement Figure S1:*** *SeptiScores after stratification of the White and Black patient subgroups with respect to sepsis severity.* SIRS patients are shown for reference. The boxplot shows the mean of the data as a horizontal line. The median is indicated by a dot either above or below the mean line. Note the difference between AUC = 0.83 for septic shock (White) vs. SIRS (White), compared to AUC = 0.96 for septic shock (Black) vs. SIRS (Black). The two ROC values differed significantly at the $p<0.003$ level (comparison method: bootstrap).

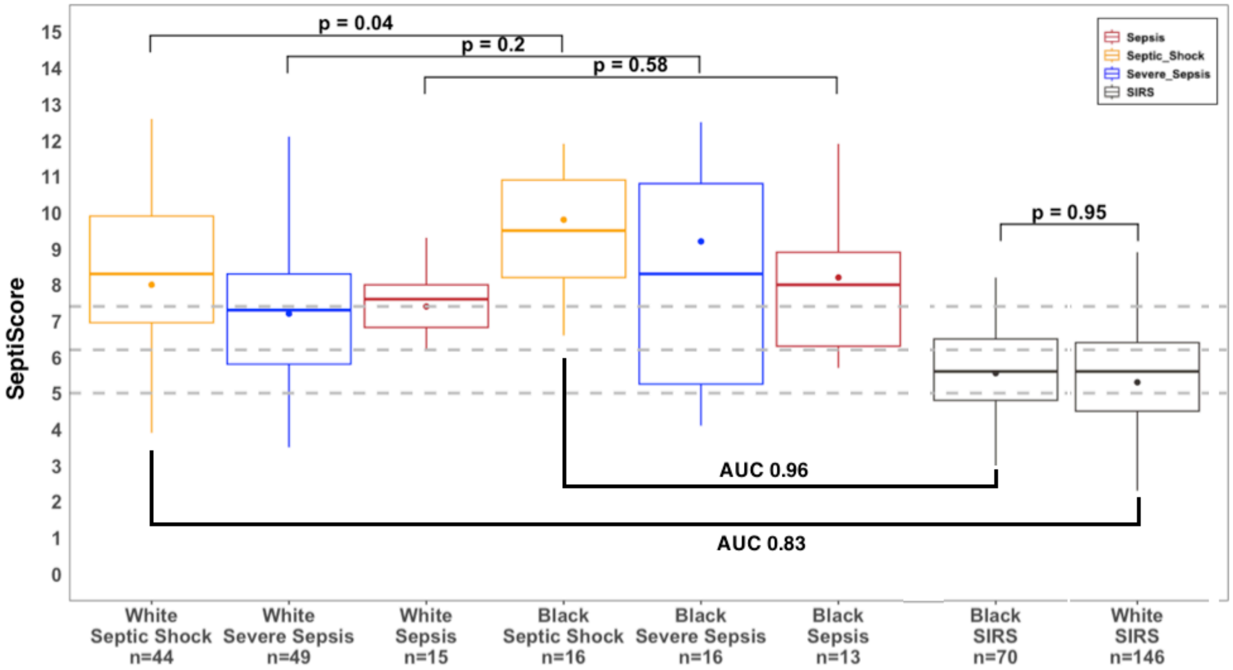

No significant difference was found when the Black sepsis+SIRS group was split by recent geographic origin (Europe: AUC 0.84 ± 0.11 vs. USA: AUC 0.85 ± 0.04). Similarly, no significant difference was found when the White sepsis+SIRS group was split by recent geographic origin (Europe: AUC 0.78 ± 0.05 vs. USA: AUC 0.82 ± 0.04). The discrimination of sepsis vs. SIRS was not significantly confounded by age for either the Black, White or Asian subgroups ($p>0.05$), although it was for the Hispanic subgroup ($p<0.03$) in which the SIRS patients (39±14 years) were significantly younger than the sepsis patients (average 57±10 years). The discrimination of sepsis vs. SIRS was not confounded by sex, for either the Black subgroup ($p=0.74$), the White subgroup ($p=0.70$) or the cohort as a whole ($p=0.62$).

### <u>2. Infection Type and Source</u>

We previously demonstrated that SeptiCyte RAPID performance was not affected by the type of sepsis pathogen defined by Gram staining (Section 9 of the Supplement to Balk et al., 2024). We have now extended this analysis to determine if different sepsis-causing pathogens were associated with different sites of infection. The study-specific Electronic Data Capture database was examined for each patient who was classified as septic by Forced RPD. For each clinically adjudicated site of infection, all positive culture and molecular test results identifying the infecting pathogen were compiled. If multiple pathogens were identified in a single patient, then all were taken into account. Results are summarized in **Tables S1** and **S2.**

**Table S1** (overview) shows that viral pathogens were more often detected in patients with pulmonary sepsis. Of 26 sepsis patients with a viral pathogen detected, 21 (81%) had pulmonary sepsis. A viral pathogen infecting the central nervous system (CNS) was detected in 3 septic patients, but not in any patients with urosepsis. Such results, besides having a biological basis, may well be influenced by current viral testing practices: patients with respiratory or CNS clinical signs would more likely be tested using a multi-analyte nucleic amplification panel test (i.e. "syndromic panel") compared to patients with clinical signs of a urinary tract infection. In urosepsis, Gram negative pathogens were more commonly isolated as compared to Gram positive pathogens.

**Table S2** (detailed breakdown) provides the following observations. (1) *Staphylococcus aureus* was the most frequently detected pathogen (19% of all pathogen detection events) and displayed the strongest association with pulmonary and blood sources of septic infection. (2) *Escherichia coli* was the second most frequently detected pathogen (15.2% of detection events) and was most strongly associated with urosepsis. (3) *Pseudomonas aeruginosa* was the third most frequently detected pathogen (8.2% of detection events) and was most strongly associated with blood and urinary sources of septic infection. Such observations are similar to those previously reported for blood cultures and next generation sequencing results in intensive care patients and across hospitals (Qin et al., 2023, Kuye et al., 2021, Yang et al., 2021).

***Supplement Table S1 (overview):*** *Sepsis patients stratified by source of infection according to 1) the initial impression by the attending physician, and 2) the class of pathogen eventually identified by culture or molecular test.* This table presents an overview of aggregated data. Note the low number of patients with central nervous system (CNS) identified as infection source.

| | **Source of Infection (Initial Impression)** | | | | | | |
|---|---|---|---|---|---|---|---|
| **pathogen** | **All sources** | **Pulmonary** | **Abdominal** | **Blood** | **Urinary** | **CNS** | **Other** |
| Gram Negative bacteria | 85 | 20 | 17 | 8 | 26 | 2 | 12 |
| Gram Positive bacteria | 75 | 23 | 17 | 10 | 12 | 1 | 12 |
| Viral | 26 | 21 | 1 | 1 | 0 | 3 | 1 |
| Fungal | 11 | 3 | 4 | 0 | 3 | 0 | 1 |
| **Bacterial genera or species** | | | | | | | |
| Staphylococcus aureus (G+) | 30 (18.8%) | 10 (23.3%) | 2 (5.9%) | 8 (44.4%) | 3 (7.9%) | 1 (33.3%) | 6 (25.0%) |
| Escherichia coli (G-) | 26 (16.2%) | 4 (9.3%) | 5 (14.7%) | 1 (5.6%)* | 10 (26.3%) | 1 (33.3%) | 5 (20.8%) |
| Pseudomonas aeruginosa (G-) | 13 (8.1%) | 3 (7.0%) | 1 (2.9%)* | 3 (16.7%) | 5 (13.2%) | 0 (0.0%) | 1 (4.2%)* |
| Enterococcus spp. (G+) | 15 (9.4%) | 3 (7.0%) | 5 (14.7%) | 1 (5.6%) | 5 (13.2%) | 0 (0.0%) | 1 (4.2%) |
| Streptococcus spp. (G+) | 11 (6.9%) | 5 (11.6%) | 4 (11.8%) | 0 (0.0%) | 0 (0.0%) | 0 (0.0%) | 2 (8.3%) |
| Klebsiella spp. (G-) | 10 (6.2%) | 3 (7.0%) | 4 (11.8%) | 0 (0.0%) | 2 (5.3%) | 0 (0.0%) | 1 (4.2%) |
| Other bacteria (G+) | 19 (11.9%) | 5 (11.6%) | 6 (17.6%) | 1 (5.6%) | 4 (10.5%) | 0 (0.0%) | 3 (12.5%) |
| Other bacteria (G-) | 36 (22.5%) | 10 (23.3%) | 7 (20.6%) | 4 (22.2%) | 9 (23.7%) | 1 (33.4%) | 5 (20.8%) |
| Total bacteria | 160 (100.0%) | 43 (100.0%) | 34 (100.0%) | 18 (100.0%) | 38 (100.0%) | 3 (100.0%) | 24 (100.0%) |

***Supplement Table S2 (detail):*** *Sepsis patients stratified by source of infection according to 1) the initial impression by the attending physician, and 2) the class of pathogen eventually identified by culture or molecular test.* This table presents a detailed breakdown by pathogen type. Abbreviation: CNS, central nervous system.

| | **Source of Infection (Initial Impression)** | | | | | | |
|---|---|---|---|---|---|---|---|
| **pathogen** | **All sources** | **Pulmonary (N=59)** | **Abdominal (N=30)** | **Blood (N=17)** | **Urinary (N=24)** | **CNS (N=6)** | **Other (N=26)** |
| **VIRUSES** | | | | | | | |
| Influenza (virus) | 6 | 0 | 0 | 1 | 0 | 0 | 0 |
| Influenza A | | 3 | 0 | 0 | 0 | 0 | 0 |
| Influenza B | | 0 | 0 | 0 | 0 | 0 | 1 |
| Coronavirus spp. | 5 | 0 | 0 | 0 | 0 | 1 | 0 |
| SARS-CoV-2 | | 3 | 1 | 0 | 0 | 0 | 0 |
| Epstein Barr virus | 1 | 0 | 0 | 0 | 0 | 1 | 0 |
| Herpes simplex virus | 1 | 1 | 0 | 0 | 0 | 1 | 0 |
| Metapneumovirus | 2 | 3 | 0 | 0 | 0 | 0 | 0 |
| Rhinovirus | 8 | 8 | 0 | 0 | 0 | 0 | 0 |
| Respiratory Syncytial virus | 3 | 3 | 0 | 0 | 0 | 0 | 0 |
| **FUNGI** | | | | | | | |
| Candida spp. | 9 | 2 | 4 | 0 | 3 | 0 | 0 |
| Aspergillus fumigatus | 2 | 1 | 0 | 0 | 0 | 0 | 1 |
| **GRAM NEGATIVE** | | | | | | | |
| Escherichia coli | 24 | 4 | 3 | 1 | 10 | 1 | 5 |
| Pseudomonas aeruginosa | 13 | 3 | 1 | 3 | 5 | 0 | 1 |
| Klebsiella spp. | 10 | 0 | 1 | 0 | 2 | 0 | 0 |

| | Source of Infection (Initial Impression) | | | | | | |
|---|---|---|---|---|---|---|---|
| **pathogen** | **All sources** | **Pulmonary (N=59)** | **Abdominal (N=30)** | **Blood (N=17)** | **Urinary (N=24)** | **CNS (N=6)** | **Other (N=26)** |
| Klebsiella pneumoniae | | 3 | 3 | 0 | 0 | 0 | 1 |
| Enterobacter aerogenes | 8 | 0 | 1 | 0 | 0 | 0 | 0 |
| Enterobacter cloacae | | 1 | 1 | 0 | 1 | 0 | 1 |
| Enterobacter spp. | | 2 | 0 | 0 | 1 | 0 | 0 |
| Proteus vulgaris | 5 | 0 | 0 | 0 | 1 | 0 | 0 |
| Proteus mirabilis | | 0 | 2 | 0 | 1 | 0 | 1 |
| Haemophilus influenzae | 4 | 1 | 0 | 1 | 1 | 0 | 0 |
| Haemophilus segnis | | 1 | 0 | 0 | 0 | 0 | 0 |
| Acinitobacter spp. | 3 | 3 | 0 | 0 | 0 | 0 | 0 |
| Bacteroides fragilis | 3 | | 1 | 1 | 0 | 0 | 1 |
| Citrobacter freundii | 2 | 1 | 0 | 1 | 0 | 0 | 0 |
| Serratia marsecens | 2 | 0 | 1 | 1 | 0 | 0 | 0 |
| Providencia rettgeri | 2 | 0 | 0 | 0 | 1 | 0 | 0 |
| Providencia stuartii | | 0 | 0 | 0 | 0 | 0 | 1 |
| Alcaligenes faecalis | 1 | 0 | 0 | 0 | 1 | 0 | 0 |
| Prevotella Nigrescens | 1 | 0 | 1 | 0 | 0 | 0 | 0 |
| Stenotrophomonas maltophilia | 1 | 0 | 0 | 0 | 0 | 1 | 0 |
| Fusobacterium spp. | 1 | 0 | 0 | 0 | 1 | 0 | 0 |
| Moraxella catarrhalis | 1 | 1 | 0 | 0 | 0 | 0 | 0 |
| Salmonella enteritidis | 1 | 0 | 1 | 0 | 0 | 0 | 0 |
| Pasturella multocida | 1 | 0 | 0 | 0 | 0 | 0 | 1 |
| Gram negative bacilli | 2 | 0 | 1 | 0 | 1 | 0 | 0 |
| **GRAM POSITIVE** | 0 | 0 | 0 | 0 | 0 | 0 | 0 |

| | Source of Infection (Initial Impression) | | | | | | |
|---|---|---|---|---|---|---|---|
| **pathogen** | **All sources** | **Pulmonary (N=59)** | **Abdominal (N=30)** | **Blood (N=17)** | **Urinary (N=24)** | **CNS (N=6)** | **Other (N=26)** |
| Staphylococcus aureus | 30 | 10 | 2 | 8 | 3 | 1 | 6 |
| Enterococcus faecalis | 15 | 2 | 2 | 1 | 3 | 0 | 0 |
| Enterococcus faecium | | 0 | 2 | 0 | 2 | 0 | 0 |
| Enterococcus spp. | | 1 | 1 | 0 | 0 | 0 | 1 |
| Streptococcus pneumoniae | 11 | 3 | 0 | 0 | 0 | 0 | 0 |
| Streptococcus agalactiae | | 0 | 1 | 0 | 0 | 0 | 0 |
| Streptococcus milleri group | | 0 | 1 | 0 | 0 | 0 | 0 |
| Streptococcus spp. | | 1 | 2 | 0 | 0 | 0 | 2 |
| Streptococcus beta hemolytic group C | | 1 | 0 | 0 | 0 | 0 | 0 |
| Staphylococcus epidermidis | 8 | 2 | 2 | 0 | 0 | 0 | 2 |
| Staphylococcus hominis | | 0 | 0 | 0 | 1 | 0 | 0 |
| Staphylococcus spp. | | 0 | 0 | 0 | 1 | 0 | 0 |
| Gram positive cocci | 1 | 0 | 1 | 0 | 0 | 0 | 0 |
| Clostridium difficile | 3 | 0 | 2 | 0 | 0 | 0 | 0 |
| Clostridium perfringens | | 0 | 1 | 0 | 0 | 0 | 0 |
| Corynebacterium spp. | 3 | 2 | 0 | 1 | 0 | 0 | 0 |
| Proprionibacterium spp. | 3 | 1 | 0 | 0 | 1 | 0 | 1 |
| Winkia neuii | 1 | 0 | 0 | 0 | 1 | 0 | 0 |

**3. Pharmaceuticals**

A list of >400 drugs was assembled by compiling entries from the study-specific Electronic Data Capture database for each patient. Relevant drugs fell into the following categories: immunosuppressants, anti-neoplastics, antibiotics, inotropes, vasopressors.

The following immunosuppressants were specified in the medical record: corticosteroids (fluticasone, betamethasone, triamcinolonacetonide hydrofiele, hydrocortisone, beclomethasone, budesonide, ciclesonide), ciclosporine (cyclosporin), azathioprine, Non-Steroidal Anti-Inflammatory Drugs (NSAIDs), and other anti-inflammatory medications with known immunosuppressive effects (carbasalate calcium, paracetamol, diclofenac, colchicine, spironolactone, digoxin, desogestrel, and valproate).

The following anti-neoplastics were specified in the medical record (data available only from MARS trial): Esketamine, Levetiracetam, Buprenorfine, Bupivacaine, Pravastatine, Emtricitabine.

The following antibiotics were specified in the medical record: Acyclovir, Amikacin, Amoxicillin, Amoxicillin/clavulanic acid (Augmentin), Ampicillin, Ampicillin+sulbactam (UNASYN), Anidulafungin, Atovaquone (mepron), Azithromycin, Aztreonam, Bacitracin, Bactrim (Trimethoprim/Sulfamethoxazole or Trimoxazole), Benzylpenicillin (Benzathine), Cefamandole, Cefazolin, Cefepime, Cefotaxime, Cefoxitin (Mefoxin), Ceftazidime, Ceftriaxone, Cefuroxime, Cephalexin, Cefazolin, Ciprofloxacin, Clarithromycin, Clindamycin, Colistin, Cotrimoxazole, Dapsone, Daptomycin, Doxycycline (Vibramycin), Ertapenem, Erythromycin, Flucloxacillin, Fluconazole, Ganciclovir, Gentamycin, Isoniazid, Keflex, Levofloxacin, Linezolid (Zyvox), Meropenem, Metronidazole, Micafungin, Moxifloxacin, Nystatin, Oseltamivir, Oxacillin, PenicillinG/Benzathine, Piperacillin, Piperacillin/Tazobactam, Primaxin, Rifampin, Rifaximin, Ritonovir, Tazobactam, Tobramycin, Valacyclovir, Vancomycin, Voriconazole, Zovirax.

The following inotropes were specified in the medical record: vasopressin, phenylephrine, epinephrine, norepinephrine, dobutamine. Note that there is some overlap with vasopressors.

The following vasopressors were specified in the medical record: dopamine, dobutamine, milrinone, digoxin, verapamil, clonidine, epinephrine, norepinephrine (levophed), vasopressin, phenylepinephrine, terlipressin. Note that there is some overlap with inotropes.

Table 3 and Figures 3 & 4 in the main text describe the results of this analysis of pharmaceutical agents. No significant effect of immunosuppressants, antibiotics (administered over -1 to +1 day of ICU admission) or inotropes/vasopressors was observed on SeptiCyte RAPID performance for discriminating sepsis from SIRS.

**4. Potential Interferents**

We tested analytically a set of 9 endogenous and 8 exogenous potential interferents which might be present in blood samples from critically ill patients (**Table S3**). The endogenous compounds were expected to be present in blood samples either under normal conditions (hemoglobin, albumin), in certain chronic medical conditions (bilirubin, triglycerides, rheumatoid factor) or elevated in response to inflammation (LBP, CRP, IL-6, s-CD14). The exogenous compounds were used to treat infections (imipenem, cephotaxime, vancomycin), or other chronic or acute medical conditions (heparin, dobutamine, furosemide, noradrenaline, dopamine).

Results are presented in **Table S4**. The following acceptance criterion was used for the analytical interference study: mean |SeptiScore difference| < 1.5 units between replicates of samples with interferent and their respective solvent controls. Results in this table demonstrate that this criterion has been met, i.e. that none of the tested compounds interfered with SeptiCyte RAPID at the tested concentrations.

***Supplement Table S3****: Compounds tested analytically for interference with SeptiCyte RAPID.*

| **Compound** | **Type** | **Function or medical use** | **Tested concentration** |
|---|---|---|---|
| Albumin | endogenous | Abundant plasma protein | 5 g/dL |
| Bilirubin | endogenous | Component of the catabolic pathway that breaks down heme | 20 mg/dL |
| CD14, soluble (s-CD14) | endogenous | Key intermediate in the host response pathway to Gram (-) bacterial infection. CD14 binds to the LPS-LBP complex, and then delivers LPS to TLR-4 (Ryu et al., 2017) | 5 µg/mL |
| Cefotaxime | exogenous | Cephalosporin (Medical use: antibiotic) | 670 µmol/L |
| C-reactive protein (CRP) | endogenous | Acute phase protein, induced in response to inflammation | 4 mg/dL |
| Dobutamine | exogenous | Beta1-adrenergic agonist, functions as an inotrope (Medical use: to treat low blood pressure or cardiogenic shock. Used in cases of septic shock.) | 12 µg/mL |
| Dopamine | exogenous | A positive inotrope (Medical use: increases the systolic blood pressure and heart rate. Used in cases of septic shock.) | 6.0 µmol/L |
| Furosemide | exogenous | Loop diuretic (Medical use: to treat edema) | 180 µmol/L |
| Hemoglobin | endogenous | Oxygen carrier in red blood cells | 20 g/dL |
| Heparin | exogenous | Anticoagulant (Medical use: to decrease blood clotting) | 3000 U/L |
| Imipenem | exogenous | Carbapenem (Medical use: antibiotic) | 1.2 mg/mL |
| Interleukin-6 (IL-6) | endogenous | Cytokine (produced at site of inflammation) | 15 pg/mL |
| Lipopolysaccharide binding protein (LBP) | endogenous | Acute phase protein that binds to lipopolysaccharide (LPS) from Gram (-) bacteria. Enhances the host response to LPS, by binding to CD14 (Meng et al., 2021) | 45 µg/mL |
| Noradrenaline | exogenous | Nonselective adrenergic agonist, functions as a vasopressor (Medical use: to treat low blood pressure. Used in cases of septic shock.) | 700 pg/mL |
| Rheumatoid factor | endogenous | Auto-antibody against the Fc portion of IgG | 45 IU/mL |
| Triglycerides | endogenous | Main constituent of body fat in humans. High levels in blood associated with heart disease | 500 mg/dL |

| Compound | Type | Function or medical use | Tested concentration |
|---|---|---|---|
| Vancomycin | exogenous | Tricyclic glycopeptide antibiotic (Medical use: antibiotic) | 70_µmol/L |
| Water | solvent | solvent for all compounds except LBP, furosemide | 10% v/v |
| Phosphate buffered saline (PBS) | solvent | solvent for LBP | 10% v/v |
| Methanol in water | solvent | solvent for furosemide | 1% MeOH + 9% water v/v |

***Supplement Table S4:*** *Results from the analytical interference study.* Each condition was tested in triplicate, except that bilirubin was tested in six replicates, with one replicate deemed an outlier.

| Compound | Mean | SD | Matrix (carrier) | Mean | \|ΔScore \| | Conclusion |
|---|---|---|---|---|---|---|
| Albumin | 5.67 | 0.153 | water | 5.47 | 0.20 | No interference |
| Bilirubin | 5.54 | 0.207 | water | 5.47 | 0.07 | No interference* |
| CD14, soluble | 5.80 | 0.200 | water | 5.47 | 0.33 | No interference |
| Cefotaxime | 5.00 | 0.200 | water | 5.47 | 0.47 | No interference |
| C-reactive protein | 5.30 | 0.173 | water | 5.47 | 0.17 | No interference |
| Dobutamine | 5.53 | 0.252 | water | 5.47 | 0.06 | No interference |
| Dopamine | 5.27 | 0.321 | water | 5.47 | 0.20 | No interference |
| Furosemide | 5.33 | 0.231 | 10% MeOH | 5.33 | 0.00 | No interference |
| Heparin | 5.37 | 0.115 | water | 5.47 | 0.10 | No interference |
| Imipenem | 5.53 | 0.115 | water | 5.47 | 0.06 | No interference |
| Hemoglobin | 5.63 | 0.058 | water | 5.47 | 0.16 | No interference |
| IL-6 | 5.93 | 0.153 | water | 5.47 | 0.46 | No interference |
| LBP | 5.37 | 0.153 | PBS | 5.23 | 0.14 | No interference |
| Noradrenaline | 5.30 | 0.100 | water | 5.47 | 0.17 | No interference |
| Rheumatoid Factor | 5.70 | 0.173 | water | 5.47 | 0.23 | No interference |
| Triglycerides | 5.87 | 0.252 | water | 5.47 | 0.40 | No interference |

| Compound | Mean | SD | Matrix (carrier) | Mean | \|ΔScore \| | Conclusion |
|---|---|---|---|---|---|---|
| Vancomycin | 5.90 | 0.100 | water | 5.47 | 0.43 | No interference |
| | | | | | | No interference |
| EDTA whole blood | 5.60 | 0.458 | | | | |
| Carrier: 10% water in EDTA whole blood | 5.47 | 0.058 | | | | |
| Carrier: 10% PBS in EDTA whole blood | 5.23 | 0.115 | | | | |
| Carrier: 1% methanol, 9% water in EDTA whole blood | 5.33 | 0.115 | | | | |

*one of six replicates rejected as being an outlier

### 5. Phenotypic Stratification

#### 5.1. PCA /HC analysis on the N=176 sepsis group

**Table S5** presents the quantitative analysis corresponding to Figure 6 in the main text. In this analysis, principal component analysis / hierarchical clustering (PCA /HC) was conducted on the N=176 sepsis group.

***Supplement Table S5:*** *Performance of SeptiCyte RAPID vs. driver variables, for discriminating sepsis vs. SIRS in designated subgroups. Subgroups* are defined by the main drivers of phenotypic variability in PCA. Data corresponds to Figure 6 in main text. AUC confidence intervals (CI) were calculated by formula of Hanley & McNeil (1982), as computed by applet: https://riskcalc.org/ci/ unless noted otherwise to indicate bootstrapping (BR). Note the high % of missing values for the Platelet.Max parameter. For small strata, the two-tailed p-values were calculated with both the t-test (T) and the Mann Whitney U test (U). NS = not significant (p > 0.05).

| | | | | **SeptiScore** | | **Biomarker** | | |
|---|---|---|---|---|---|---|---|---|
| **Phenotypic group** | **Phenotypic subgroup** | **N sepsis** | **N SIRS** | **AUC (95% CI)** | **p-value** | **AUC** | **p-value** | **Missing values (% out of 419)** |
| Blood Properties | WBC low (< 6 E+06/mL) | 30 | 42 | 0.78 (0.67-0.89) | 3.8 E-05 | 0.54 | NS | WBC.Min: 12 (2.9%)<br>WBC.Max: 16 (3.8%) |
| | WBC mid (6-12 E+06/mL) | 30 | 68 | 0.79 (0.68-0.90) | 4.5 E-05 | 0.54 | NS | |
| | WBC high (> 12 E+06/mL) | 112 | 128 | 0.85 (0.80-0.90) | 5.4 E-22 | 0.61 | WBC.Min 0.004<br>WBC.Max 0.0003 | |
| | Platelets < 150,000 / uL | 48 | 66 | 0.82 (0.72-0.90) | 1.9 E-09 | 0.56 (BR) | NS | Platelet.Min: 122 (29%)<br>Platelet.Max: 263 (63%) |
| | platelets 150,000 - 440,000/uL | 68 | 105 | 0.85 (0.79-0.91) | 5.1 E-15 | 0.62 (BR) | Platelet.Min 0.0012 (T)<br>Platelet.Min 0.0037 (U)<br>Platelet.Max 0.0029 (T)<br>Platelet.Max 0.0067 (U) | |
| | platelets > 445,000 / uL | 8 | 8 | 0.73 (0.46-0.92) | NS | 0.74 (0.40-1.00) (BR) | NS | |
| Clinical Chemistry | Lactate <1 mmol/L | 7 | 17 | 0.84 (0.64-1.04) | 8.6 E-03 (T)<br>1.2 E-02 (U) | 0.55 | NS | 140 (33.4%) |
| | Lactate 1-3 mmol/L | 74 | 89 | 0.83 (0.76-0.90) | 8.6 E-14 | 0.52 | NS | |
| | Lactate >3 mmol/L | 48 | 44 | 0.76 (0.66-0.86) | 2.6 E-06 | 0.54 | NS | |
| | PCT <0.5 ng/mL | 32 | 122 | 0.71 (0.60-0.82) | 9.9 E-05 (T)<br>3.2 E-04 (U) | 0.59 | NS | 72 (17.2%) |
| | PCT ≥ 0.5 ng/mL | 117 | 76 | 0.82 (0.76-0.88) | 1.7 E-15 | 0.74 (0.67-0.81) (BR) | 9.4 E-04 | |

| | | | | | | | | |
|---|---|---|---|---|---|---|---|---|
| Vital Signs | MAP < 70 mm Hg always | 33 | 33 | 0.82 (0.71-0.92) | 5.2 E-06 | MAP.Min 0.50 (BR) | NS | MAP.Min: 9 (2.2%)<br>MAP.Max: 95 (22.7%) |
| | MAP 70-100 mm Hg | 28 | 35 | 0.83 (0.71-0.97) | 1.3 E-06 | MAP.Min 0.51 (BR)<br>MAP.Max 0.52 (BR) | NS | |
| | MAP >100 mm Hg<br>(and never <70 mm Hg) | 18 | 56 | 0.86 (0.75-0.97) | 5.5 E-08 | MAP.Min 0.50 (BR)<br>MAP.Max 0.53 (BR) | NS | |
| | MAP >100 mm Hg<br>(above at least once) | 58 | 115 | 0.85 (0.78-0.92) | 3.3 E-18 | MAP.Min 0.63 (BR)<br>MAP.Max 0.50 (BR) | MAP.Min 0.0026 (T)<br>MAP.Min 0.0026 (U)<br>MAP.Max NS | |
| | MAP <70 mm Hg and<br>MAP >100 mm Hg ** | 40 | 59 | 0.86 (0.78-0.94) | 9.4 E-12 | MAP.Min 0.64 (BR)<br>MAP.Max 0.50 (BR) | MAP.Min 0.014 (T)<br>MAP.Min 0.016 (U)<br>MAP.Max NS | |
| | RR.max ≤ 21 breaths/min | 9 | 35 | 0.82 (0.66-0.98) | 0.0003 (T)<br>0.0035 (U) | 0.63 | NS | 124 (29.6%) |
| | RR.max > 21 breaths/min | 114 | 137 | 0.81 (0.76-0.86) | 1.6 E-20 | 0.58 | RR.Max 0.02<br>RR.Min 0.02 | RR.Min: 129 (30.8%)<br>RR.Max: 124 (29.6%) |
| | Systol. BP.max/min ≤100 mm Hg | 63 | 73 | 0.82 (0.75-0.89) | 5.4 E-12 | 0.57 | NS | SBP.Min: 219 (52.3%)<br>SBP.Max: 217 (51.8%) |
| | Systol. BP.max/min >100 mm Hg | 17 | 47 | 0.84 (0.72-0.96) | 8.9 E-07 (T)<br>4.6 E-05 (U) | 0.60 | NS | |
| Organ Dysfunction | SOFA (≥ 2) *** | 141 | 175 | 0.82 (0.77-0.86) | 1.21 E-27 (T)<br>< 1 E-14 (U) | 0.59 (0.53-0.65) (BR) | 0.015 (T)<br>0.0047 (U) | 46 (11.0%) |
| | SOFA (<2) *** | 16 | 41 | 0.79 (0.65-0.91) | 3.8 E-04 (T)<br>6.8 E-04 (U) | 0.49 (BR) | NS | |

*MAP.Max measurement available for only 3 of 66 patients in this stratum

** Individual patients for whom the MAP parameter was measured outside the 70-100 mm Hg range in both directions

*** Patients with partial SOFA scores included in the calculation. A total of 270/419 (64.4%) patients had complete SOFA scores reported. The number (%) of patients with partial SOFA scores were: 72/419 (17.2%) with 1of 6 components missing; 6/419 (1.4%) with 2 of 6 components missing; 8/419 (1.9%) with 3 of 6 components missing, 1/419 (0.24%) with 4 of 6 components missing, 0 with 5 of 6 components missing. There were 46/419 (11.0%) of patients with no SOFA components entered in the line data, and these were not used in the calculation.

#### 5.2. PCA /HC analysis on the entire N=419 cohort

We next repeated the PCA/ HC analysis on the entire (N=419) cohort, without first separating the sepsis and SIRS patients. In dimensions 1 & 2 in this alternative PCA / HC analysis, the most significant drivers of variance (i.e. the variables with greater than an average expected contribution of 6.25%) were WBC (max & min), glucose (min & max), platelets (min), heart rate (min & max), MAP.Min, and temperature (max). These driving variables were the same as identified from the previous PCA / HC analysis conducted on just the N=176 sepsis group. A visual representation of the PCA /HC analysis on the N=419 complete cohort is shown in **Figure S2**.

***Supplement Figure S2***: *PCA plot of the entire study cohort (N=419) with superimposed HC.* This analysis used the 9 phenotypic variables identified as the main PCA drivers, plus an additional 7 variables often used in sepsis adjudication (16 phenotypic variables total). Subgroup 1 (black) contains 88 sepsis and 184 SIRS. Subgroup 2 (red) contains 11 sepsis and 18 SIRS. Subgroup 3 (green) contains 77 sepsis and 41 SIRS. The first 3 dimensions of the PCA explained 36.8% of the variance (Dim 1: 14.9%, Dim2: 11.34%, Dim3: 10.54%).

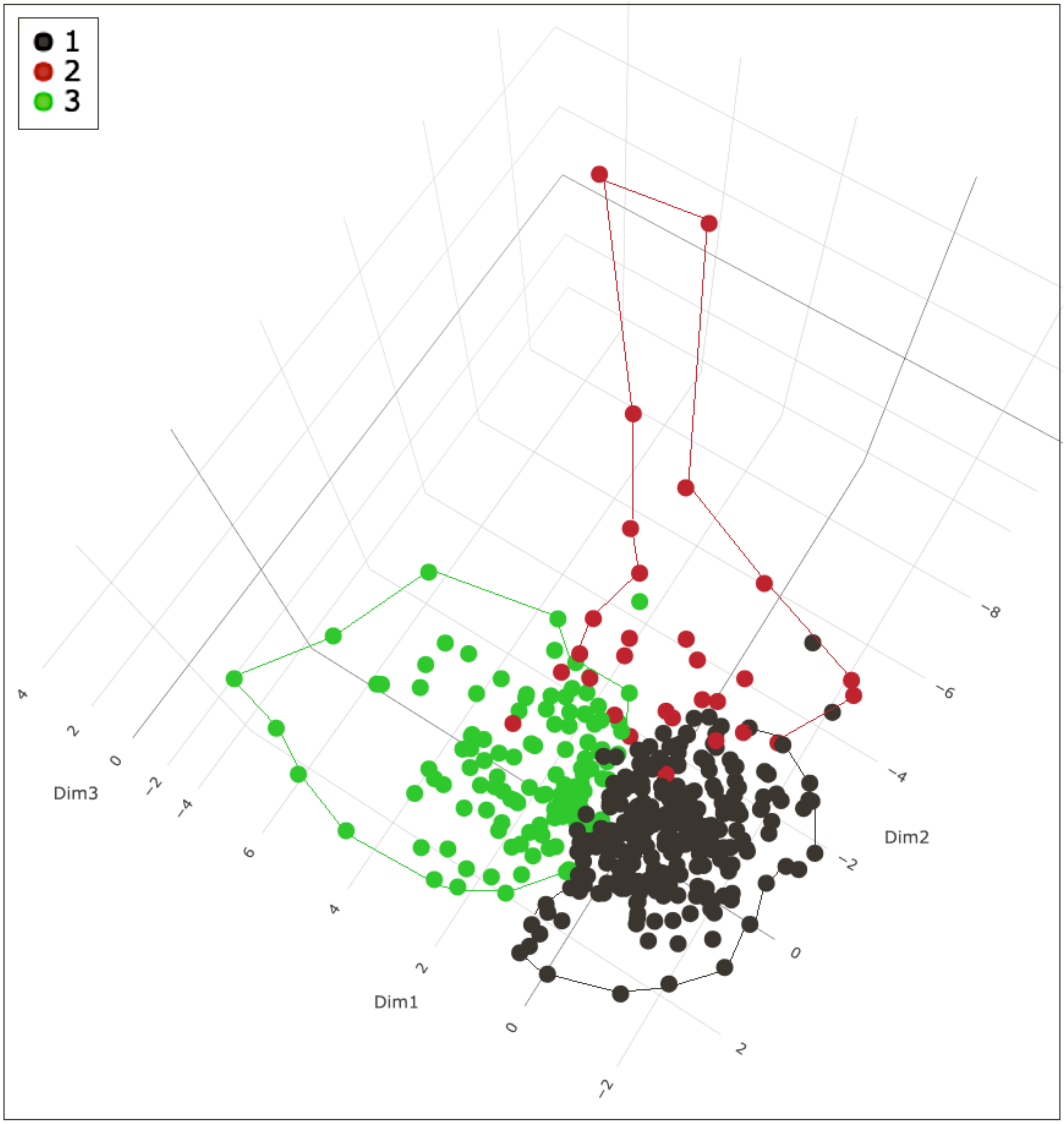

We evaluated the ability of SeptiCyte RAPID to discriminate sepsis vs. SIRS in each of the phenotypic subgroups in Figure S2. The analysis indicates that the test performance is high across the subgroup 1 (88 sepsis vs. 184 SIRS; AUC = 0.79, p = 5.9 E-14) and subgroup 3 (77 sepsis vs. 41 SIRS; AUC = 0.85, p = 4.3 E-13). The test performance appears even higher for subgroup 2 (11 sepsis vs. 18 SIRS; AUC = 0.89, p = 0.001), although the small patient numbers in this group imply a relatively larger uncertainty in the calculated AUC and p-values.

#### 5.3 Concordance between PCA / HC analyses on N=176 sepsis group and N=419 cohort

We next determined the degree of concordance between the PCA / HC analysis conducted on just the N=176 sepsis group (main text, Figure 5), and the PCA / HC analysis conducted on the N=419 entire cohort (supplementary material, Figure S2). Concordance was defined as agreement in the classification of sepsis patients into different subgroups. Results of the concordance analysis are presented in **Table S6.** The unweighted concordance statistic $\kappa = 0.562\pm0.058$ according to the method of Cohen (1960), and $\kappa = 0.562\pm0.052$ according to the modified calculation by Fleiss et al. (1969). Kappa falls within the range 0.40-0.60, and therefore (according to the interpretation of Cohen, 1960) indicates a moderate agreement between the two PCA / HC analyses.

***Supplement Table S6****: Concordance between the two PCA / HC analyses.* Calculation of $\kappa$ was performed with the applet at http://vassarstats.net/kappa.html.

<table>
<tr><td colspan="2" rowspan="2"></td><td colspan="4">PCA / HC on sepsis group (N=176)</td><td rowspan="2"></td></tr>
<tr><td>sepsis subgroup 1</td><td>sepsis subgroup 2</td><td>sepsis subgroup 3</td><td>sepsis subgroup 4</td></tr>
<tr><td rowspan="4">PCA / HC on entire cohort (N=419)</td><td>sepsis subgroup 1</td><td>84</td><td>3</td><td>1</td><td>0</td><td>Σ = 88</td></tr>
<tr><td>sepsis subgroup 3</td><td>20</td><td>46</td><td>11</td><td>0</td><td>Σ = 77</td></tr>
<tr><td>sepsis subgroup 2</td><td>6</td><td>1</td><td>3</td><td>1</td><td>Σ = 11</td></tr>
<tr><td>sepsis subgroup 4</td><td>0</td><td>0</td><td>0</td><td>0</td><td>Σ = 0</td></tr>
<tr><td colspan="2"></td><td>Σ = 110</td><td>Σ = 50</td><td>Σ = 15</td><td>Σ = 1</td><td>Σ Σ = 176</td></tr>
</table>

#### 5.4 k-means clustering analysis on N=419 cohort

We next conducted a k-means clustering analysis on the complete N=419 patient cohort. The algorithm determined that the optimal number of k-clusters should be 2. Results of this k-means analysis are shown in **Figure S3.**

***Supplement Figure S3****: Resolution of the N=419 cohort into two subgroups, based on k-means clustering.* A value k=2 was identified as optimal by the clustering algorithm. The large symbols indicate the centroids of the groups. Subgroup 1 (N=178) consists of 53 sepsis and 125 SIRS. Subgroup 2 (N=241) consists of 123 sepsis and 118 SIRS.

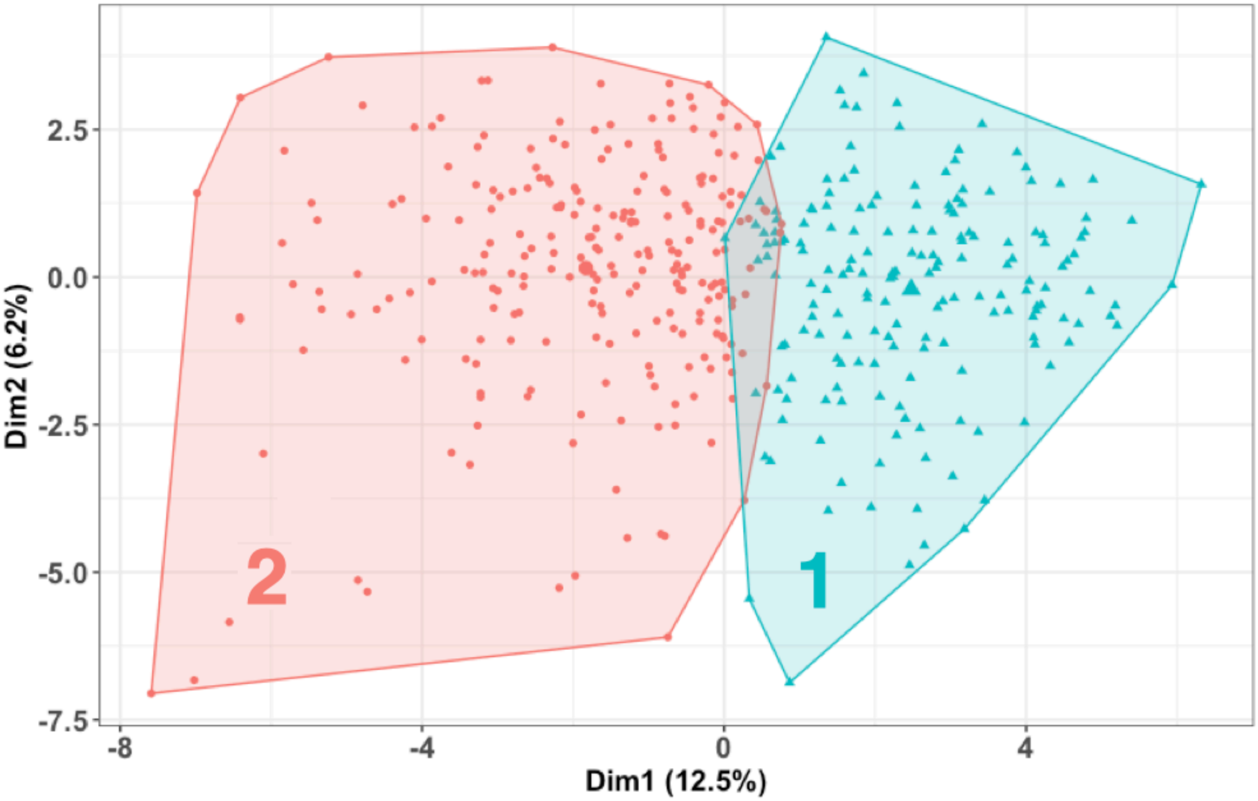

The performance of SeptiCyte RAPID for discrimination of sepsis vs. SIRS is comparable between the two subgroups (**Table S7**).

***Supplement Table S7**: Performance of SeptiCyte RAPID for discrimination between sepsis and SIRS in the different phenotypic strata identified by the k-means clustering analysis.*

| Phenotypic stratum | N sepsis | N SIRS | AUC | p-value |
|---|---|---|---|---|
| Fig S3, subgroup 1 (N=178) | 53 | 125 | 0.80 | 7.1 E-09 |
| Fig S3, subgroup 2 (N=241) | 123 | 118 | 0.82 | 2.4 E-20 |

An ANOVA was performed to identify those phenotypic variables contributing to separation between the subgroups in the k-means analysis of Figure S3. Results of the ANOVA, showing only those variables with $p<0.01$, are presented in **Table S8**. Regarding differences between the two subgroups, it appears that subgroup 2 (N=241) is more seriously ill, being characterized by higher glucose max, qSOFA and SOFA; greater percentage of positive culture results; and higher use of vasopressors and mechanical ventilation.

***Supplement Table S8***: *Phenotypic variables contributing to separation between subgroups in the k-means analysis of Figure S3.* Only variables with p<0.01 by one-way ANOVA are presented in this table. Abbreviations: GCS, Glasgow Coma Score; PCT, procalcitonin; RR, respiratory rate; SOFA, sequential organ failure assessment; WBC, white blood cells.

| **Characteristic** | **Missing values (%)** | **Subgroup 1 (N = 178), median (IQR) or N (%)** | **Subgroup 2 (N = 241), median (IQR) or n (%)** | **p-value** |
|---|---|---|---|---|
| Age | 0 (0%) | n=57 (42-68) | n=61 (48-71) | 0.009 |
| Bacterial culture positive | NA | n=45 (25%) | n=104 (43%) | <0.001 |
| Molecular or immunological test or culture(s) positive | NA | n=10 (5.6%) | n=32 (13%) | 0.01 |
| Glucose (max) >110 mg/dl | 29 (7%) | n=115 (64.6%) | n=215 (89.2%) | <0.001 |
| Heart Rate (Max), bpm | 0 (0%) | 106 (92-123) | 119 (107-138) | <0.001 |
| Lactate, mmol/L | 140 (33%) | 2.00 (1.30-2.60) | 2.45 (1.60-4.08) | 0.005 |
| Mean Art Pressure (Min), mm Hg | 9 (2%) | 74 (65-84) | 57 (49-64) | <0.001 |
| PCT, ng/mL | 77 (18%) | 0 (0-1) | 2 (0-11) | <0.001 |
| PCT_binary (>0.5 ng/mL) | 77 (18%) | n=140 (78.7%) | n=53 (22%) | <0.001 |
| qSOFA | 31 (7%) | 0 (0-1) | 2 (2-2) | <0.001 |
| qSOFA (GCS component) | 58 (14%) | n=19 (10.7%) | n=115 (47.7%) | <0.001 |
| qSOFA (RR component) | 124 (30%) | n=54 (30.3%) | n=197 (81.7%) | <0.001 |
| Respiratory Rate (Max), bpm | 124 (30%) | 24 (21-27) | 26 (24-32) | <0.001 |
| Respiratory Rate (Min), bpm | 129 (31%) | 15 (11-21) | 18 (13-24) | 0.009 |
| qSOFA>=2 | 31 (7%) | n=13 (7.3%) | n=175 (72.6%) | <0.001 |
| qSOFA<2 | | n=153 (86%) | n=47 (19.5%) | <0.001 |
| SOFA score | 46 (11%) | 3.0 (1.0-4.0) | 7.0 (5.8-10.0) | <0.001 |
| Temperature (Min), °C | 64 (15%) | 36.30 (36.00-36.70) | 35.60 (34.90-36.30) | <0.001 |
| Vasopressors (Y/N) | 0 (0%) | n=15 Y (8.4%) | n=123 Y (51%) | <0.001 |
| Mechanical ventilation (Y/N) | 0 (0%) | n=31 Y (17%) | n=122 Y (51%) | <0.001 |
| WBC (Max) | 16 (4%) | 11 (8-16) | 15 (11-20) | <0.001 |
| WBC (Min) | 12 (3%) | 9.4 (6.4-12.4) | 11.9 (7.4-15.9) | <0.001 |

#### 5.4 Concordance between k-means analyses on N=176 sepsis group and N=419 cohort

Finally, we determined the concordance between the k-means analyses on the N=176 sepsis group (Figure 7 in the main text), and the k-means analyses on the N=419 entire cohort (Supplement Figure S3). Concordance was defined as agreement in the classification of sepsis patients into different subgroups. Results of this concordance analysis are presented in **Table S9.** The unweighted concordance statistic $\kappa = 0.528 \pm 0.062$ according to the method of Cohen (1960), and $\kappa = 0.528 \pm 0.055$ according to the modified calculation by Fleiss et al. (1969). Kappa falls within the range 0.40-0.60, and therefore (according to the interpretation of Cohen, 1960) indicates a moderate agreement between the two k-means analyses.

***Supplement Table S9:*** *Concordance between the two k-means analyses, according to Cohen's* $\kappa$. Calculations were performed with the applet at http://vassarstats.net/kappa.html./

| | | k-means on entire cohort (N=419) (Figure S3, Supplement) | | |
|---|---|---|---|---|
| | | Sepsis subgroup 1 | Sepsis subgroup 2 | |
| k-means on sepsis group (N=176) (Figure 7, main text) | Sepsis subgroup 1 | 53 | 43 | Σ = 96 |
| | Sepsis subgroup 2 | 0 | 80 | Σ =80 |
| | | Σ =53 | Σ =123 | Σ Σ = 176 |

### 6. Notes on Statistical Analysis of Small Strata

A number of analyses described in the main text or in this Supplement were performed on strata with relatively small N. We have investigated the effect of small N on the quantitative measure (AUC) used to evaluate SeptiCyte RAPID performance.

This section is organized as follows:

6.1 ROC curves for SeptiCyte RAPID performance in strata with small N
6.2 Sensitivity Analysis: Confidence Interval (CI) for AUC with small N
6.3 Further Investigation of Bootstrap Estimates of CI with small N

#### 6.1 ROC curves for different strata with small N

**Figures S4, S5, S6, S7** presents graphical ROC curves for selected strata with small numbers in one or both groups, or for which the group numbers are unbalanced. In these panels, p = probability of the observed AUC value, compared to the null hypothesis that the true AUC value is 0.5 (Hanley & McNeil, 1982; Zweig & Campbell, 1993; Shoonjans, 2017). For all analyses except platelets > 440,000/uL (Figure S7, plot #20) the ROC curves are statistically distinguishable from the null AUC 0.5 diagonal line.

***Supplement Figure S4:*** *ROC curves for demographic strata with small numbers in one or both groups, or for which the group numbers are unbalanced.* The order of panels corresponds to the order of entries in Table T10 below. Abbreviations: case = SeptiScores from sepsis patients; con = SeptiScores from SIRS patients.

1. Asian (10 case, 11 con)

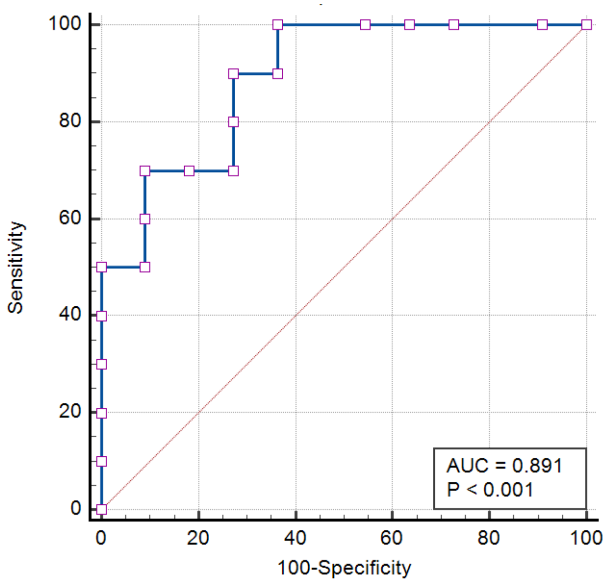

2. Hispanic (10 case, 12 con)

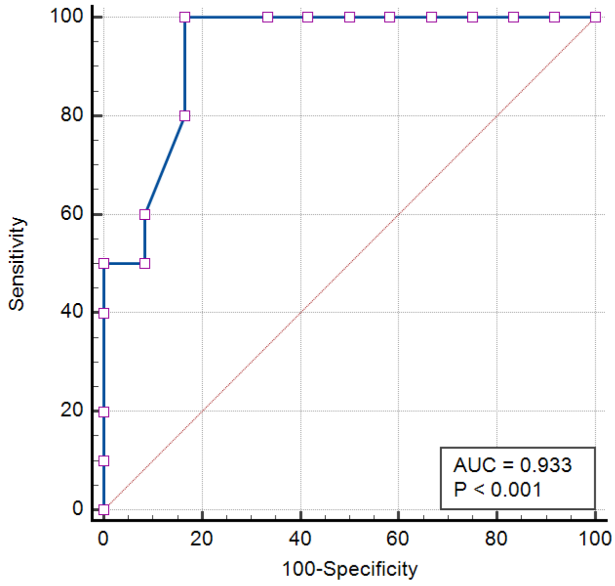

3. Black: Septic shock vs. SIRS (16 case, 70 con)

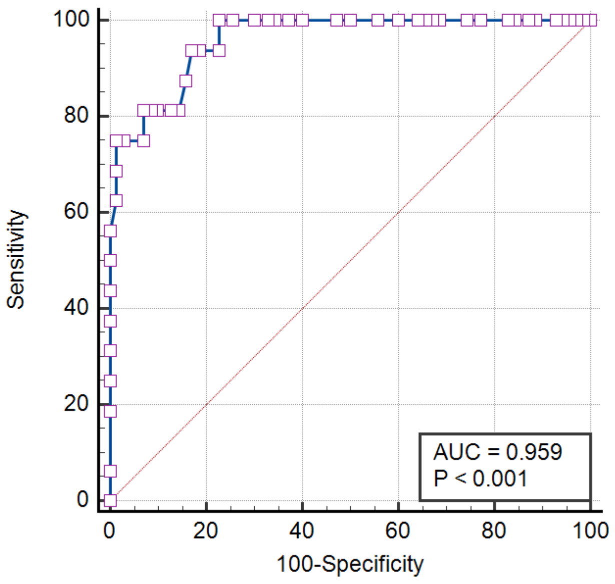

4. White: Septic shock vs. SIRS (44 case, 146 con)

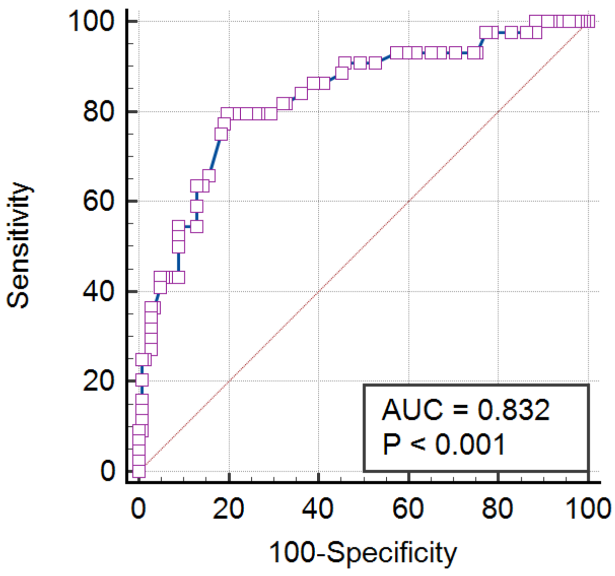

***Supplement Figure S5:*** *ROC curves for comorbidity strata with small numbers in one or both groups, or for which the group numbers are unbalanced.* The order of panels corresponds to the order of entries in Table T11 below. Abbreviations: case = SeptiScores from sepsis patients; con = SeptiScores from SIRS patients.

5. diabetic hyperglycemia (10 case, 16 con)

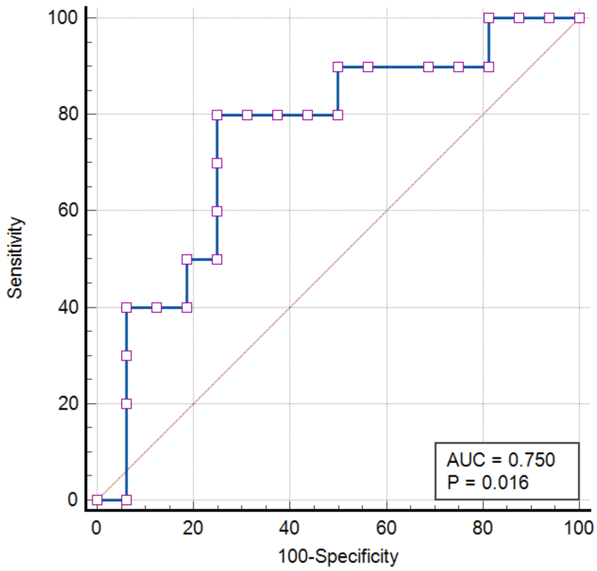

6. impaired immunity (27 case, 32 con)

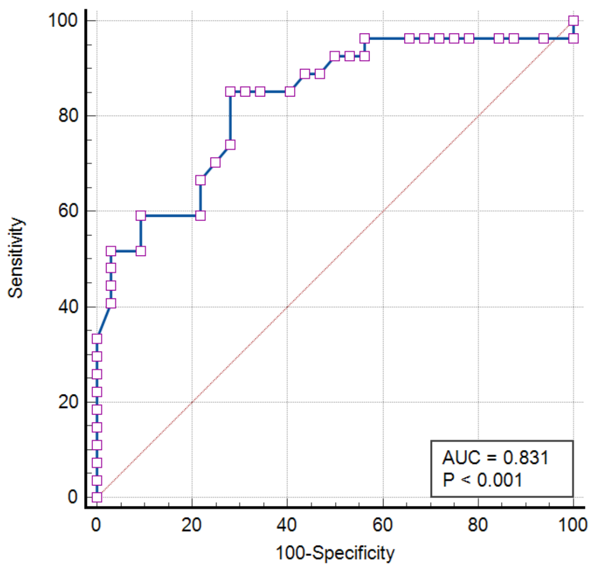

7. hypertension (24 case, 26 con)

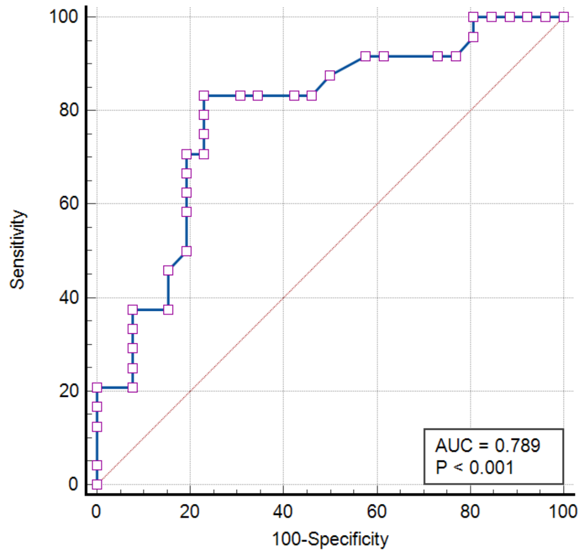

8. cardiovasc. disease, acute (7 case, 6 con)

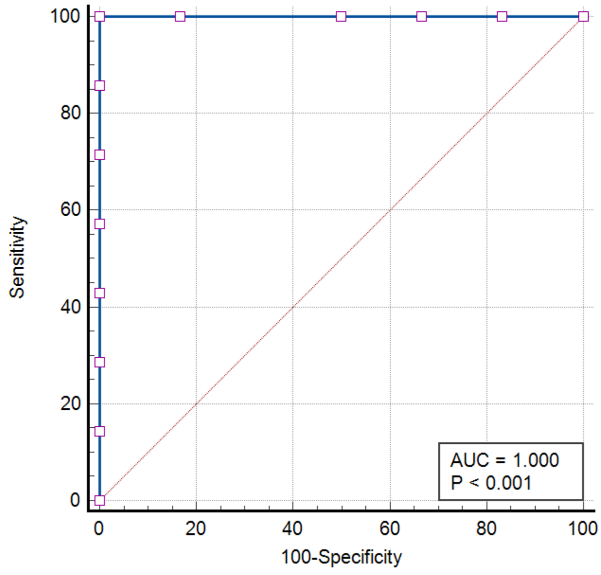

9. cardiovasc. disease, chronic (9 case, 17 con)

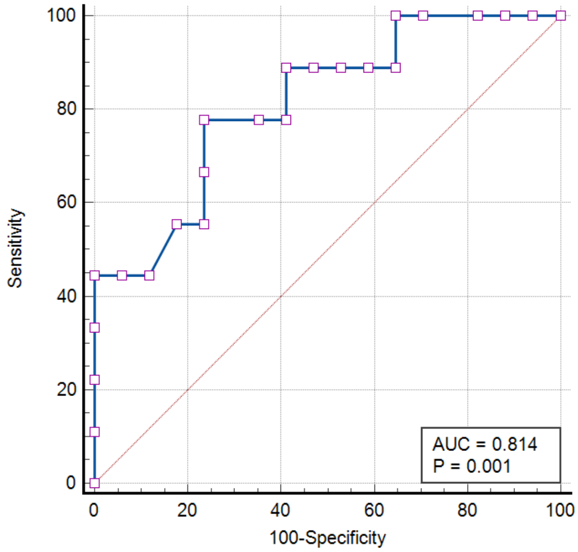

10. kidney disease, acute (13 case, 13 con)

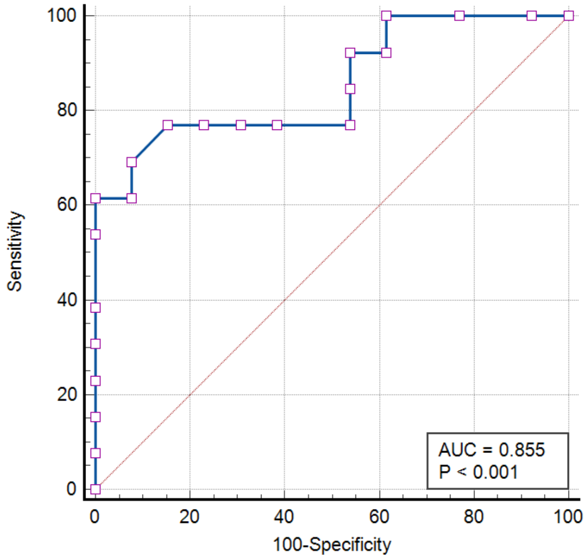

11. kidney disease, chronic (20 case, 21 con)

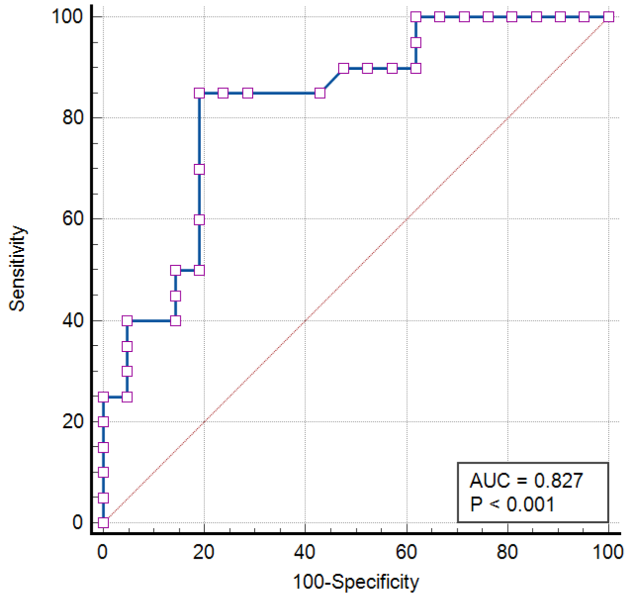

12. obesity (6 case, 14 con)

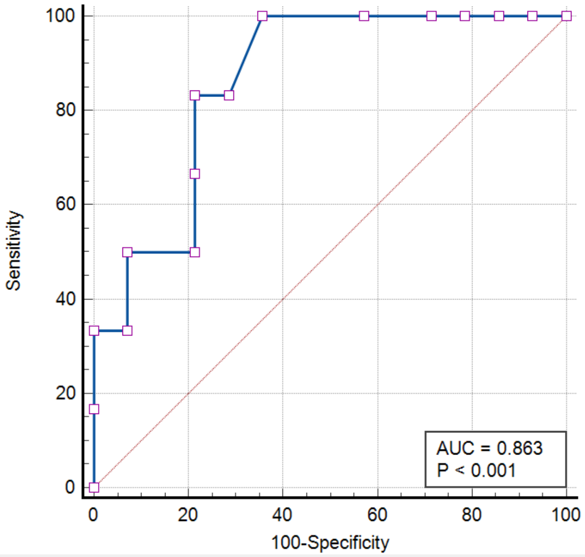

***Supplement Figure S6:*** *ROC curves for infection source strata with small numbers in one or both groups, or for which the group numbers are unbalanced.* The order of panels corresponds to the order of entries in Table T12 below. Abbreviations: case = SeptiScores from sepsis patients; con = SeptiScores from SIRS patients.

13. CNS source (5 case, 207 con)

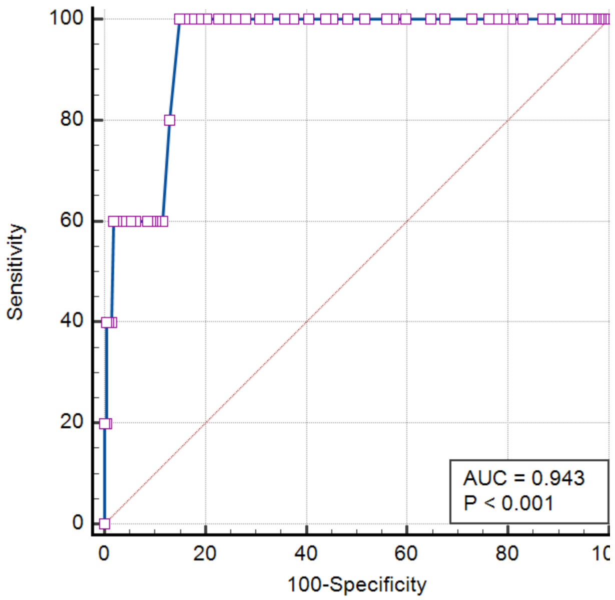

14. Other source (14 case, 207 con)

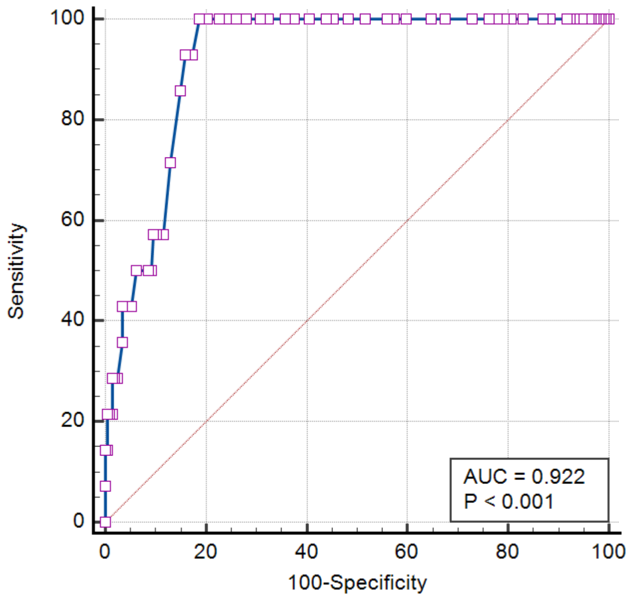

15. Blood source (16 case, 207 con)

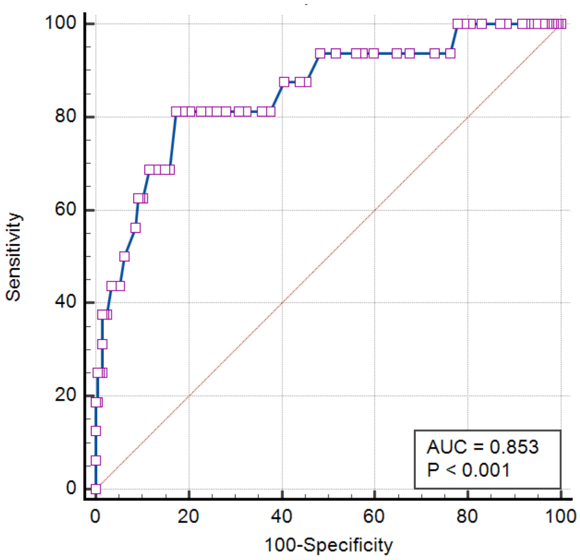

16. Source unidentified (19 case, 207 con)

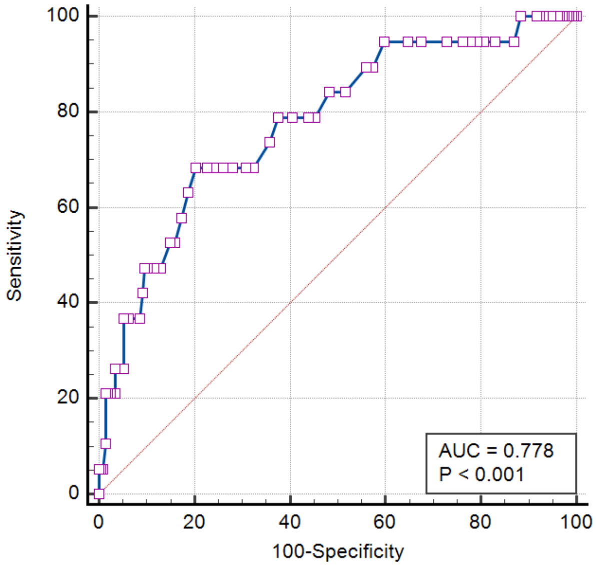

17. Urinary source (23 case, 207 con)

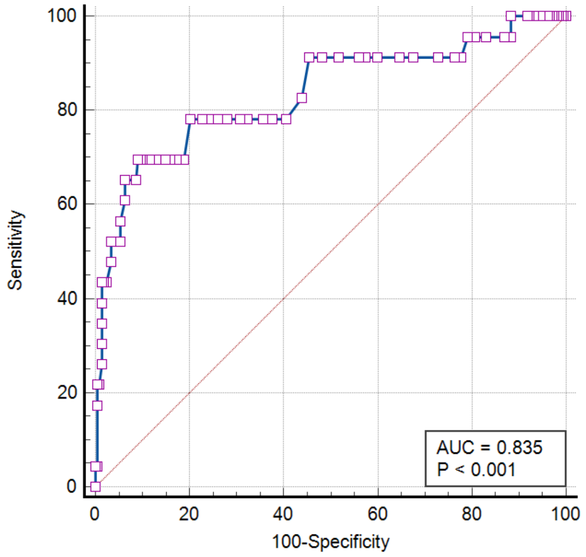

18. Abdominal source (27 case, 207 con)

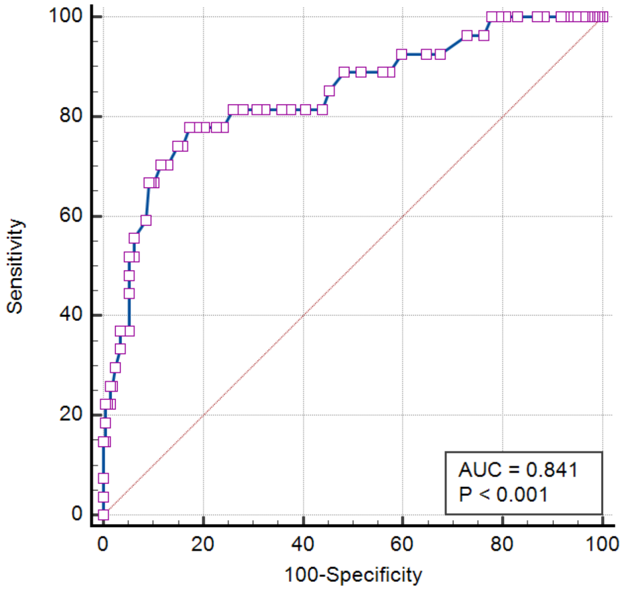

19. Pulmonary source (50 case, 207 con)

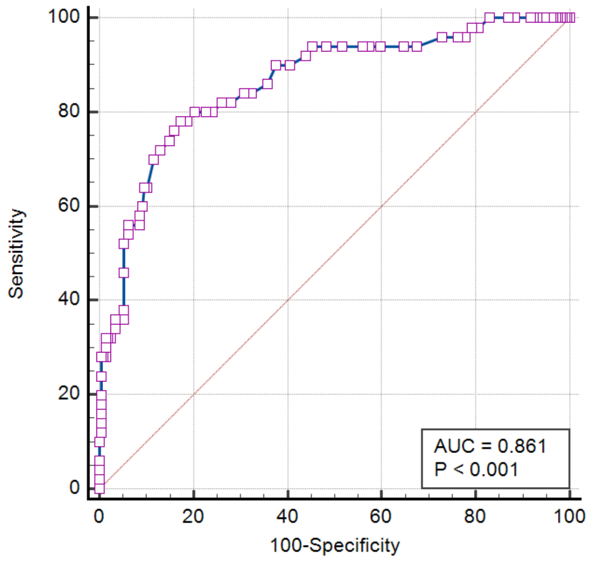

***Supplement Figure S7:*** *ROC curves for strata based on clinical variables, with small numbers in one or both groups, or for which the group numbers are unbalanced.* The order of panels corresponds to the order of entries in Table T13 below. Abbreviations: case = SeptiScores from sepsis patients; con = SeptiScores from SIRS patients.

20. platelets > 445,000/uL (8 case, 8 con)

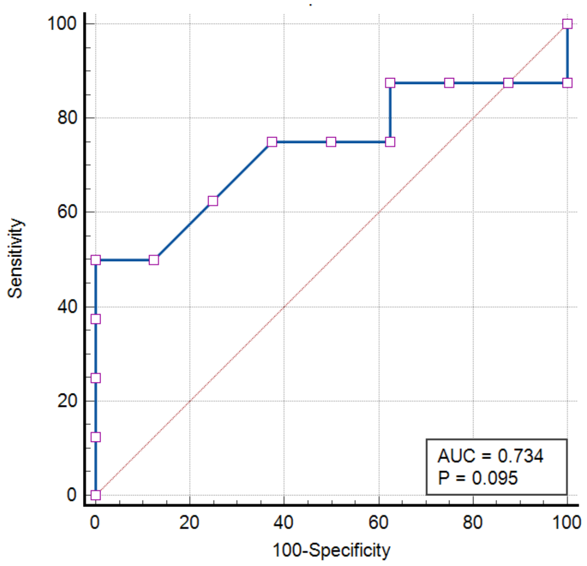

21. Lactate <1 mmol/L (7 case, 17 con)

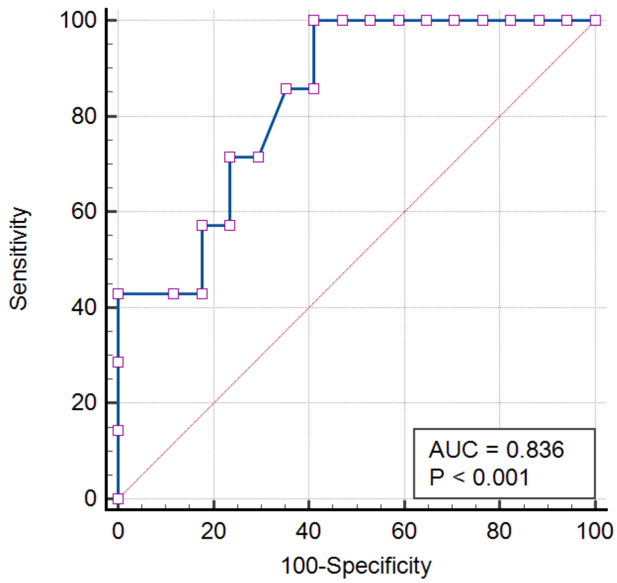

22. PCT <0.5 ng/mL (32 case, 122 con)

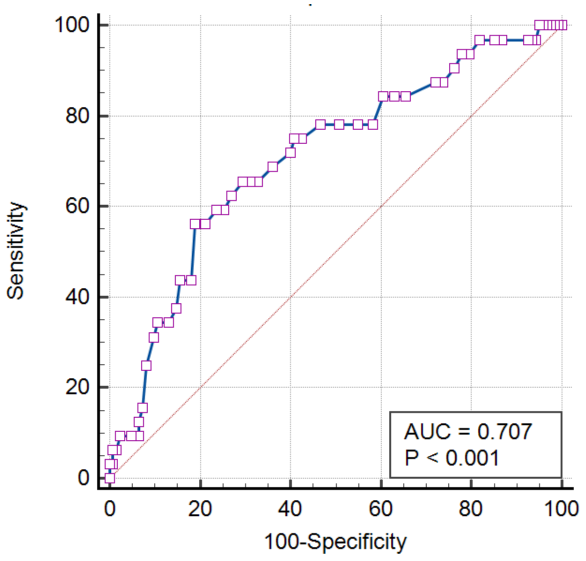

23. MAP <70 mm Hg always (33 case, 33 con)

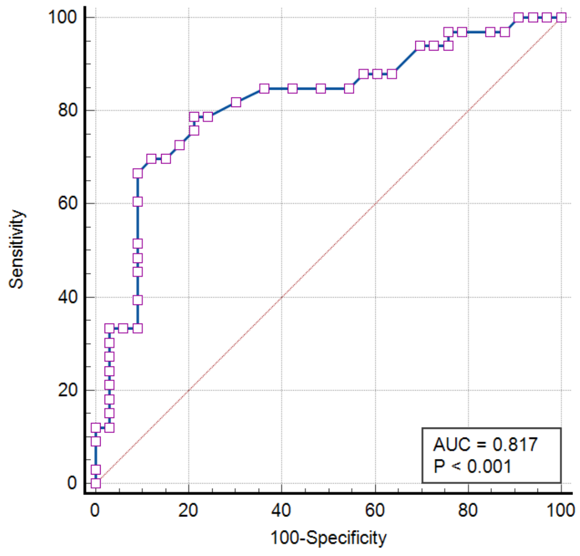

24. MAP 70-100 mm Hg (28 case, 35 con)

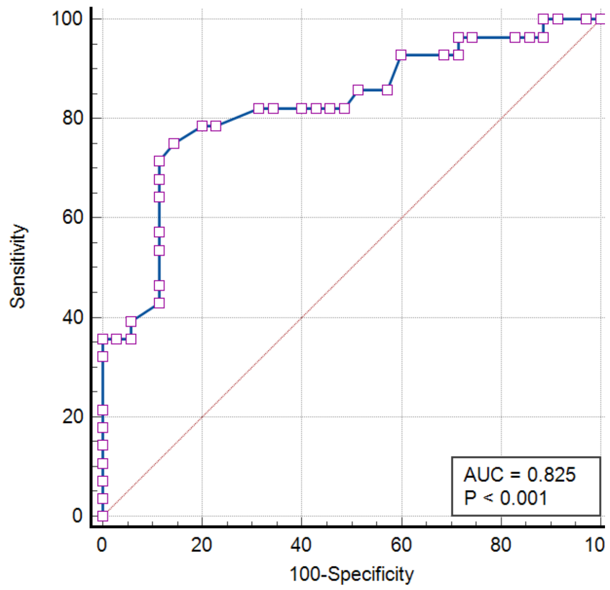

25. MAP >100, not <70 mm Hg (18 case, 56 con)

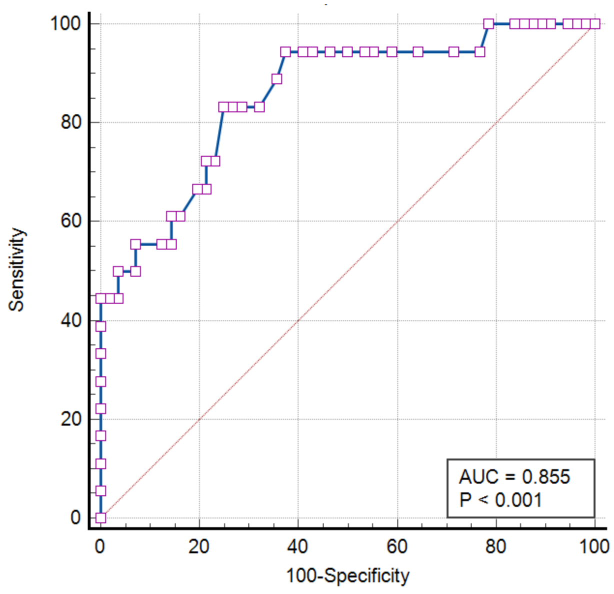

26. MAP >100 mm Hg 1-2x (58 case, 115 con)

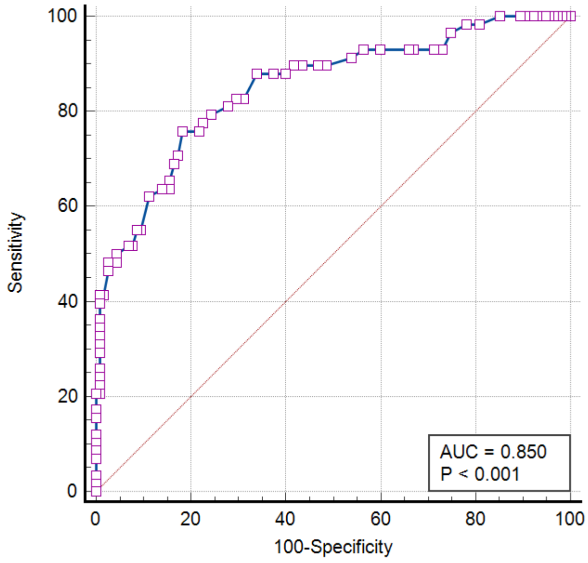

27. MAP <70 and >100 mm Hg (40 case, 59 con)

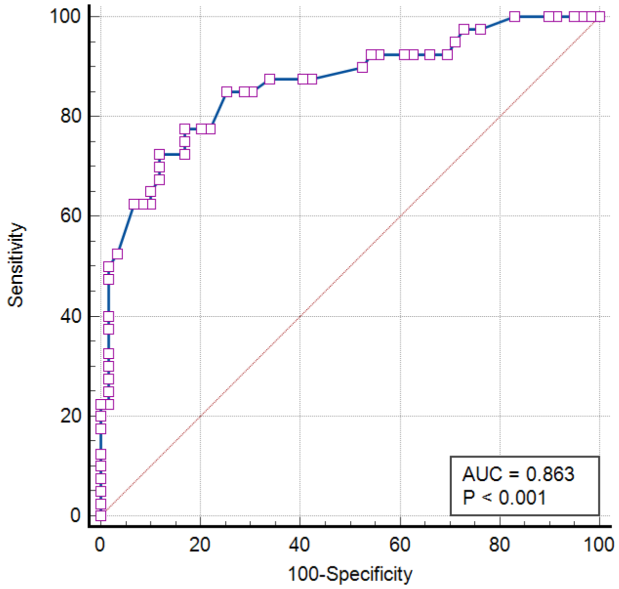

28. RR.max ≤ 21 breaths/min (9 case, 35 con)

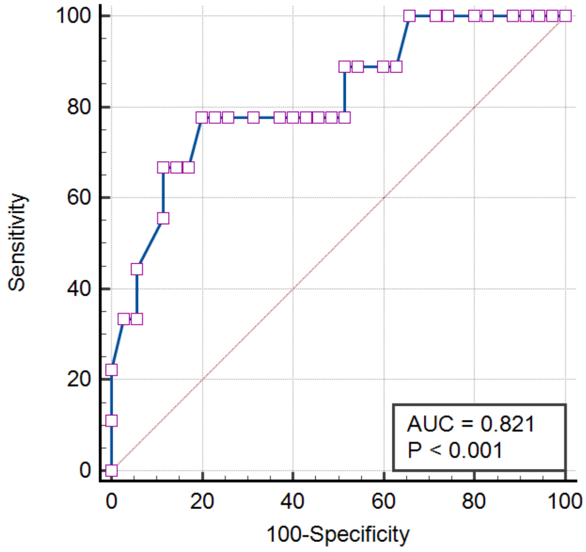

29. Systolic BP >100 mm Hg (17 case, 47 con)

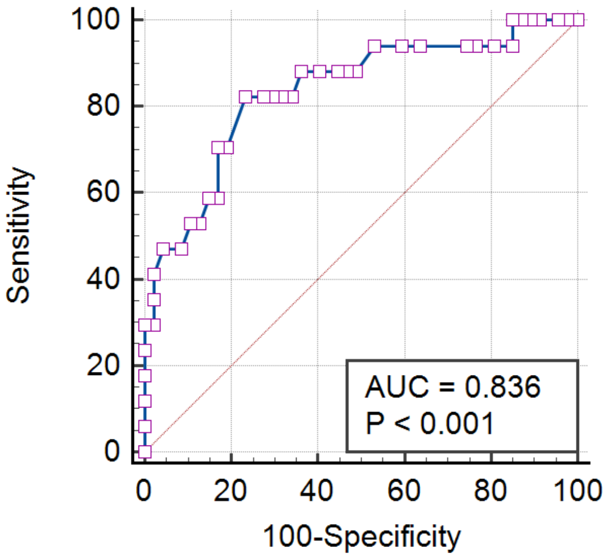

#### 6.2 Sensitivity Analysis: AUC and CI by different methods

To gain assurance that our estimates of AUC and CI for small strata are self-consistent and accurate, we compared four different AUC calculation methods: Binomial Exact, Hanley & McNeil (1982), De Long et al. (1988), and Bootstrapping.

BE – Binomial Exact. This method is based on a probabilistic interpretation of the AUC (Mason & Graham, 2002; Zhou et al., 2011; Anonymous (biyee.net), 2019; Goldstein-Greenwood, 2022), which asserts (in terms relevant to this manuscript) the following: Success = Event where SeptiScore (sepsis patient) > SeptiScore (SIRS patient), and P(Success) = AUC. The underlying basis for this interpretation is the equivalence relation $AUC = U/(n_1*n_2)$ where U = the Mann-Whitney U statistic and $n_1$, $n_2$ are the sizes of the SIRS and sepsis groups (Mason & Graham, 2002; Xu et al., 2013). Consequently P(Success) is a binomial random variable, with a confidence interval (CI) calculated by the Clopper-Pearson binomial exact method, or by a Normal approximation for large sample sizes. (For a situation in which $AUC * (n_1 + n_2)$ does not equal an integer, the CI is calculated for the next-lowest and next-highest integers and a linear interpolation is performed.)

HM – The method of Hanley & McNeil (1982) was used to estimate the Standard Error (SE) of the AUC. Following this, the 95% CI is calculated as AUC ± 1.96 SE.

DL – The method of De Long et al. (1988) was used to estimate the Standard Error (SE) of the ROC AUC. Following this, the 95% CI is calculated as AUC ± 1.96 SE.

BR – Bootstrap resampling with replacement was performed with the web applet of Skalsk´a & Freylich (2006), available at http://www.freccom.cz/stomo/input.php. Multiple bootstraps were conducted, with the number of resamples ranging from 1,000 to 5,000. The 95% CI was estimated directly from the resampled AUC distributions.

Results from these comparisons are summarized in **Tables S10**, **S11, S12, S13**. A general conclusion for confidence interval (CI) widths appears to be that widths increase in the order BE < BR < HM ≈ DL. This relation generally holds, although several exceptions were noted in the tables below. As expected, the CI widths decrease as the group sizes increase.

***Supplement Table S10:*** *ROC analyses for different strata, based on demographic variables (calculations with MedCalc software).* Abbreviations: CI, confidence interval; BE, binomial exact; HM, Hanley & McNeill (1982); DL, DeLong et al. (1988). Entries arranged in order of increasing N.

| **No.** | **Reference** | **Stratum** | **AUC** | **95% CI** | **CI width** | **p*** | **N** | **Case (sepsis)** | **Control (SIRS)** | **Control/case ratio** |
|---|---|---|---|---|---|---|---|---|---|---|
| 1 | Table 1 | Asian | 0.891 | 0.678-0.983 (BE)<br>0.755-1.000 (HM)<br>0.755-1.000 (DL) | 0.305 (BE)<br>0.245 (HM)<br>0.245 (DL) | <0.001 | 21 | 10 | 11 | 1.10 |
| 2 | Table 1 | Hispanic | 0.933 | 0.741-0.995 (BE)<br>0.830-1.000 (HM)<br>0.831-1.000 (DL) | 0.254 (BE)<br>0.170 (HM)<br>0.169 (DL) | <0.001 | 22 | 10 | 12 | 1.20 |
| 3 | Figure S1 | Black: Septic Shock vs. SIRS | 0.96 | 0.893-0.990 (BE)<br>0.918-1.000 (HM)<br>0.918-1.000 (DL) | 0.097 (BE)<br>0.082 (HM)<br>0.082 (DL) | <0.001 | 86 | 16 | 70 | 4.38 |
| 4 | Figure S1 | White: Septic Shock vs. SIRS | 0.83 | 0.772-0.883 (BE)<br>0.761-0.904 (HM)<br>0.760-0.904 (DL) | 0.111 (BE)<br>0.143 (HM)<br>0.144 (DL) | <0.001 | 190 | 44 | 146 | 3.32 |

p* = probability that the observed AUC is different from the null hypothesis value of 0.5 (Hanley & McNeil, 1982; Zweig & Campbell, 1993; Shoonjans, 2017).

***Supplement Table S11:*** *ROC analyses for different strata, based on comorbidities (calculations with MedCalc software).* Abbreviations: CI, confidence interval; BE, binomial exact; HM, Hanley & McNeill (1982); DL, DeLong et al. (1988). Entries arranged in order to match the entries in Table 2.

| No. | Reference | Stratum | AUC | 95% CI | CI width | p* | N | Case (sepsis) | Control (SIRS) | Control/case ratio |
|---|---|---|---|---|---|---|---|---|---|---|
| 5 | Table 2 | diabetic hyperglycemia | 0.750 | 0.543-0.897 (BE) | 0.354 (BE) | 0.016 | 26 | 10 | 16 | 1.60 |
| 6 | Table 2 | impaired immunity | 0.831 | 0.711-0.916 (BE) | 0.205 (BE) | <0.001 | 59 | 27 | 32 | 1.18 |
| 7 | Table 2 | hypertension | 0.789 | 0.651-0.892 (BE) | 0.241 (BE) | <0.001 | 50 | 24 | 26 | 1.08 |
| 8 | Table 2 | cardiovascular disease, acute | 1.000 | 0.753-1.000 (BE) | 0.247 (BE) | <0.001 | 13 | 7 | 6 | 0.86 |
| 9 | Table 2 | cardiovascular disease, chronic | 0.814 | 0.613-0.938 (BE) | 0.325 (BE) | 0.001 | 26 | 9 | 17 | 1.89 |
| 10 | Table 2 | kidney disease, acute | 0.855 | 0.662-0.961 (BE)<br>0.706-1.000 (HM)<br>0.704-1.000 (DL) | 0.299 (BE)<br>0.294 (HM)<br>0.296 (DL) | <0.001 | 26 | 13 | 13 | 1.00 |
| 11 | Table 2 | kidney disease, chronic | 0.827 | 0.677-0.927 (BE) | 0.250 (BE) | <0.001 | 41 | 20 | 21 | 1.05 |
| 12 | Table 2 | obesity | 0.863 | 0.637-0.974 (BE) | 0.337 (BE) | <0.001 | 20 | 6 | 14 | 2.33 |

p* = probability that the observed AUC is different from the null hypothesis value of 0.5 (Hanley & McNeil, 1982; Zweig & Campbell, 1993; Shoonjans, 2017).

***Supplement Table S12:*** *ROC analyses for different strata, based on the initial identification of infection source (calculations with MedCalc software).* Abbreviations: AUC, area under curve; CI, confidence interval; BE, binomial exact; HM, Hanley & McNeill (1982); DL, DeLong et al. (1988); $BR_{NNNN}$, bootstrap resampling with NNNN cycles. Entries arranged in order of increasing N.

| No. | Reference | Infection Source (initial identification) | AUC | 95% CI | CI width | p* | N | case | control | Control/case ratio |
|---|---|---|---|---|---|---|---|---|---|---|
| 13 | Figure 2 | CNS | 0.943 | 0.903-0.970 (BE)<br>0.885-1.000 (HM)<br>0.880-1.000 (DL) | 0.067 (BE)<br>0.115 (HM)<br>0.120 (DL) | <0.001 | 212 | 5 | 207 | 41.4 |
| 14 | Figure 2 | other | 0.922 | 0.878-0.953 (BE)<br>0.879-0.964 (HM)<br>0.879-0.965 (DL) | 0.075 (BE)<br>0.085 (HM)<br>0.086 (DL) | <0.001 | 221 | 14 | 207 | 14.79 |
| 15 | Figure 2 | blood | 0.853 | 0.799-0.896 (BE)<br>0.747-0.958 (HM)<br>0.744-0.961 (DL) | 0.097 (BE)<br>0.211 (HM)<br>0.217 (DL) | <0.001 | 223 | 16 | 207 | 12.94 |
| 16 | Figure 2 | not identified | 0.778 | 0.718-0.830 (BE)<br>0.666-0.889 (HM)<br>0.664-0.892 (DL)<br>0.672-0.888 ($B_{2000}$)<br>0.669-0.886 ($B_{5000}$) | 0.112 (BE)<br>0.223 (HM)<br>0.228 (DL)<br>0.216 ($BR_{2000}$)<br>0.217 ($BR_{5000}$) | <0.001 | 226 | 19 | 207 | 10.89 |
| 17 | Figure 2 | urinary | 0.835 | 0.781-0.881 (BE)<br>0.730-0.940 (HM)<br>0.728-0.942 (DL) | 0.100 (BE)<br>0.210 (HM)<br>0.214 (DL) | <0.001 | 230 | 23 | 207 | 9.00 |
| 18 | Figure 2 | abdominal | 0.841 | 0.788-0.886 (BE)<br>0.754-0.929 (HM)<br>0.753-0.930 (DL) | 0.098 (BE)<br>0.175 (HM)<br>0.177 (DL) | <0.001 | 234 | 27 | 207 | 7.67 |
| 19 | Figure 2 | pulmonary | 0.861 | 0.813-0.901 (BE)<br>0.801-0.921 (HM)<br>0.801-0.921 (DL) | 0.088 (BE)<br>0.120 (HM)<br>0.120 (DL) | <0.001 | 257 | 50 | 207 | 4.14 |

p* = probability that the observed AUC is different from AUC 0.5 under the null hypothesis (Hanley & McNeil, 1982; Zweig & Campbell, 1993; Shoonjans, 2017).

***Supplement Table S13:*** *ROC analyses for different strata, based on clinical variables (calculations with MedCalc software).* Abbreviations: CI, confidence interval; BE, binomial exact; HM, Hanley & McNeill (1982); DL, DeLong et al. (1988); BRNNNN, bootstrap resampling with NNNN cycles. Entries arranged in correspondence to Table T5.

| **No.** | **Reference** | **Stratum** | **AUC** | **95% CI** | **CI width** | **p*** | **N** | **Case (sepsis)** | **Control (SIRS)** | **Control/case (ratio)** |
|---|---|---|---|---|---|---|---|---|---|---|
| 20 | Table T5 | platelets > 445,000/uL | 0.734 | 0.460-0.918 (BE) | 0.458 (BE) | 0.09 | 16 | 8 | 8 | 1 |
| | | | | 0.470-0.999 (HM) | 0.529 (HM) | | | | | |
| | | | | 0.459-1.000 (DL) | 0.541 (DL) | | | | | |
| | | | | 0.469-0.977 ($BR_{2000}$) | 0.508 ($BR_{2000}$) | | | | | |
| | | | | 0.477-0.969 ($BR_{5000}$) | 0.492 ($BR_{5000}$) | | | | | |
| 21 | Table T5 | Lactate < 1 mmol/L | 0.836 | 0.629-0.954 (BE) | 0.325 (BE) | <0.001 | 24 | 7 | 17 | 2.43 |
| | | | | 0.672-1.000 (HM) | 0.328 (HM) | | | | | |
| | | | | 0.669-1.000 (DL) | 0.331 (DL) | | | | | |
| 22 | Table T5 | PCT < 0.5 ng/mL | 0.707 | 0.629-0.778 (BE) | 0.149 (BE) | <0.001 | 154 | 32 | 122 | |
| | | | | 0.605-0.810 (HM) | 0.205 (HM) | | | | | |
| | | | | 0.604-0.811 (DL) | 0.207 (DL) | | | | | |
| 23 | Table T5 | MAP < 70 mm Hg always | 0.817 | 0.702-0.901 (BE) | 0.199 (BE) | <0.001 | 66 | 33 | 33 | 1 |
| | | | | 0.710-0.923 (HM) | 0.213 (HM) | | | | | |
| | | | | 0.710-0.924 (DL) | 0.214 (DL) | | | | | |
| 24 | Table T5 | MAP 70-100 mm Hg | 0.825 | 0.709-0.909 (BE) | 0.200 (BE) | <0.001 | 63 | 28 | 35 | 1.25 |
| 25 | Table T5 | MAP > 100 and never < 70 mm Hg | 0.855 | 0.754-0.926 (BE) | 0.172 (BE) | <0.001 | 74 | 18 | 56 | 3.11 |

| No. | Reference | Stratum | AUC | 95% CI | CI width | p* | N | Case (sepsis) | Control (SIRS) | Control/case (ratio) |
|---|---|---|---|---|---|---|---|---|---|---|
| 26 | Table T5 | MAP > 100 mm Hg at least 1x | 0.85 | 0.788-0.899 (BE) | 0.111 (BE) | <0.001 | 173 | 58 | 115 | 1.98 |
| 27 | Table T5 | MAP < 70 and MAP > 100 mm Hg | 0.863 | 0.780-0.924 (BE) | 0.144 (BE) | <0.001 | 99 | 40 | 59 | 1.48 |
| 28 | Table T5 | RR.max ≤ 21 breaths/min | 0.821 | 0.676-0.920 (BE)<br>0.660-0.981 (HM)<br>0.654-0.987 (DL) | 0.244 (BE)<br>0.321 (HM)<br>0.333 (DL) | <0.001 | 44 | 9 | 35 | 3.89 |
| 29 | Table T5 | Systolic BP .max/min >100 mm Hg | 0.836 | 0.718-0.954 (HM) | 0.24 (HM) | <0.001 | 64 | 17 | 47 | 2.76 |

p* = probability that the observed AUC is different from the null hypothesis value of 0.5 (Hanley & McNeil, 1982; Zweig & Campbell, 1993; Shoonjans, 2017).

### 6.3. Further Investigation of Bootstrap Estimates of CI

We investigated the variability in bootstrap resampling estimates of AUC and CI, for a moderate-sized dataset (MAP<70 mm Hg) consisting of 33 sepsis, 33 SIRS. Different numbers of bootstrap resamples were used, as well as different random seed values.

***Supplement Table S14:*** *summary of AUC estimation results, for a moderately-sized dataset.* The dataset used in this example was MAP<70 (33 sepsis, 33 SIRS). AUC CI calculations by the HM, DL and BE methods used MedCalc (www.medcalc.org), while the bootstrapping used an online applet at http://www.freccom.cz/stomo/input.php (Skalsk´a & Freylich, 2006).

| **Method** | **AUC** | **95% CI** | **95% CI width** |
|---|---|---|---|
| De Long | 0.817 | 0.710-0.924 (±1.96 x 0.0547) | 0.214 |
| Hanley & McNeil | 0.817 | 0.710-0.923 (±1.96 x 0.0544) | 0.213 |
| Binomial Exact | 0.817 | 0.702-0.901 | 0.199 |
| Bootstrap x 1,000 | 0.807 | 0.694-0.908 | 0.214 |
| | 0.802 | 0.692-0.907 | 0.215 |
| | 0.805 | 0.694-0.906 | 0.212 |
| Bootstrap x 2,000 | 0.799 | 0.682-0.900 | 0.218 |
| | 0.803 | 0.690-0.906 | 0.216 |
| | 0.800 | 0.691-0.896 | 0.205 |
| | 0.802 | 0.689-0.903 | 0.214 |
| | 0.802 | 0.694-0.903 | 0.209 |
| Bootstrap x 3,000 | 0.805 | 0.689-0.904 | 0.215 |
| | 0.802 | 0.692-0.900 | 0.208 |
| Bootstrap x 4,000 | 0.803 | 0.690-0.901 | 0.211 |
| | 0.803 | 0.691-0.903 | 0.212 |

The HM, DL and BE methods produce equivalent AUC estimates (0.817), while the bootstrap method produces an average AUC estimate of 0.803 which is 0.14 units lower – a difference which is significant at $p<0.001$. The width of the 95% CI appears the same for all methods and bootstrap attempts. Bootstrap trials with different random seeds and different numbers of iterations produced essentially the same results. An example bootstrap analysis is shown in **Figure S8**.

***Supplement Figure S8:*** *Example of bootstrap resampling AUC distribution for a moderate-sized balanced comparison (MAP<70 mm Hg group, 33 sepsis, 33 SIRS).* This particular resampled AUC distribution is centered at 0.799.

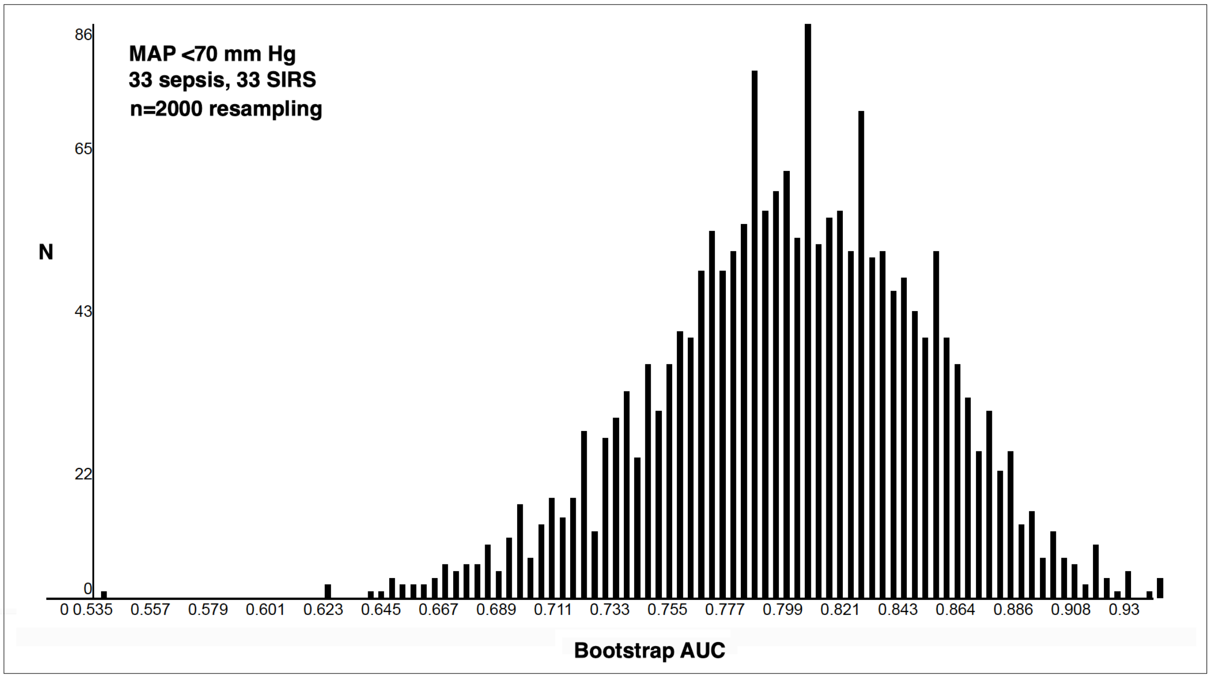

We next considered the smallest of our stratification analyses, lying just below the edge of significance: platelets >445,000/uL with 8 sepsis and 8 SIRS.

Student's t-test gave p = 0.066 for separation of SIRS and sepsis groups. Initial ROC calculations gave AUC 0.73 (95% CI 0.460-0.918) with p-value 0.083-0.095. Calculation of the Mann-Whitney U statistic gave U = 47 for the higher ranking group, which when divided by $n_1 * n_2$ yielded 47/(8*8) = 0.734, a Z score of 1.5228 and a p-value of 0.643, which again falls below the conventional significance cutoff of p=0.05.

The HM, DL and BE methods all estimate the value AUC 0.734, while the Bootstrap method produces an estimate of AUC 0.743 which is 0.09 units higher (p<0.001). The width of the 95% CI appears the same for all estimates. Bootstrap trials with different seeds and different numbers of iterations produced essentially the same results.

***Supplement Table S15:*** *Summary of AUC estimation results, for a small dataset on the edge of significance.* The dataset used in this example was platelets >445,000/uL (8 sepsis and 8 SIRS). AUC CI calculations by methods of DeLong, Hanley & McNeil and binomial exact used MedCalc, while the bootstrapping used an online applet at the following website: http://www.freccom.cz/stomo/input.php (described in Skalsk´a & Freylich, 2006).

| **AUC Method** | **AUC** | **95% CI** | **95% CI width** |
|---|---|---|---|
| De Long | 0.734 | 0.459-1.000 (±1.96 x 0.141) | 0.541 |
| Hanley & McNeil | 0.734 | 0.470-0.999 (±1.96 x 0.135) | 0.529 |
| Binomial exact | 0.734 | 0.460-0.918 | 0.458 |
| Bootstrap x 1,000 | 0.748 | 0.469-0.984 | 0.515 |
| | 0.742 | 0.453-0.984 | 0.531 |
| | 0.744 | 0.469-0.969 | 0.500 |
| Bootstrap x 2,000 | 0.746 | 0.476-0.969 | 0.493 |
| | 0.743 | 0.453-0.977 | 0.524 |
| | 0.744 | 0.477-0.969 | 0.492 |
| Bootstrap x 3,000 | 0.747 | 0.477-0.977 | 0.500 |
| | 0.739 | 0.469-0.969 | 0.500 |
| | 0.742 | 0.445-0.977 | 0.532 |
| Bootstrap x 4,000 | 0.744 | 0.477-0.969 | 0.492 |
| | 0.742 | 0.469-0.977 | 0.508 |
| | 0.742 | 0.445-0.969 | 0.524 |
| Bootstrap x 5,000 | 0.745 | 0.484-0.969 | 0.485 |
| | 0.741 | 0.461-0.969 | 0.508 |
| | 0.744 | 0.469-0.969 | 0.500 |

When a plot of the bootstrapping results is examined graphically (**Figure S9**), the distribution unquestionably falls above 0.5 indicating discrimination between the sepsis and SIRS groups, even though the p-values in Student's t, Mann-Whitney U and ROC tests fall below p=0.05. For AUC evaluations with this small stratum, although the calculated ROC curves all fall around 0.73-0.75 and thus lie well above the 0.5 null value, they nonetheless have wide CIs which limits their usefulness in supporting a quantitative performance claim.

***Supplement Figure S9:** Example of bootstrap resampling distribution for small stratum (platelets >445,000/uL), consisting of 8 sepsis and 8 SIRS patients.* This particular bootstrap resampling involved 2,000 repetitions, and is centered around 0.74. Fifty histogram bins were used to construct this plot. The right side of the resampling distribution is truncated at AUC 1.000.

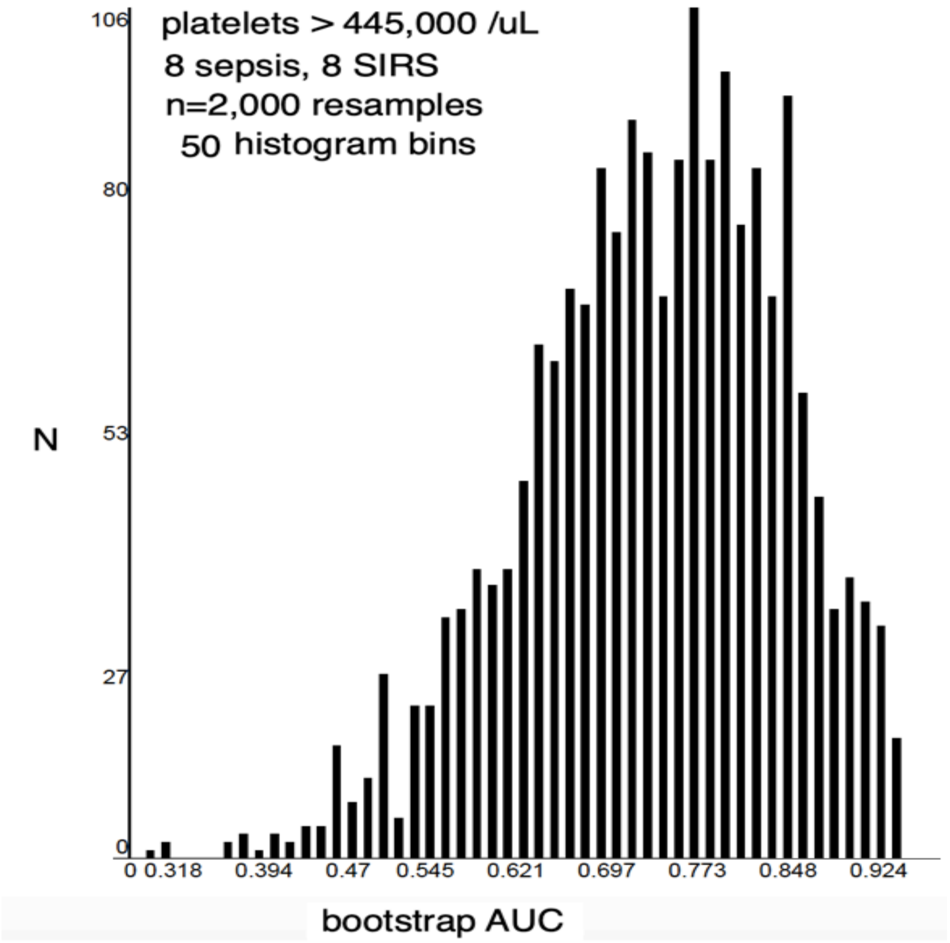

### 7. References

Anonymous (biyee.net). Binomial Confidence Interval and its application in reliability tests. **2019**. Available online: https://www.biyee.net/data-solution/resources/binomial-confidence-interval.aspx (accessed August 19, 2024).

Cohen, J. A coefficient of agreement for nominal scales. *Educ. Psychol. Meas.***1960**, *20*, 213-220. DOI: 10.1177/001316446002000104.

DeLong E.R., DeLong D.M., Clarke-Pearson D.L. Comparing the areas under two or more correlated receiver operating characteristic curves: a nonparametric approach. *Biometrics.* **1988**, *44*, 837-845. DOI: 10.2307/2531595.

Fleiss, J.L., Cohen, J., and Everitt, B.S. Large sample standard errors of kappa and weighted kappa. *PsychoL. Bull.* **1969**, *72*, 323-327. DOI:10.1037/h0028106.

Goldstein-Greenwood, J. (UVA Library StatLab). ROC Curves and AUC for Models Used for Binary Classification. **2022**, Available online*:* https://library.virginia.edu/data/articles/roc-curves-and-auc-for-models-used-for-binary-classification (accessed August 19, 2024).

Hanley J.A., McNeil B.J. The meaning and use of the area under a receiver operating characteristic (ROC) curve. *Radiology*. **1982**, *143*, 29-36. DOI: 10.1148/radiology.143.1.7063747.

Kuye I., Anand V., Klompas M., Chan C., Kadri S.S., Rhee C. Prevalence and Clinical Characteristics of Patients With Sepsis Discharge Diagnosis Codes and Short Lengths of Stay in U.S. Hospitals. *Critical Care Explor.* **2021**, *3*, e0373. DOI: 10.1097/CCE.0000000000000373.

Mason S.J., Graham N.E. Areas beneath the relative operating characteristics (ROC) and relative operating levels (ROL) curves: Statistical significance and interpretation. *Q. J. R. Meteorol. Soc.* **2002**, *128*, 2145–2166. doi:10.1256/003590002320603584.

Qin C., Zhang S., Zhao Y., Ding X., Yang F., Zhao Y. Diagnostic value of metagenomic next-generation sequencing in sepsis and bloodstream infection. *Front. Cell. Infect. Microbiol.* **2023**, *13*, 1117987. DOI: 10.3389/fcimb.2023.1117987.

Shoonjans, F. MedCalc manual: Easy-to-use statistical software. **2017.** MedCalc Software bvba, Ostend, Belgium. ISBN 978-1520321578.

Skalsk´a H., Freylich V. (2006) Web-Bootstrap Estimate of Area Under ROC Curve. *Austrian J Stat.* **2006**, *35*, 325–330. DOI: 10.17713/ajs.v35i2&3.379.

Xu, W., Dai, J., Hung, Y. S., & Wang, Q. Estimating the area under a receiver operating characteristic (ROC) curve: Parametric and nonparametric ways. *Signal Process.* **2013***, 93,* 3111–3123. DOI:10.1016/j.sigpro.2013.05.010.

Yang L., Lin Y., Wang J., Song J., Wei B., Zhang X., et al. Comparison of Clinical Characteristics and Outcomes Between Positive and Negative Blood Culture Septic Patients: A Retrospective Cohort Study. *Infect Drug Resist.* **2021**, *14*, 4191–205. DOI: 10.2147/IDR.S334161.

Zhou X.H., Obuchowski N.A., McClish D.K. Statistical methods in diagnostic medicine, 2$^{nd}$ Edition. John Wiley and Sons, Hoboken, New Jersey, USA. **2011.** ISBN: 978-0-470-18314-4.

Zweig M.H., Campbell G. Receiver-operating characteristic (ROC) plots: a fundamental evaluation tool in clinical medicine. *Clin. Chem.* **1993**, *39*, 561-577. DOI: 10.1093/clinchem/39.4.561. Erratum in: *Clin. Chem.* **1993**, *39*, 1589. DOI: 10.1093/clinchem/39.8.1589.
